# A Systematic Process for Assessing Fitness-for-Purpose of Health Outcomes for Computable Phenotyping with Electronic Health Record Data

**DOI:** 10.1101/2025.08.29.25334394

**Authors:** Nicole M. Gatto, David J. Cronkite, Paige D. Wartko, Robert Ball, David S. Carrell, Rhoda Eniafe, Rishi Desai, James S. Floyd, Terrence Lee, Jennifer C. Nelson, Fatma M. Shebl, Ryan Schoeplein, Sengwee Toh, Mingfeng Zhang, Sascha Dublin, José J. Hernández-Muñoz

## Abstract

**Purpose:** Information from electronic health records (EHRs) may be incorporated into computable phenotype algorithms in efforts to overcome inaccuracies of algorithms based on administrative claims data alone. However, such efforts can be resource-intensive and unsuccessful. Assessing the feasibility of computable phenotyping for a health outcome of interest (HOI) before proceeding is therefore recommended.

**Methods:** We developed a systematic fitness-for-purpose (FFP) assessment process to implement concepts outlined in a previously described general framework for computable phenotyping incorporating EHR data. Our process includes verifying the HOI is well-defined, reviewing clinical information about the HOI, identifying existing algorithms and their performance, evaluating HOI clinical and data complexity, and determining an overall FFP conclusion and recommendation. We applied this process to ten HOIs lacking high-performing claims-based algorithms, selecting HOIs of public health importance that varied in clinical and data complexity, including neutropenia, pericardial effusion and drug-induced liver injury.

**Results:** HOIs assessed as having moderate (vs. easy) overall difficulty had characteristics such as the need for natural language processing, integration of multiple laboratory test results, or longitudinal EHR data. HOIs assessed as having high difficulty required using data from multiple EHR sources, ruling out many other potential causes, or relying on low-sensitivity diagnostic tests. Input from experts in EHR data and clinical care was crucial.

**Conclusion:** EHR data have potential to enhance accuracy of defining certain HOIs for research and surveillance compared to administrative claims data. The process and tools we created will support others in assessing FFP of HOIs for computable phenotyping.

**Five key points:**

- Incorporating electronic health record (EHR) data into computable phenotypes could improve accurate identification of health outcomes of interest (HOIs), but such work can be resource intensive.
- We developed a systematic fitness-for-purpose (FFP) process and tools to assess the feasibility of computable phenotyping for HOIs.
- Steps include identifying existing algorithms and their performance, ensuring the HOI is well-defined, evaluating clinical and data complexity, and determining a feasibility recommendation.
- Difficulty increased with a need for natural language processing, multiple laboratory tests, longitudinal EHR data, multiple EHR sources or ruling out other potential causes.
- Input from EHR data and clinical care experts was crucial to the FFP assessment process.

**Plain Language Summary (PLS):** Attempts to identify diseases and health conditions by applying computer programs to information easily gleaned from insurance claims of tens of thousands of patients (such as FDA’s ongoing safety monitoring of approved drugs or medical products) are often unsuccessful because the data lack nuance. Incorporating information from electronic health records (EHR) and patient chart notes may improve accurate identification of health outcomes. Because this can be resource-intensive, we designed a process and tools to assess the feasibility of including EHR data in computer algorithms to identify health outcomes. Steps included identifying existing algorithms and their performance, building familiarity with the outcome and making sure it is well-defined, evaluating clinical and data complexity, and determining a conclusion about feasibility. We applied our process to ten health outcomes of public health importance. Health outcomes were considered moderately difficult for computerized algorithms if they required natural language processing, integration of multiple laboratory tests, or EHR data from multiple timepoints. Health outcomes having high difficulty required using multiple EHR data types, ruling out many alternative causes of the HOI (other than medications), or relying on diagnostic tests of low accuracy. Input from EHR data and clinical care experts was crucial for the assessment process.

## PURPOSE

Administrative claims are a major data source used in large-scale medical research and public health surveillance to identify diseases and other health outcomes of interest (HOI). Key strengths of claims data include the comprehensive capture of outpatient pharmacy dispensing, medical encounters, and hospitalizations during well-defined enrollment periods for large patient populations in addition to being relatively easy to obtain. In the United States (US) Food and Drug Administration’s (FDA) Sentinel System (“Sentinel”),^1^ for medical product safety surveillance, when a safety concern is identified prior to product approval, the FDA first determines whether it can be accurately assessed using adverse event reports or existing administrative claims data^2^, before requiring the manufacturer to conduct studies for a post-marketing requirement (PMR).^2^ The FDA forms this assessment by evaluating five “domains” of epidemiologic study design: study population, exposure, health outcome, covariates, and analytic tools.^3^ Because administrative claims data are designed for billing purposes, these data are frequently insufficient to evaluate a safety concern because of a lack of granular clinical information. An analysis of the first six years of Sentinel’s Active Risk Identification and Analysis (ARIA) post-marketing surveillance system indicated that only 133 of 330 safety concerns (40%) could be sufficiently evaluated using ARIA^2,4^, citing the inability to accurately identify an HOI as the most frequent obstacle. In response, Sentinel expanded access to clinical data for large populations by creating the Real-World Evidence Data Enterprise (RWE-DE), a network linking claims and electronic health record (EHR) data for >25 million individuals across six sites.

Sentinel is developing tools and processes to integrate these data into routine monitoring.^5-7^ Validation of existing HOI algorithms and development of new EHR-based HOI algorithms will be essential to utilize RWE-DE in a scalable way for research and surveillance.^8^ In some instances, it may be necessary to augment claims data by creating more complex computable phenotypes (computerized algorithms in an EHR system or clinical data repository using a defined set of data elements and logical expressions)^9,10^ ^11^. Computable phenotyping approaches often require moving beyond standard rule-based approaches and utilizing more advanced methods such as natural language processing (NLP) and machine learning (ML). Studies have shown early promise in developing EHR-based algorithms for acute pancreatitis,^12^ anaphylaxis,^13^ rhabdomyolysis,^14^ and COVID-19 disease.^15^ For acute pancreatitis, incorporating EHR data posed minimal burden and yielded large gains in accuracy: integrating one laboratory test result, lipase, with existing diagnosis codes yielded a highly accurate algorithm (positive predictive value [PPV] 92%, sensitivity 85%) superior to diagnosis codes alone (PPV=61%). For anaphylaxis, developing a sufficiently accurate EHR-based algorithm proved more challenging. It involved using NLP to extract a wide range of features from clinical notes curated with guidance from clinical and informatics experts. These two examples demonstrate the range of difficulty encountered in creating EHR-based HOI algorithms.

Because the development of computable phenotypes can be resource- and labor-intensive,^13,16^ it is worthwhile to prioritize HOIs where reasonable effort is likely to achieve sufficiently high performance. As part of a general framework proposed for phenotype development, Carrell et al. recommended an early ‘fitness-for-purpose’ (FFP) assessment to determine whether an HOI is a good candidate prior to committing resources to phenotyping with EHR data.^17^ They proposed that the HOI’s clinical and data complexity be assessed first. Clinical complexity refers to the difficulty a clinician experiences in diagnosing an HOI. Data complexity refers to the difficulty of identifying the needed information from electronic healthcare data, such as insurance claims or EHR. Carrell et al. described a conceptual approach but did not operationalize or prospectively implement the FFP assessment process.

To address this gap, we created and pilot-tested a systematic process for FFP assessment to build on the proposed general framework. Our team of clinicians, epidemiologists and data informatics specialists developed an initial process and refined it iteratively through application to varied HOIs lacking high-performing claims-based algorithms. Here we describe the FFP assessment process, present results for ten HOIs, and share key lessons learned. We provide supporting tools including protocols and worksheets, intended to be transferable to other HOIs and settings. Our objective is to facilitate systematic assessment of feasibility and likelihood of success when developing a computable phenotype for an HOI using clinical EHR data.

## METHODS

Our protocol for FFP assessment (Appendix 1) includes a series of steps (**Figure 1**) described in greater detail below. While a generalist can lead the process, the protocol assumes the ability to consult experts in clinical medicine and informatics.

**Figure 1.**
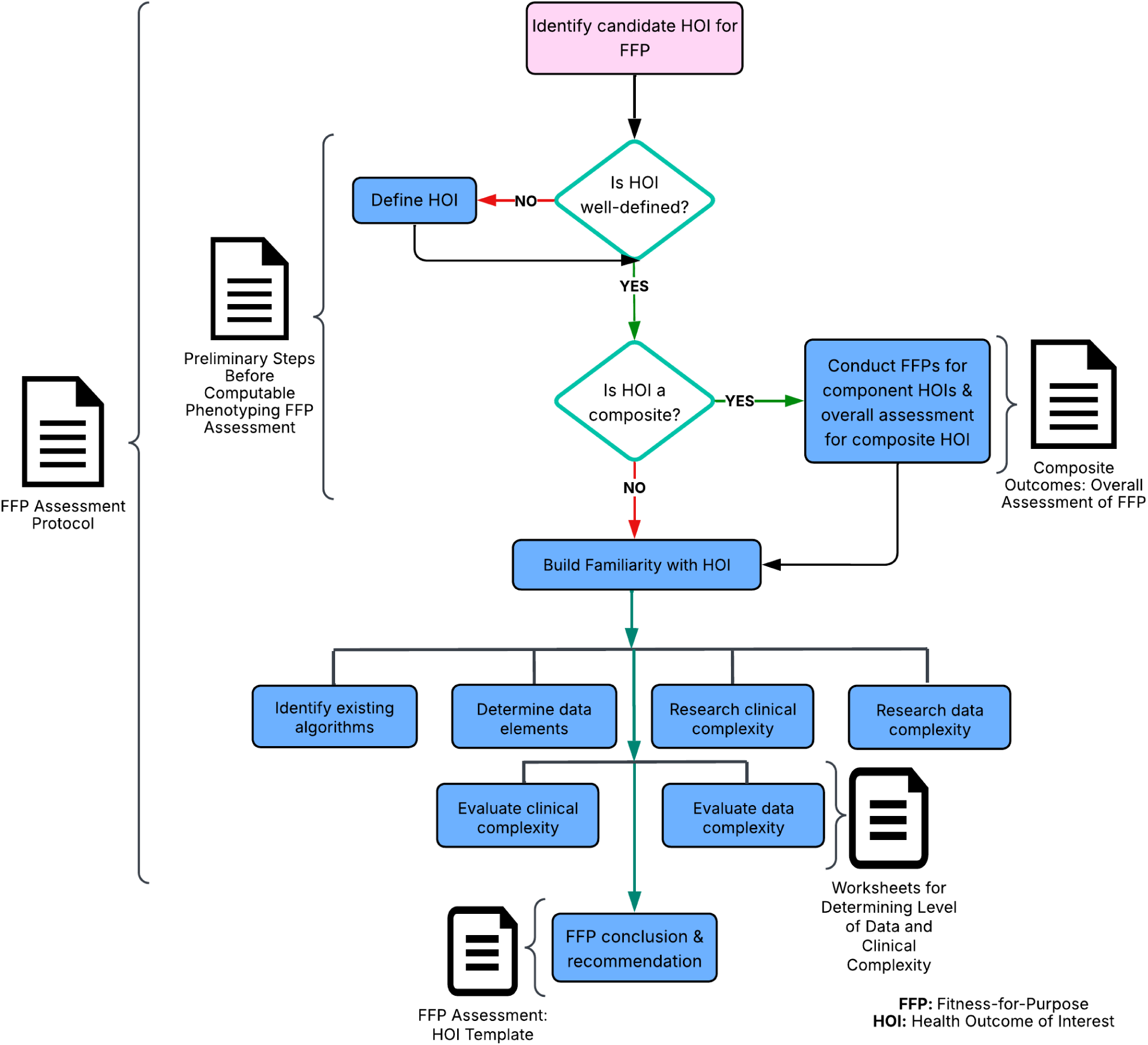
Fitness-For-Purpose (FFP) Assessment Process Flow Chart and Accompanying Tools to Assist with Each Step.

### Check whether the HOI is well-defined

We first verified the HOI was well-defined as a specific medical condition(s) that can be diagnosed in a healthcare setting and for which there is general agreement among health professionals about what is meant by the condition(s) (Appendix 2).^11^ Assessment of overly broad HOIs (e.g., “hematologic disorders,” which encompasses a variety of problems with different blood components) could not proceed. We attempted to narrow down and clarify such HOIs by reviewing medical references and consulting with experts.

### Review clinical information about the HOI

We searched clinical knowledge bases (e.g., UpToDate), reviewed medical society guidelines, patient-facing websites (e.g., National Institutes of Health, Cleveland Clinic), and FDA clinical guidance.^18^ This step provided foundational information about a generally accepted clinical definition, how the HOI is diagnosed, what level of agreement is likely present between clinicians, and which EHR data elements would be needed to develop a computable phenotype (informing a later step).

We reviewed reasons for insufficiency of claims-based data in relevant ARIA memos (Appendix 3) to ensure we understood FDA’s original concerns related to insufficiency.

### Identify existing HOI algorithms and their performance

We searched the literature for studies describing (and ideally, validating) HOI algorithms based on diagnostic codes, procedure codes and/or EHR data, noting when no studies could be found. When multiple algorithms were found, we prioritized those based on International Classification of Diseases, 10^th^ Revision, Clinical Modification (ICD-10-CM) rather than ICD-9-CM codes given relevance to future surveillance activities. We assessed the accuracy of algorithms based on PPV and sensitivity. When we found multiple validation studies, we prioritized US-based studies as more relevant to Sentinel due to differences between countries in healthcare practices, systems and coding. We focused on the “gold standard” used in algorithm development and validation (e.g., chart review) as this knowledge informed our interpretation of performance measures and in some cases helped us understand how EHR data could be leveraged to better identify the HOI.

### Evaluate clinical and data complexity of the HOI

Building on the general framework,^17^ we operationalized concepts of clinical and data complexity (Appendix 4). We considered various domains of clinical and data complexity affecting the difficulty of developing a computable phenotype algorithm for an HOI (Table 1). We assigned a rating of low, medium, or high complexity for each domain and provided a rationale.

**Table 1.**
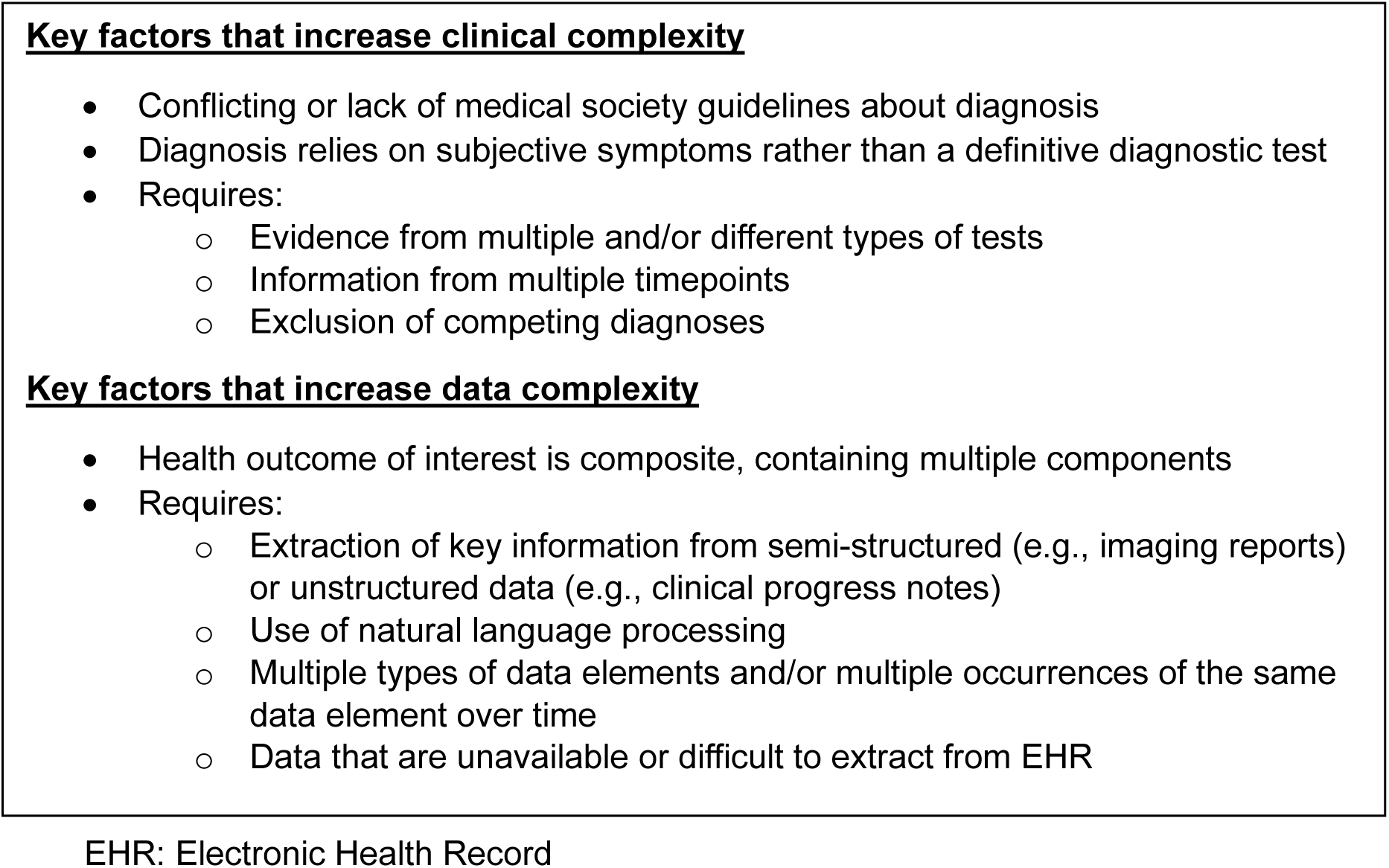
Characteristics that Increase Clinical and Data Complexity.

### Determine an overall FFP conclusion and recommendation

We used all acquired information to assign each HOI an overall difficulty rating of easy, moderate, or hard. When clinical and data complexity ratings were similar, reaching an overall assessment was usually straightforward; if they were discordant, it was more challenging.

Generally, the highest level of either clinical or data complexity drove the overall determination. In addition to the FFP conclusion and rationale, we recommended next steps, such as describing further preliminary work or suggesting additional questions to be answered prior to undertaking phenotype development.

Findings were summarized using the Fitness-for-Purpose Assessment HOI Template (Appendix 5).

### Composite HOIs (Appendix 6)

Composite HOIs represent a group of related conditions or events. We approached the assessment of composite outcomes, such as “major bleed” and “serious infection”, differently than HOIs representing a single health condition, because within a composite HOI, each component has its own clinical and data complexity. First, we specified the component HOIs that make up the composite. We next conducted a separate assessment for each component. The overall conclusion weighed and synthesized the clinical and data complexity of individual components. We factored in how much flexibility is tolerated in the components, how many components comprise the composite, and the potential overlap in the work needed to develop a phenotype for each component.

## RESULTS

Our overall FFP assessment of ten candidate HOIs with accompanying clinical and data complexity are summarized in Table 2. Figure 2 shows the relationship between overall phenotype difficulty and clinical and data complexity. We describe in detail below three HOIs with easy, moderate, and hard overall difficulty.

**Figure 2.**
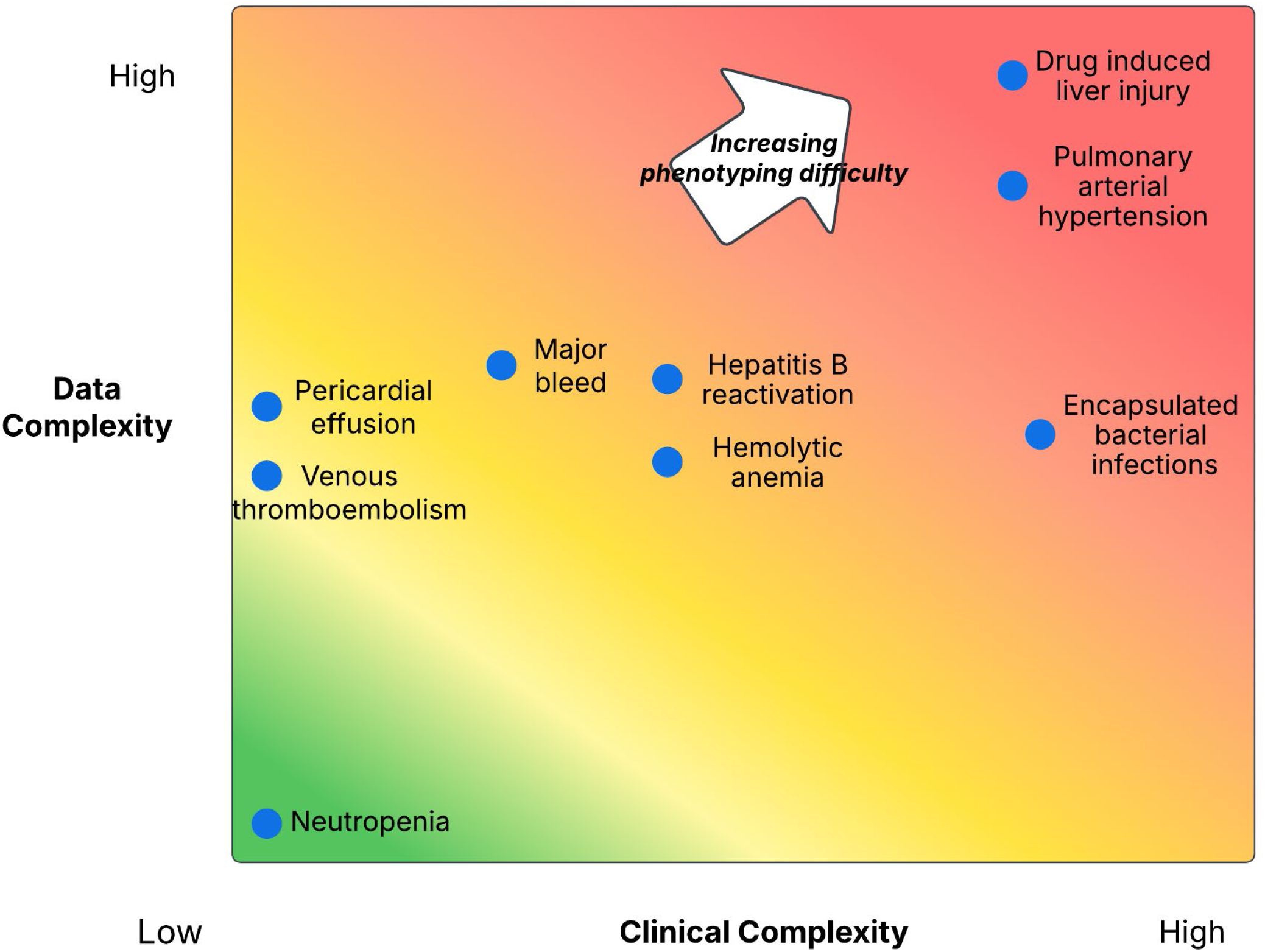
Graph of Clinical and Data Complexity of Health Outcomes of Interest (HOIs) Evaluated by Fitness-for-Purpose (FFP) Process.

**Table 2.** Candidate Health Outcomes of Interest (HOIs) Evaluated with the Fitness-for-Purpose (FFP) Process and Results of the Assessment.

| HOI | Definition | Reason(s) claims data found insufficient† | Clinical complexity | Data complexity | Overall assessment |
| --- | --- | --- | --- | --- | --- |
| Serious infections* | No clear or widely-accepted definition | 4 | Not assessed | Not assessed | Not assessed |
| Neutropenia | A deficiency of a type of white blood cells, leading to an increased risk of bacterial infection | 1, 3 | Low | Low | Easy |
| Pericardial effusions | Accumulation of fluid within the pericardial sac (around the heart) | 2 | Low | Medium | Moderate |
| Venous thromboembolism | Blood clot obstructing blood vessels, including deep vein thrombosis, typically occurring in the upper or lower extremities, and pulmonary embolism, affecting arteries within the lungs | 3 | Low | Medium | Moderate |
| Major bleed* | A composite outcome with multiple components, including (1) an intracranial hemorrhage (a bleed within the brain) OR (2) an extracranial hemorrhage a) requiring red blood cell or whole blood transfusion, or b) involving critical anatomical sites, specifically, intraarticular (in a joint space), pericardial (around the heart) or retroperitoneal (abdominal), or c) that was fatal (resulting in death within 30 days of the bleeding event) | 1, 3, 5 | Low Medium | Medium | Moderate |
| Hepatitis B reactivation (HBV-R) | A clinical syndrome characterized either by 1) the reappearance of serum HBV DNA or development of Hepatitis B surface antigen (HBsAg) in individuals with a previous resolved HBV infection, or 2) a substantial increase in serum HBV DNA in individuals with inactive chronic hepatitis B (CHB). HBV-R can occur spontaneously but is often associated with a compromised immune system (e.g., immunosuppressive therapies, autoimmune disease, or organ transplantation) | 1, 2 | Medium | Medium | Moderate |
| Hemolytic anemia | Anemia due to the destruction (hemolysis) of red blood cells (RBCs) in which the production of new RBCs is unable to keep up with the pace of destruction | 1, 3 | Medium | Medium | Moderate |
| Encapsulated bacterial infections | Infections due to certain bacteria (typically *Streptococcus pneumoniae*, *Neisseria meningitidis* and *Hemophilus influenzae*) that are encased in a polysaccharide capsule which allows them to evade the host immune system and phagocytosis | 3 | High | Medium | Hard |
| Drug-induced liver injury (DILI) | An injury to the liver caused by a medication, herbal remedy, or dietary supplement | 1 | High | High | Hard |
| Pulmonary arterial hypertension (PAH) | A form of pulmonary hypertension, which is high blood pressure in the lungs | 3 | High | High | Hard |

HOI: Health Outcome of Interest
\*Composite HOI
†Initial reason(s) why the HOI was unable to be studied in the FDA Sentinel ARIA System. 1: Supplemental structured clinical data needed, 2: Data elements captured in unstructured clinical data needed, 3: Absence of validated claims-based algorithm, 4: Identification of clinical concepts is not possible or inadequate, 5: Requires linkage to additional data source that is unavailable.

### Neutropenia (Appendix 7)

We assigned an overall difficulty level of “easy” to neutropenia and concluded that computable phenotyping of this HOI using EHR data has a high likelihood of success. A neutropenia diagnosis is based on objective findings, specifically, results from a complete blood count with differential. The laboratory test needed for diagnosis is commonly ordered, yields numerical results, and has an uncomplicated interpretation. Clinical complexity was rated low based on the HOI being straightforward to diagnose. The required laboratory results are recorded in structured form, aligning with low data complexity. A computable phenotype could be created by extracting laboratory values and comparing to a selected cut-off value. Work would be needed to harmonize data across tests and clinical sites due to different laboratory reporting practices for neutrophil values but was determined to be feasible.

We noted the lack of adequate information about validity of existing claims-based algorithms^19,20^. We found no large-scale validation studies using ICD-10-CM codes. Thus, we recommend a validation study of ICD-10-CM codes as a first step prior to investing resources in developing a new computable phenotype based on EHR laboratory data.

### Pericardial Effusion (Appendix 8)

Our assessment concluded that pericardial effusion has “moderate” overall difficulty, indicating good suitability and high likelihood of success for computable phenotyping. The need for more complex methods to extract data drove our determination. Pericardial effusion is readily detected by transthoracic echocardiography (the “gold standard” diagnostic test), as well as computed tomography or magnetic resonance imaging,^21^ contributing to a clinical complexity rating of low. Phenotyping would require applying NLP to imaging reports, which have substantial semi-structured free text and standardized language, making NLP more straightforward than other types of free text (e.g., clinical notes). Keyword-based NLP algorithms have shown good performance for pericardial effusions.^22-24^ These factors led to a data complexity rating of medium. Because there are no validated algorithms using ICD-10-CM codes to identify pericardial effusion, we recommend starting with a validation study focused on ICD-10-CM code-based algorithms.

### Drug-Induced Liver Injury (DILI) (Appendix 9)

We concluded that using EHR data to capture DILI through a computable phenotype would be complex and may not achieve desired accuracy, earning DILI an overall assessment of hard. While professional society guidelines for DILI exist,^25,26^ they differ considerably, for instance regarding which laboratory tests are included and the number of timepoints required. Available clinical practice diagnostic algorithms lack clarity and proven accuracy.^26^ Numerous other potential causes of liver injury must be ruled out for a DILI diagnosis to be made. Together, these features make clinical complexity high. Data complexity is also high, as a computable phenotype would likely require longitudinal laboratory values in addition to diagnosis codes and features derived from NLP. An existing algorithm which used laboratory tests medications, ICD-9-CM codes, and NLP of clinical notes had poor performance.^27^ Algorithms based on ICD-10-CM codes were examined in European validation studies, but they did not attempt to exclude other potential causes of liver injury,^28-30^ suggesting an opportunity to develop and validate a more comprehensive claims-based algorithm.

## DISCUSSION

We developed a systematic process to enable researchers and regulators to assess the feasibility of building computable phenotypes for HOIs using clinical EHR data. Our FFP assessment process yields a conclusion as to whether developing an EHR-based computable phenotype would be easy, moderate or hard, aiding in prioritization and resource allocation.

Additionally, we created tools to support the process including a protocol for FFP assessment and worksheets for assessing clinical and data complexity. This work was motivated by needs of the FDA’s Sentinel System, which is accessing and integrating EHR data to augment its current system that predominantly draws on claims data.

Our experience provides general insights that are relevant to FFP assessments of other HOIs. Moderate overall difficulty often resulted from HOIs involving NLP to process EHR data, which is more challenging to implement than extracting structured EHR data (e.g., laboratory test results). Other characteristics of HOIs with “moderate” difficulty were the need to integrate results from multiple laboratory tests and for longitudinal data to compare pre- and post-exposure results. HOIs determined to be “hard” often required using data from multiple EHR sources (including unstructured free text), ruling out many other potential causes, or relying on diagnostic tests with very low sensitivity.

We identified key lessons learned about the process. We recommend first considering whether the select HOI represents a specific well-defined medical condition. Inadequately defined HOIs should be refined and clarified prior to the assessment process. For example, FFP assessment of the “serious infections” HOI was not possible due to lack of consensus on which infections should be included, the definition of “serious”, and how nosocomial infections should be handled. Although it may seem desirable to rule out all alternative causes for an outcome in a regulatory setting, implementing this requirement dramatically increases data complexity and would have resulted in a determination of “hard” for virtually all HOIs, which would not be practical or realistic in a real-world data environment. Careful thought about when to require exclusion of alternative causes is necessary.

When no HOI algorithms based on diagnostic codes existed, or validation studies were outdated or from settings with limited relevance to Sentinel, we usually recommended that a validation study be completed for a claims-based algorithm before investing resources to develop a more complex EHR-based algorithm. An accurate claims-based algorithm is of value because it will increase applicability and power since claims data on large populations are currently more easily available than EHR data. We recognize that high-quality validation studies are costly. Additionally, claims-based algorithms for certain outcomes may be expected to have low validity and therefore would not be pursued.

Generally, determining the overall FFP assessment for an HOI is most straightforward when clinical and data complexity are of comparable levels, and more challenging when they are dissimilar. Nevertheless, there is no “one-size-fits-all” approach to an assessment; clinical and data complexity of HOIs are nuanced, and complexity criteria must be carefully weighed to reach an overall determination. The overall difficulty rating may not be the sole driver of whether to undertake phenotyping. Factors such as importance to research or surveillance may influence the decision of whether to undertake phenotyping, even if the HOI is determined to be hard. One approach to ascertaining important HOIs with high overall difficulty could be to use imperfect algorithms for initial case finding to improve sensitivity and then validate cases through chart review to enhance PPV.

In the process of iterating a systematic FFP process that could be applied across a variety of HOIs, our experience was that each HOI presented unique considerations. We consistently found practical skills, real-world knowledge and subject matter expertise invaluable. Consultation with experts on local EHR data systems is critical. For encapsulated bacterial infections, a physician-scientist team member shared key information that laboratory tests needed for diagnosis are often not ordered and when ordered are typically low-yield. This drove our assessment and was not readily apparent from an in-depth literature review. Similarly, communication with practicing physicians revealed the approach to diagnosing an intraarticular hemorrhage varies considerably by physician specialty and care setting. If evaluating teams do not include clinical and data experts, they should identify experts for consultation.

Given the recent rapid evolution in artificial intelligence, its potential for use in evaluating drug safety reports must be considered^1^. A key application of large language models (LLMs) to post-marketing drug safety surveillance is probabilistic phenotyping of HOIs.^31^ Matheny et al. identified two general areas in which LLMs might provide assistance: annotation of data and extraction of information from clinical narratives, biomedical literature and other knowledge sources. Based on our experience, we concur that LLMs may be helpful when assessing whether an HOI is well-defined. Nevertheless, we emphasize the need for human expertise to carefully scrutinize any LLM-derived information in the context of the real-world experience of health professionals in specific settings.

Our focus was on identifying health outcomes; we did not address other important aspects of study design^2^, such as defining the study population, characterizing exposure, and capturing important covariates. Our FFP process provides an assessment that is a “snapshot” at a specific time point. As clinical practice patterns, guidelines, technology, and coding patterns are constantly changing, FFP assessments should be updated periodically. While we selected a varied set of HOIs to pilot our FFP process, these ten likely do not capture all possible scenarios of clinical and data complexity. Additional HOIs should be assessed with our process to further evaluate its performance. It is unknown whether a computable phenotype will be transportable between different healthcare settings, particularly when NLP is required, as variation and heterogeneity in free text is likely greater than in structured data such as laboratory results. Questionable portability emphasizes the need for interchangeability and standardization across EHRs, especially in how and where information is stored. As our project did not in fact build computable phenotypes, we cannot confirm the accuracy of our predictions about the difficulty of building them. While our efforts are a step toward operationalization of an FFP process, more work is needed to understand how this process could be incorporated into a safety review framework.

Developing multi-site computable phenotyping algorithms can be time-consuming and expensive.^13,35,36^ It requires a range of skillsets, from clinicians to informatics experts to data programmers, to identify relevant data elements, manage missing or conflicting data, create “gold standard” cases for algorithm development and validation, and evaluate the proposed model, along with technical infrastructure to run the algorithm at multiple sites. An advantage of our FFP process is that it incorporates a “go/no-go” determination prior to committing to build a computable phenotype, reducing the risk of investing resources in developing HOI algorithms with a low likelihood of success. Other work in computable phenotype development such as by Observational Health Data Sciences and Informatics has emphasized data and technical solutions for representing information captured in EHRs.^32-34^ Ours is the first attempt that we know of to create a standardized approach guided by theory to assess the feasibility of computable phenotyping for HOIs.

In conclusion, EHR data such as laboratory results and clinical notes have potential to improve accuracy of identifying HOIs, compared to claims data. However, the process of integrating these data into computable phenotyping may be complex and expensive. We proposed a methodology for systematically assessing the overall difficulty of computable phenotype development that included evaluating data and clinical complexity. We hope that our process and practical tools will be useful to other groups and settings. The expansion of data resources and methods available to support research and surveillance such as applies to medication safety will require thoughtful evaluation of when and how to incorporate novel data sources and methods to optimize their use, with an ultimate goal of timely detection of risks from medical products to support safe use and ultimately, better population health.

## ETHICS STATEMENT

### Ethics approval statement

N/A. Not human subjects research.

### Patient consent statement

N/A. Informed consent was not applicable for this research.

## Data Availability

N/a no data used for manuscript

## ACKNOWLEDGEMENTS

We thank Linda Kiel for her contribution as project manager of the KPWHRI team.

## Permission to reproduce material from other sources

N/A. No other sources included.

## LIST OF APPENDICES

Appendix 1. Fitness-for-Purpose (FFP) Assessment Protocol

Appendix 2. Preliminary Steps Before Computable Phenotyping Fitness-for-Purpose (FFP) Assessment: Ensuring the Health Outcome of Interest (HOI) is Ready for Evaluation

Appendix 3. Select Publicly Available FDA ARIA Insufficiency Memos for Health Outcomes of Interest (HOI) Included in Fitness-for-Purpose (FFP) Assessment

Appendix 4. Worksheets for Determining Level of Data and Clinical Complexity

Appendix 5. Fitness-for-Purpose (FFP) Assessment: Health Outcome of Interest (HOI) Template

Appendix 6. Composite Outcomes: Overall Assessment of Fitness-for-Purpose (FFP)

Appendix 7. Fitness-for-Purpose (FFP) Assessment Report for Neutropenia

Appendix 8. Fitness-for-Purpose (FFP) Assessment Report for Pericardial Effusion

Appendix 9. Fitness-for-Purpose (FFP) Assessment Report for Drug-Induced Liver Injury (DILI)

Appendix 10. Fitness-for-Purpose (FFP) Assessment Report for Serious Infections

Appendix 11. Fitness-for-Purpose (FFP) Assessment Report for Venous Thromboembolism

Appendix 12. Fitness-for-Purpose (FFP) Assessment Report for Major Bleed

Appendix 13. Fitness-for-Purpose (FFP) Assessment Report for Hepatitis B Reactivation (HBV-R)

Appendix 14. Fitness-for-Purpose (FFP) Assessment Report for Hemolytic Anemia

Appendix 15. Fitness-for-Purpose (FFP) Assessment Report for Encapsulated Bacterial Infections

Appendix 16. Fitness-for-Purpose (FFP) Assessment Report for Pulmonary Arterial Hypertension (PAH)

## REFERENCES

1. Ball R, Robb M, Anderson SA, Dal Pan G. The FDA’s Sentinel Initiative--A comprehensive approach to medical product surveillance. Clin Pharmacol Ther. Mar 2016;99(3):265–8. doi:10.1002/cpt.320

2. Maro JC, Nguyen MD, Kolonoski J, et al. Six years of the US Food and Drug Administration’s postmarket active risk identification and analysis system in the Sentinel Initiative: Implications for real world evidence generation. Clin Pharmacol Ther. Oct 2023;114(4):815–824. doi:10.1002/cpt.2979

3. Sentinel. Assessing the ARIA System’s ability to evaluate a safety concern. Accessed March 25, 2025. https://sentinelinitiative.org/studies/drugs/assessing-arias-ability-evaluate-safety-concern

4. Brown JS, Maro JC, Nguyen M, Ball R. Using and improving distributed data networks to generate actionable evidence: The case of real-world outcomes in the Food and Drug Administration’s Sentinel system. J Am Med Inform Assoc. May 1 2020;27(5):793–797. doi:10.1093/jamia/ocaa028

5. Desai RJ, Marsolo K, Smith J, et al. The FDA Sentinel Real World Evidence Data Enterprise (RWE-DE). Pharmacoepidemiol Drug Saf. Oct 2024;33(10):e70028. doi:10.1002/pds.70028

6. Desai RJ, Matheny ME, Johnson K, et al. Broadening the reach of the FDA Sentinel system: A roadmap for integrating electronic health record data in a causal analysis framework. NPJ Digit Med. Dec 20 2021;4(1):170. doi:10.1038/s41746-021-00542-0

7. Schneeweiss S, Desai RJ, Ball R. A future of data-rich pharmacoepidemiology studies: Transitioning to large-scale linked electronic health record + claims data. Am J Epidemiol. Feb 5 2025;194(2):315–321. doi:10.1093/aje/kwae226

8. He T, Belouali A, Patricoski J, et al. Trends and opportunities in computable clinical phenotyping: A scoping review. J Biomed Inform. Apr 2023;140:104335. doi:10.1016/j.jbi.2023.104335

9. Richesson RL, Smerek MM, Blake Cameron C. A Framework to Support the Sharing and Reuse of Computable Phenotype Definitions Across Health Care Delivery and Clinical Research Applications. EGEMS (Wash DC). 2016;4(3):1232. doi:10.13063/2327-9214.1232

10. Masison J, Lehmann HP, Wan J. Utilization of Computable Phenotypes in Electronic Health Record Research: A Review and Case Study in Atopic Dermatitis. J Invest Dermatol. May 2025;145(5):1008–1016. doi:10.1016/j.jid.2024.08.025

11. Real-world data: Assessing electronic health records and medical claims data to support regulatory decision-making for drug and biological products guidance for industry (2024).

12. Floyd JS, Bann MA, Felcher AH, et al. Validation of acute pancreatitis among adults in an integrated healthcare system. *Epidemiology (Cambridge*, Mass). Jan 1 2023;34(1):33–37. doi:10.1097/ede.0000000000001541

13. Carrell DS, Gruber S, Floyd JS, et al. Improving methods of identifying anaphylaxis for medical product safety surveillance using natural language processing and machine learning. Am J Epidemiol. Feb 1 2023;192(2):283–295. doi:10.1093/aje/kwac182

14. Gibson TB, Nguyen MD, Burrell T, et al. Electronic phenotyping of health outcomes of interest using a linked claims-electronic health record database: Findings from a machine learning pilot project. J Am Med Inform Assoc. Jul 14 2021;28(7):1507–1517. doi:10.1093/jamia/ocab036

15. Smith JC, Williamson BD, Cronkite DJ, et al. Data-driven automated classification algorithms for acute health conditions: Applying PheNorm to COVID-19 disease. J Am Med Inform Assoc. Feb 16 2024;31(3):574–582. doi:10.1093/jamia/ocad241

16. Ahmad FS, Ricket IM, Hammill BG, et al. Computable phenotype implementation for a national, multicenter pragmatic clinical trial: Lessons learned from ADAPTABLE. Circ Cardiovasc Qual Outcomes. Jun 2020;13(6):e006292. doi:10.1161/circoutcomes.119.006292

17. Carrell DS, Floyd JS, Gruber S, et al. A general framework for developing computable clinical phenotype algorithms. J Am Med Inform Assoc. Aug 1 2024;31(8):1785–1796. doi:10.1093/jamia/ocae121

18. Food and Drug Administration. Search for FDA guidance documents. February 6, 2025. https://www.fda.gov/regulatory-information/search-fda-guidance-documents

19. Kim SY, Solomon DH, Liu J, Chang CL, Daniel GW, Schneeweiss S. Accuracy of identifying neutropenia diagnoses in outpatient claims data. Pharmacoepidemiol Drug Saf. Jul 2011;20(7):709–13. doi:10.1002/pds.2157

20. Knerr S, Hu EY, Zeliadt SB. Incidence of neutropenia in veterans receiving lung cancer chemotherapy: A comparison of administrative coding and electronic laboratory data. EGEMS (Wash DC). 2017;5(1):1269. doi:10.13063/2327-9214.1269

21. Spangler S, Gentlesk PJ. Acute pericarditis workup. February 7, 2025. https://emedicine.medscape.com/article/156951-workup?form=fpf

22. Silvert E, Hester L, Ramudu E, et al. Identifying signs and symptoms of AL amyloidosis in electronic health records using natural language processing, diagnosis codes, and manually abstracted registry data. Am J Hematol. Sep 2023;98(9):E255–e258. doi:10.1002/ajh.27019

23. Nath C, Albaghdadi MS, Jonnalagadda SR. A natural language processing tool for large-scale data extraction from echocardiography reports. PLoS One. 2016;11(4):e0153749. doi:10.1371/journal.pone.0153749

24. Jiang R, Gulbranson S, Bhattacharya M, et al. Incidence of pre-treatment and off-treatment pericardial and pleural effusions by line of therapy in patients with small cell lung cancer: An analysis of electronic health records data. J Clin Oncol. 2018;36(15_suppl):e20569-e20569. doi:10.1200/JCO.2018.36.15_suppl.e20569

25. Fontana RJ, Liou I, Reuben A, Suzuki A, Fiel MI, Lee W, Navarro V. AASLD practice guidance on drug, herbal, and dietary supplement-induced liver injury. Hepatology. Mar 1 2023;77(3):1036–1065. doi:10.1002/hep.32689

26. Chalasani NP, Maddur H, Russo MW, Wong RJ, Reddy KR. ACG clinical guideline: Diagnosis and management of idiosyncratic drug-induced liver injury. The American journal of gastroenterology. May 1 2021;116(5):878–898. doi:10.14309/ajg.0000000000001259

27. Weng C, Overby C. Drug induced liver injury. PheKB. Accessed February 7, 2025. https://phekb.org/phenotype/135

28. Forns J, Cainzos-Achirica M, Hellfritzsch M, et al. Validity of ICD-9 and ICD-10 codes used to identify acute liver injury: A study in three European data sources. Pharmacoepidemiol Drug Saf. Jul 2019;28(7):965–975. doi:10.1002/pds.4803

29. Giunta DH, Karlsson P, Younus M, Berglind IA, Kieler H, Reutfors J. Validation of diagnoses of liver disorders in users of systemic azole antifungal medication in Sweden. BMC Gastroenterol. Jan 5 2024;24(1):21. doi:10.1186/s12876-023-03110-w

30. Timmer A, de Sordi D, Kappen S, et al. Validity of hospital ICD-10-GM codes to identify acute liver injury in Germany. Pharmacoepidemiol Drug Saf. Oct 2019;28(10):1344–1352. doi:10.1002/pds.4855

31. Matheny ME, Yang J, Smith JC, et al. Enhancing Postmarketing Surveillance of Medical Products With Large Language Models. JAMA Netw Open. Aug 1 2024;7(8):e2428276. doi:10.1001/jamanetworkopen.2024.28276

32. Hripcsak G, Duke JD, Shah NH, et al. Observational Health Data Sciences and Informatics (OHDSI): Opportunities for Observational Researchers. (1879-8365 (Electronic))

33. Ostropolets A, Ryan P, Hripcsak G. Phenotyping in distributed data networks: selecting the right codes for the right patients. (1942-597X (Electronic))

34. Ostropolets A, Hripcsak G, Husain SA, Richter LR, Spotnitz M, Elhussein A, Ryan PB. Scalable and interpretable alternative to chart review for phenotype evaluation using standardized structured data from electronic health records. J Am Med Inform Assoc. Dec 22 2023;31(1):119–129. doi:10.1093/jamia/ocad202

35. Pacheco JA, Rasmussen LV, Wiley K, Jr., et al. Evaluation of the portability of computable phenotypes with natural language processing in the eMERGE network. Scientific reports. Feb 3 2023;13(1):1971. doi:10.1038/s41598-023-27481-y

36. Wong J, Prieto-Alhambra D, Rijnbeek PR, Desai RJ, Reps JM, Toh S. Applying Machine Learning in Distributed Data Networks for Pharmacoepidemiologic and Pharmacovigilance Studies: Opportunities, Challenges, and Considerations. Drug Saf. May 2022;45(5):493–510. doi:10.1007/s40264-022-01158-3

