## Appendices for "A Systematic Process for Assessing Fitness-for-Purpose of Health Outcomes for Computable Phenotyping with Electronic Health Record Data"

Sentinel

### Fitness-for-Purpose (FFP) Assessment Protocol

#### Introduction

Existing claims-based algorithms for certain health outcomes of interest (HOI) may be insufficiently accurate for research or surveillance. In such instances, it may be useful to develop a computable phenotype, i.e., a more sophisticated HOI algorithm that both leverages richer data in the electronic health record (EHR) (e.g., laboratory data or features from unstructured text in clinical notes) and uses more automated and thus reusable computational modeling methods (e.g., machine learning). This protocol defines the recommended steps to assess how feasible it would be to develop a computable phenotype for an HOI for which claims-based algorithms are insufficient and electronic health record data may be available, that is, a 'fitness-for-purpose' (FFP) assessment. It assumes that the 'Preliminary Steps Before FFP Assessment' document has been completed to ensure that the HOI under consideration is well-defined and doesn't represent a composite outcome. In addition, it is recommended that the team member(s) completing this activity review Carrell et al. (2024) for a clearer understanding of the objectives.

#### Methods

The following steps are intended to be completed by a single individual ('**team member**'), with consultation from both clinical and data experts to ensure full understanding of the approach, content, and application of the research. This consultation will be particularly important when evaluating clinical and data complexity.

While the steps are intended to be followed consecutively, performing an FFP is an iterative process in which later steps may feed back and inform additional work on prior steps.

While the focus of this protocol is on a series of steps to perform an FFP assessment, it additionally provides guidance on completing the 'FFP HOI Template' – a template provided for writing up the findings of the assessment. The structure of this report is discussed in the Appendix.

*At this point, the team member is ready to make a copy of the template, fill out the first page, and replace highlighted mentions of '**HOI**' with the name of the HOI being assessed.*

##### Step 1. Orientation to the HOI

An understanding of the HOI is important for completing the FFP assessment. The team member must become acquainted with the HOI and understand what it entails, the terminology commonly used, and how it is diagnosed in different clinical settings. This research will provide a foundation for both assessing the clinical complexity and understanding which EHR data may be relevant.

If the study team member is not a clinician and/or unfamiliar with the HOI subject area, using a search engine to find reputable patient-facing medical websites, such as the Mayo Clinic or Cleveland Clinic, can provide a helpful orientation to the HOI. Then, once the team member has a basic familiarity with the HOI, good resources for exploring the HOI in more depth include:

- UpToDate,
- Merative Micromedex,
- PubMed,
- ClinicalKey,
- the CDC website,
- the NIH website, and
- consultation with health professionals

In addition, the team member should identify any available FDA clinical guidance[1] as well as all relevant US-based medical society guidelines (which may include information about standardized diagnostic criteria). These can be found linked within UpToDate or in literature searches on PubMed, Google Scholar, or generic search engine results. The goal is to understand the 'gold standard' diagnostic criteria by which clinicians diagnose the condition.

*The information learned in this process should be succinctly summarized in the 'Orientation to HOI' section. The focus of this section is to provide a concise description of the HOI and present the diagnostic criteria. Supporting tables and figures (such as lists of ICD diagnosis codes) can be included as an appendix.*

At this stage, the team member will have a deeper familiarity with the HOI and is well prepared to again assess whether or not the current HOI is still both well-defined and not a composite of more than one HOI. For example, the HOI of 'major bleeding' may at first appear to not be a composite condition. Following the 'orientation to the HOI' step, it will become clear that 'major bleeding' is actually composite, consisting of at least six separate HOIs: intracranial hemorrhage, intraarticular hemorrhage, pericardial hemorrhage, hemorrhage requiring transfusion, retroperitoneal hemorrhage, and fatal hemorrhage. As described in the 'Preliminary Steps Before FFP Assessment' document, each of these must be evaluated separately.

#### Step 2. Literature Review of Previous Validation Studies/Algorithms

After orienting to the HOI, the team member should investigate previous work focused on methods to identify the HOI from electronic health data. The team member should first identify any validation studies using administrative claims data (typically focusing on ICD diagnostic codes), and then explore any natural language processing (NLP) studies available. This process will help to determine whether computable phenotyping is needed and inform the data complexity. If, for example, a validation study has been completed using ICD-10 codes and shows acceptable performance then this would suggest that a computable phenotype is not needed. In contrast, finding several ICD-10 validation studies showing poor or inconsistent performance and studies describing complex implementations of NLP algorithms would suggest that a computable phenotype with high data complexity is required.

While the team's interest in developing a computable phenotype may be to target a specific population (e.g., pediatrics), it is important to review work across multiple populations if not the general population. Often, it is difficult to find algorithms or

[1] https://www.fda.gov/regulatory-information/search-fda-guidance-documents

validation studies that are limited to specialized populations (such as pediatrics or immunosuppressed patients). Gauging performance in the general population, even if it is not the final population of interest, can still be helpful for informing the level of data complexity.

Searches on PubMed, Google Scholar, and any general-purpose search engine can help locate relevant publications. Some useful search templates include:

- HOI + 'icd' + 'validation'
- HOI + 'diagnosis' + 'validation'
- HOI + 'algorithm'
- HOI + 'agreement'
- HOI + 'icd' + 'validity'
- HOI + 'diagnosis' + 'validity'
- HOI + 'nlp'/ 'natural language processing'

In general, avoid papers that deal with clinical decision support and prediction/scoring tools. These tend not to focus on validating approaches to identify an HOI from administrative data.

The introductory sections of most of these papers should help elucidate earlier studies which the current work is building on. This is especially true with NLP studies which are often undertaken when algorithms using structured administrative data (such as ICD diagnosis codes) have poor performance. It may be useful to obtain and review the earlier studies cited in these introductory sections.

*The most relevant studies should be written up briefly in the 'Literature Review of Previous Validation Studies/Algorithms' section. Relevance is defined by recency of the data being used (not necessarily the publication date), locality (e.g., US studies preferred over studies from other regions), and latest codes (e.g., preference for ICD-10 over ICD-9). The populations should be succinctly described along with the gold standard used and performance metrics (particularly, sensitivity and positive predictive value). Briefly describe the applicability of each algorithm to the HOI and population or question of interest.*

#### Step 3. Determine Relevant Data Elements

The team member will identify the specific types of data needed for the clinical diagnostic criteria (e.g., laboratory tests, radiology reports, etc.) and/or claims-based algorithms in the EHR, and determine where these data are located within the EHR (i.e., structured data – such as ICD codes or laboratory test results; unstructured – such as clinical notes). Consideration will be given to the availability of records within a health system (i.e., whether the data elements are likely to be found in inpatient vs. outpatient records). EHR data may also be required to rule out other etiologies for the HOI, apart from medications. For instance, when considering a computable phenotype for Drug Induced Liver Injury, it is necessary to consider how to exclude other causes of liver injury, such as infections, cancer and other causes.

*All data elements are summarized in a table with the header 'Data Elements' based on whether they are 'necessary' or could be 'useful' in identifying the HOI.*

*Elements are 'necessary' if the HOI could not be completed without them. Elements are 'useful' if they, even while not strictly required, would prove helpful in providing additional context or information. For example, information about the HOI pericardial effusion would be gleaned from echocardiograms ('necessary') but might also be available in other radiology reports like CTs or MRIs ('useful'). 'Useful' may be left blank.*

#### Step 4. Assess Clinical Complexity

Once the relevant information has been collected from the literature, the FFP Checklist will be used to develop a rating for clinical complexity of LOW, MEDIUM, or HIGH.

When assessing clinical complexity, it is important to not place too much emphasis on the potential for undetected or asymptomatic cases, except in the rare case where clinically relevant cases are deemed to be missing. No computable phenotype relying on EHR data will be able to identify cases that did not come to medical attention, due to the inherent nature of the data (generated from medical encounters). Thus, it will be necessary to accept the limitation that some undetected or asymptomatic cases will be missed. For example, neutropenia is diagnosed by quantifying neutrophils in the patient's blood. Patients who have neutropenia (i.e., a neutrophil count below some threshold) but are asymptomatic are unlikely to be diagnosed and are thus out of reach of both diagnosis codes and a computable phenotype. Therefore, it is still reasonable to assign neutropenia's clinical complexity as LOW since neutrophil counts unambiguously determine neutropenia.

In certain rare clinically relevant cases, missingness of key data may be important to consider. For example, encapsulated bacterial infections are diagnosed by the isolation of the bacterial species from a body fluid (e.g., sputum, blood, etc.), thereby confirming the etiology of the infection. Individuals with severe illness who are hospitalized are likely to have these tests conducted, but the diagnostic yield of these tests is low. This severely limits the ability to identify the HOI from EHR data even for individuals with a clinically significant disease. Thus, in assessing the clinical complexity for encapsulated bacterial infections, the low likelihood of being able to identify an etiologic pathogen from the specimen results in HIGH clinical complexity.

Even with the protocol steps completed so far, it may prove difficult to come to a definitive assessment of clinical complexity. The team member may need to consult with clinicians to better understand nuances of how a diagnosis is made in practice. In addition, a literature review for studies documenting the level of agreement between clinicians will prove valuable. If there is high inter-rater agreement among clinicians provided with the same information, the clinical complexity will likely be LOW, whereas low agreement suggests MEDIUM or HARD clinical complexity.

The FFP Checklist is meant to ensure that all relevant aspects have been considered in formulating an assessment of complexity level. It is not meant to be used as a score sheet, and there is no simple heuristic that translates the number or ratio of items marked as 'more' or 'less' difficult directly to the final assessment. That is, we do not recommend attempting to specify a cut point or algorithm for each level of complexity. A larger number of items marked 'more difficult' may indicate a higher level of clinical complexity. On the other hand, an HOI may have many items marked as “easier” and only 1-2 marked as ‘more difficult’, yet ultimately be deemed HARD. This requires nuanced interpretation of the worksheet and experience with making these determinations. It may be helpful to review the examples provided for each level of complexity on the introductory page of the FFP checklist.

*The final determination (LOW/MEDIUM/HIGH) should be written clearly at the top of the ‘Clinical Complexity’ section followed by a single sentence summary of the rationale. In the following paragraph(s), the detailed reasoning for the assessed rating should be presented.*

#### Step 5. Assess Data Complexity

Once the relevant information has been collected from the literature, use the FFP Checklist to develop a rating for data complexity of LOW, MEDIUM, or HIGH. In general, higher complexity is associated with the following characteristics: increasing numbers of data points needed, wider range of types of data needed, and any use of unstructured data (NLP).

Even with the protocol steps completed so far, it may prove difficult to come to a definitive conclusion on the level of data complexity. The team member may need to consult with data experts knowledgeable about how structured and unstructured administrative data are typically captured to guide their decision. Questions might include how available or easy-to-parse lab values are, or how complex a high-performing NLP algorithm is.

Simple NLP algorithms identify keywords (e.g., ‘regular expressions’) in clinical text and may include straightforward rules (e.g., the outcome is determined to be present if a note contains a particular keyword and does not contain another). Additional complexity is added when the NLP requires contextual information (e.g., identification of ‘negation’), imposes weights, or relies on structural cues (e.g., locating section headings, ‘sentence segmentation’). Complex algorithms are described with language like ‘deep learning’, ‘transformers’, or the names of various machine learning algorithms. While these may perform better at a single site, they can be very difficult to implement, time-consuming, computationally expensive to run, and have unclear performance when used across multiple sites. If NLP is required, this suggests a data complexity of MEDIUM (for simple algorithms) or HARD (for complex algorithms).

Again, as discussed in Step 5, the FFP Checklist is meant to provide guidance when determining the level of data complexity, ensuring all relevant considerations are brought to light, but there is no algorithm or cut point that can be used to directly translate the findings on the worksheet into a specific level of complexity. This process requires nuanced interpretation of the worksheet and experience with making these determinations. It may be helpful to review the examples provided for each level of complexity on the introductory page of the FFP checklist.

*The final determination (LOW/MEDIUM/HIGH) should be written clearly at the top of the ‘Data Complexity’ section followed by a single sentence summary of the rationale. In the following paragraph(s), the detailed reasoning for the assessed rating should be presented.*

#### Step 6. Write Conclusion/Fitness for Purpose Recommendation

The final section of the 'Detailed Discussion', the 'Conclusion/Fitness for Purpose Recommendation' requires a summary of the clinical and data complexity, as well as an evaluation of the overall assessment as EASY, MODERATE, or HARD. This conclusion must present a case for the overall assessment weighing both the clinical and data complexity. There is no simple formula for integrating the two determinations. This decision is easiest if the clinical and data complexity are similar (e.g., if both are LOW then the HOI can be assessed as EASY; if both are HIGH, it can be assessed as HARD). The challenge arises when the findings for clinical and data complexity are discordant. In general, we expect that the highest level of difficulty will disproportionately affect the determination. For example, if clinical complexity is LOW but data complexity is HIGH, the HOI will likely be deemed HARD. HOI-specific considerations may affect the final determination.

Next, given the information presented so far in the document, it is important to highlight what the next steps should be. For example, these might include recommending an ICD code validation study (e.g., if there are promising validation studies using ICD-9 codes but no studies yet of ICD-10 codes), validating an NLP algorithm, exploring the availability/missingness of particular laboratory results, or evaluating the interpretability of key laboratory results.

*The overall assessment for HOI difficulty (EASY/MODERATE/HARD) should be presented in the 'Conclusion/Fitness-for-Purpose Recommendation' section, including how the data and clinical complexity are reconciled. A recommendation for next steps should be given.*

#### Step 7. References and Appendices

A bibliography of the sources used, particularly for determining clinical and data complexity, should be provided under the 'References' heading. This will not only document the sources underlying the current work but provide a starting point for future work including the development of a computable phenotype.

Appendices can be added to list and/or summarize clinical guidelines for the HOI, list ICD diagnostic codes, or other useful information. These are intended to be illustrative rather than exhaustive.

#### Step 8. Compose Executive Summary

The executive summary is a 1-page summary of the elements in the 'Detailed Description' section completed so far. Each of these sections provides a brief summary of the content provided in the 'Detailed Description' to make the content more easily digestible, and with the understanding that a more complete discussion is available in the 'Detailed Description'.

Sections:

- **Overall Assessment**: Clearly state the overall FFP assessment as EASY/MODERATE/HARD, followed by a 1 sentence summary of the requirements (e.g., 'A computable algorithm would require laboratory results…').

- **Orientation to HOI**: A short description of the HOI and diagnostic criteria.
- **Existing Computable Algorithms**: A brief summary of the findings from the most salient computable algorithms/validation studies.
- **Clinical Complexity**: Provide the assessed clinical complexity (LOW/MEDIUM/HIGH) with a brief explanation of the reasons for the determination.
- **Data Complexity**: Provide the assessed data complexity (LOW/MEDIUM/HIGH) with a brief explanation of the reasons for the determination.
- **Conclusion / Fitness-for-Purpose Recommendation**: This section should state the conclusion of the report along with any recommendations for next steps.

**Limitations**: A brief statement about any limitations relevant to the FFP assessment. Typically, this will include the date the literature review was conducted and a statement about what level of positive predictive value and/or sensitivity were considered acceptable.

#### References

Carrell, D. S. *et al.* (2024) 'A general framework for developing computable clinical phenotype algorithms', *Journal of the American Medical Informatics Association: JAMIA*. Oxford University Press (OUP), 31(8), pp. 1785–1796.

### Appendix 1: Fitness-for-Purpose (FFP) HOI Template

While the focus of this protocol is on a series of steps to perform an FFP assessment, it additionally provides guidance on completing the 'FFP HOI Template' – a template for writing up the findings of the assessment. The template is composed of two primary sections:

- The **Executive Summary** provides a 1-page overview of the FFP assessment, providing a succinct overview of the key issues.
- The **Detailed Description** explores in greater detail the relevant studies and reasons for clinical, data, and overall complexity decisions.

The **Executive Summary** consists of the following subsections:

- The **Overall Assessment** includes the overall difficulty rating for the FFP (*easy*, *moderate,* or *hard*) with a short explanation.
- The **Orientation to HOI** briefly orients the reader to the HOI.
- The **Existing Computable Algorithms** introduces the most important papers relating to past attempts to identify or validate this HOI using computable algorithms (including algorithms based on ICD diagnosis codes).
- **Clinical Complexity** reveals the rating for the level of clinical complexity (*low*, *medium*, or *high*) along with a brief explanation.
- **Data Complexity** reveals the rating for the level of data complexity (*low*, *medium*, or *high*) along with a brief explanation.
- The **Conclusion / Fitness-for-Purpose Recommendation** provides the most important takeaways from the assessment as well as any recommendations for how to proceed.
- **Limitations** summarizes key limitations in the performance of the assessment, and captures assumptions made.

The **Detailed Description** has the following structure:

- The **Orientation to HOI** orients the reader to the HOI and includes any relevant diagnostic criteria.
- The **Literature Review of Previous Validation Studies/Algorithms** orients the reader to relevant validation studies. In a well-studied HOI, this will likely only include the more salient studies.
- **Data Elements** breaks down the EHR data which would be either necessary (i.e., the computable phenotype could not be completed without this resource) or useful (i.e., this resource could be useful in developing a computable phenotype). This may include particular lab tests, medications, or particular types of clinical notes.
- **Clinical Complexity** discusses in detail the reason(s) for assigning the selected level of complexity to the HOI.

- **Data Complexity** discusses in detail the reason(s) for assigning the selected level of complexity to the HOI.
- **Conclusion / Fitness-for-Purpose Recommendation** discusses how the clinical and data complexity decisions inform the overall assessed difficulty for a computable phenotype. In addition, where appropriate, a recommendation is made for how to proceed if this HOI were selected for implementation/development.

### Preliminary Steps Before Computable Phenotyping Fitness-for-Purpose (FFP) Assessment: Ensuring the Health Outcome of Interest (HOI) is Ready for Evaluation

#### 1. Is the Health Outcome of Interest Well-Defined?

An HOI must be well-defined before it can be put through a computable phenotyping fitness-for-purpose (FFP) assessment. This means it must represent a specific medical condition or conditions that can be diagnosed in a conventional healthcare setting and that there is general understanding among health professionals about what is meant by the condition(s) included in the HOI.

An example of a well-defined HOI is neutropenia, which refers to a deficiency in the number of neutrophils, a type of white blood cell. Its meaning is clear since it refers to a specific medical condition. In contrast, the HOI "hematologic abnormalities" is not well-defined. The term "hematologic" is very broad and could refer to different components of the blood, including cells or the coagulation system. It could refer to diverse types of abnormalities, for instance cell counts being too high or too low. It is not possible to conduct an FFP assessment of hematologic abnormalities without additional work to define the specific abnormalities of concern.

Additional examples come from infectious diseases. Meningitis is an example of a well-defined HOI since it represents infection in a specific location in the body. In contrast, the HOI "serious infections" is not well defined. There is no single, agreed-upon definition. There is no agreement on which infections should be included or how many. The word "serious" can have varying meanings. Different existing algorithms for serious infections include different types and numbers of infections, showing that this term may have different meanings in different settings or contexts (Barber 2013).

The first step in approaching an HOI that is not well-defined is to seek more clarification. This may involve discussing it with the responsible division within the Office of New Drugs and asking them to provide a more specific description of the HOI. It may help to outline a set of questions that need to be answered to move forward and to seek input from groups with a stake in the investigation. For example, questions for the HOI "serious infections" could include: Which infections should be included? How many different infections should be included? What types of infections are most important? What is meant by "serious"? Narrowing down the HOI to a specific, well-defined entity will make it possible to proceed with the FFP assessment.

#### 2. Is the Health Outcome of Interest a Composite Outcome?

Some HOIs represent a single clinical condition, while others represent a group of conditions or events, that is, a composite outcome. For instance, myocardial infarction (MI) represents a single clinical condition, while Major Adverse Cardiovascular Events (MACE) is a composite outcome consisting of MI, stroke, cardiovascular death, and sometimes other components. The conditions which make up a composite HOI must be enumerated and considered individually before the HOI can be put through a computable phenotyping FFP assessment.

In most cases, it will not be possible to conduct a single FFP assessment on a composite outcome as a whole because results of the assessment may differ for the different subcomponents within the composite outcome. For example, individual outcomes may differ in their data complexity and clinical complexity. The answer to questions such as “What types of data are needed to define this outcome?” and “Is there broad agreement between society guidelines about how this outcome is defined?” may vary across the components of a composite outcome.

Before beginning the FFP process, you should break the composite down into its individual components (Figure) and also consider whether any of them are themselves composite outcomes that need to be further disaggregated. Once you have deconstructed the composite outcome to the level of individual outcomes, then each of these outcomes can go through the FFP process as its own entity. If there are many components, the FFP assessment may take more time and resources to conduct. You may wish to consider prioritizing the components and assessing a subgroup of them, such as those that are the most common or the most serious.

Once each component’s clinical and data complexity have been assessed, you are ready to consider them as a whole and determine the overall level of complexity and feasibility for the composite. That step of the process will be addressed in a separate document.

It is also possible for a composite outcome to be ill-defined and require further clarification. For instance, “serious infections” is an HOI that is both composite and not well-defined. Before you can break it down into its component parts, you will first need to determine what infections (components) should be included. Once you have broken down a composite outcome into its components, you will need to make sure each individual component is well-defined. If they are not, you should return to the first process described above to clarify that outcome further and then return to the process of assessing the composite outcome.

#### References

Barber, C., Lacaille, D. and Fortin, P. R. (2013) Systematic review of validation studies of the use of administrative data to identify serious infections: Administrative data to identify infections, *Arthritis care & research*. Wiley, 65(8), pp. 1343–1357.

### Figure: Process for Assessing Feasibility of a Composite Outcome

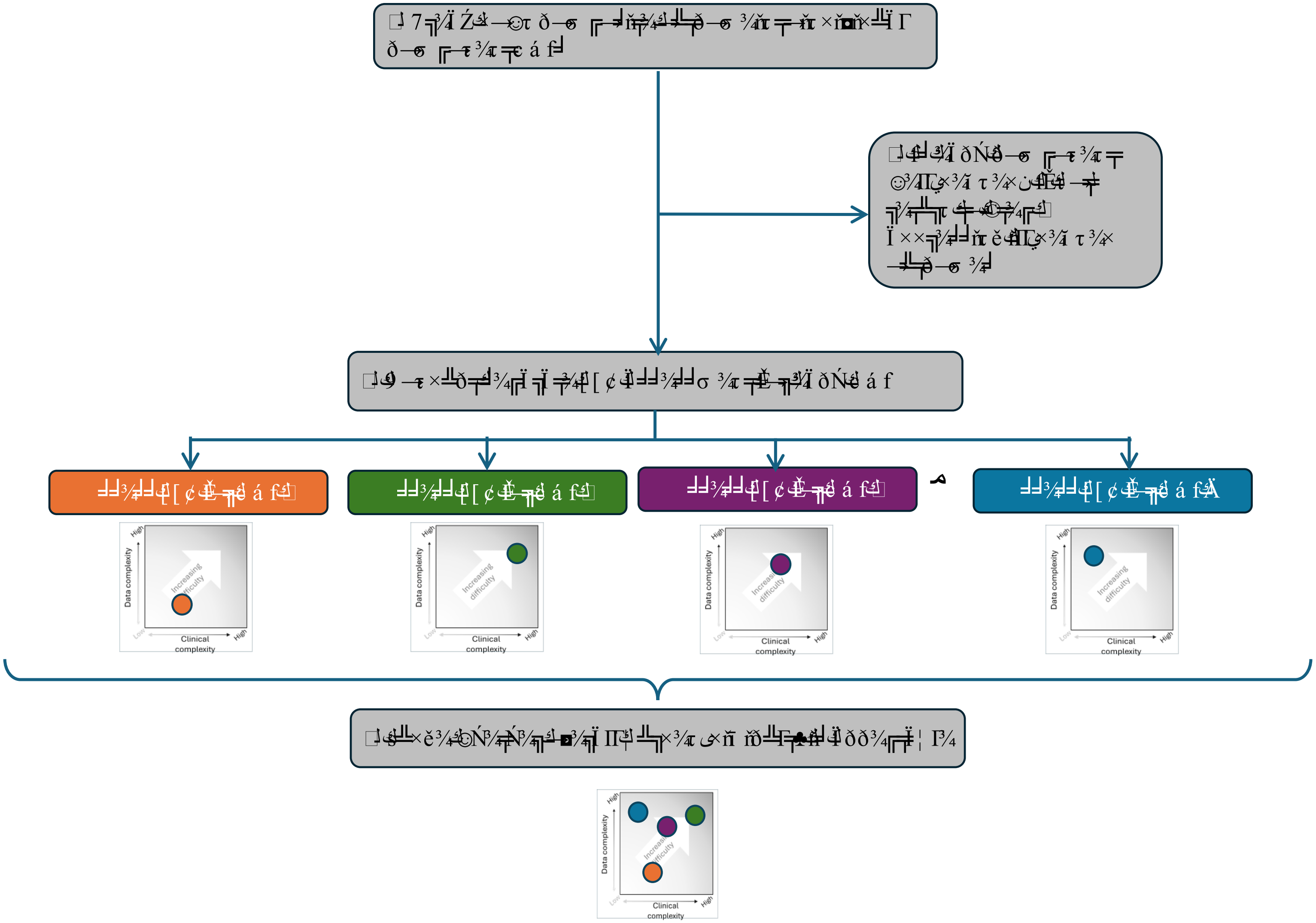

### Appendix 3. Select Publicaly-Available FDA ARIA Insufficiency Memos for Health Outcomes of Interest (HOI) Included in Fitness-for-Purpose (FFP) Assessment

| Health Outcomes of Interest (HOI) | FDA ARIA Insufficiency Memos (link) |
|---|---|
| Neutropenia | https://www.sentinelinitiative.org/studies/drugs/assessing-arias-ability-evaluate-safety-concern/siliq-brodalumab |
| Pericardial Effusion | https://www.sentinelinitiative.org/studies/drugs/assessing-arias-ability-evaluate-safety-concern/omegaven-fish-oil-triglycerides |
| Drug-Induced Liver Injury (DILI) | https://www.sentinelinitiative.org/studies/drugs/assessing-arias-ability-evaluate-safety-concern/jynarque-tolvaptan |
| Serious Infections | https://www.sentinelinitiative.org/studies/drugs/assessing-arias-ability-evaluate-safety-concern/enjaymo-sutimlimab-jome |
| Major Bleed | https://www.sentinelinitiative.org/studies/drugs/assessing-arias-ability-evaluate-safety-concern/pradaxa-dabigatran-etexilate |
| Hemolytic Anemia | https://www.sentinelinitiative.org/studies/drugs/assessing-arias-ability-evaluate-safety-concern/arakoda-tafenoquine |
| Encapsulated Bacterial Infections | https://www.sentinelinitiative.org/studies/drugs/assessing-arias-ability-evaluate-safety-concern/empaveli-pegcetacoplan |
| Pulmonary Arterial Hypertension | https://www.sentinelinitiative.org/studies/drugs/assessing-arias-ability-evaluate-safety-concern/fintepla-fenfluramine |

Sentinel

#### Data Complexity

*Data complexity refers to how difficult it is to accurately identify the Health Outcome of Interest (HOI) using retrospective health data, including elements from insurance claims and the electronic medical record. Domains of data complexity include data availability, data structure and ease of extraction, dimensionality (number of data points or data types required), and ease of interpretation.*

**Descriptions of classifications of data complexity:**

**High:** Complex integration of structured and unstructured data elements. Features based on unstructured data may require application of Natural Language Processing (NLP) to multiple concepts or interpretation of images. Statistical modeling of information from dozens or hundreds of features likely needed to generate adequate performance. There may be little or no prior research to guide these efforts, which may require extensive pilot and development work. Examples:

- Anaphylaxis (requires data about numerous signs and symptoms in structured and unstructured format; high dimensionality; incomplete documentation of important information, e.g., timing of events)
- Drug-induced liver injury (lack of clinical consensus on diagnostic criteria; phenotype based on numerous data types, some unstructured; requires repeated measures over time; need to rule out many other possible causes)

**Medium:** Requires complex structured data or simple application of NLP on semi-structured data (e.g., imaging reports). Some pilot or validation work likely required to develop computable phenotyping algorithm. Examples:

- Pericardial effusion
- Hepatitis B reactivation

**Low:** Data required are widely available, such as laboratory tests in structured format or combinations of diagnosis codes. Few barriers to implementation, and little or no pilot or validation work required. Examples:

- Acute kidney injury (defined from baseline and follow-up tests for creatinine)
- Neutropenia (defined from absolute neutrophil count)

<u>Questions to consider before completing the worksheet:</u>

- What data elements are needed (e.g., laboratory results, imaging, vital signs, symptoms, physical exam findings)?
- Are the data elements recorded as structured or semi-structured data or in free text?
- Is the interpretation of laboratory test results difficult?
- Are separate algorithms required for multiple elements that together make up a composite outcome (e.g., for serious infection, components may include pneumonia, skin or soft tissue infection, bacteremia, and others)?
- Are statistical models that integrate information from numerous features (constructed variables) required to identify the outcome with adequate accuracy (e.g., anaphylaxis)?

Sentinel

#### Worksheet for Determining Level of Data Complexity

| Question or Characteristic | Easier | More Difficult |
|---|---|---|
| What type(s) of data elements are needed? | | |
| Laboratory test: | | |
| routine, results in structured form (e.g., creatinine, for acute kidney injury) | ☐ Yes | |
| less common (may be "send out" or stored as PDF) | | ☐ Yes |
| results may be free text or unstructured | | ☐ Yes |
| Imaging/procedure result: | | |
| more common finding, standardized language (e.g., pericardial effusion) | ☐ Yes | |
| less common finding, more variable language | | ☐ Yes |
| Clinical free text: | | |
| unstructured, signs or symptoms found in free text, (e.g., self-report of symptoms in a clinical note) | | ☐ Yes |
| How many different data elements are required to determine the diagnosis? | | |
| 1-2 | ☐ Yes | |
| 3+ | | ☐ Yes |
| Is natural language processing required? | ☐ No | ☐ Yes |
| Timing and number of laboratory or imaging results: | | |
| Does making the diagnosis require values from repeated laboratory or imaging results over time? | | ☐ Yes |
| Does the diagnosis rely on integrating results from several laboratory or imaging tests? (e.g., Hepatitis B reactivation) | | ☐ Yes |
| Does the diagnosis require comparing current lab or imaging results to a baseline? | | ☐ Yes |
| *If* "Yes": is it likely the baseline lab or imaging results may be found in a different healthcare system? | | ☐ Yes |
| Are the desired data elements likely to be found in an image (e.g., a scanned PDF) rather than in structured data elements? (e.g., send-out labs, pulmonary function tests, EKGs) | | ☐ Yes |
| Are multiple algorithms required to identify a composite outcome? (e.g., serious infection) | | ☐ Yes |
| Is the interpretation of laboratory test results complex (e.g., hepatitis B reactivation)? | | ☐ Yes |
| For laboratory results: is this a commonly ordered lab that is expected to be available for most exposed patients? | ☐ Yes | |
| Are statistical models that integrate information from numerous features (e.g., sign, symptom, lab result) required to identify the outcome with adequate accuracy (e.g., anaphylaxis)? | | ☐ Yes |

### Clinical Complexity

*Clinical complexity can be defined as how difficult it is for a provider to accurately diagnose the condition. Some of the contributing factors include whether evidence from multiple or different types of test or imaging results is needed to make a diagnosis, whether there is a definitive diagnostic test(s) or the diagnosis relies on subjective symptoms, and whether there is consensus about diagnostic criteria.*

**Descriptions of classifications of clinical complexity:**

**High:** Clinical diagnosis difficult. No gold standard diagnostic test. Diagnosis based on physician assessment and synthesis of signs and symptoms (e.g., heart failure, anaphylaxis). Requires discriminating the diagnosis of interest from competing diagnoses (e.g., drug-induced liver injury [DILI]). Disagreement may be common even between two expert physicians. Consensus clinical guidelines for diagnosis may not exist. Examples:

- Anaphylaxis
- DILI

**Medium:** Diagnosis may require synthesis of multiple data sources and consideration of one or two well-defined competing diagnoses. May require evaluation of changes in multiple laboratory tests over time (e.g., Hepatitis B reactivation). Significant challenges in availability and/or diagnostic yield of key laboratory tests. Examples:

- Hepatitis B reactivation
- Encapsulated bacterial infections

**Low:** Diagnosis based on a simple diagnostic test. Little or no uncertainty in diagnosis or disagreement between physicians. Examples:

- Acute kidney injury (diagnosis based on change in a single widely-used laboratory test, creatinine)
- Neutropenia (diagnosis based on a single laboratory test [absolute neutrophil count])

**Questions to consider before completing the worksheet:**

- Are there consensus clinical guidelines for diagnosis?
- Is there a gold standard diagnostic test, and is the interpretation of results unambiguous?
- When different physicians receive the same information, is agreement between them expected to be high?
- Are there competing diagnoses that are particularly challenging for the provider to distinguish between?

### Worksheet for Determining Level of Clinical Complexity

| Question or Characteristic | Easier | More Difficult |
|---|---|---|
| Medical specialty society or other clinical society guidelines | | |
| One or a few guidelines, clear, generally in agreement | ☐ Yes | |
| No guideline available[1] | | ☐ Yes |
| Multiple guidelines, conflicting or inconsistent | | ☐ Yes |
| Diagnosis is based on: | | |
| Objective measures or tests (e.g., lab results or imaging) | ☐ Yes | |
| Subjective findings (symptoms, physical exam findings) | | ☐ Yes |
| Diagnosis is based on 1 or 2 "gold standard" test results that are straightforward to interpret | ☐ Yes | |
| "Gold standard" is problematic or difficult to interpret; for instance, results may be ambiguous or the sensitivity of the test may be very low, leading to uncertainty | | ☐ Yes |
| Diagnosis requires considerable clinical judgement and/or interpretation of multiple pieces of data | | ☐ Yes |
| If different clinicians were given identical information, the agreement between them is likely to be: | | |
| High | ☐ Yes | |
| Low | | ☐ Yes |
| Does making the diagnosis require data from multiple points in time, such as multiple encounters with the patient or longitudinal lab results? | | ☐ Yes |
| Does making the diagnosis require integrating data from many different tests or sources? E.g., results from multiple lab tests, or different types of data (lab results, imaging, signs, symptoms) | | ☐ Yes |
| Can the diagnosis be made using 1-2 relatively common lab results that are available in structured or numeric form with clear, consistent cut off values? (e.g., neutropenia) | ☐ Yes | |
| Is it necessary to exclude competing diagnoses? (e.g., DILI) | | ☐ Yes |
| Does making the diagnosis require identifying and defining a baseline state and then a subsequent state? (e.g., Hepatitis B reactivation) | | ☐ Yes |

[1]The lack of a society guideline typically means greater clinical complexity because it indicates less certainty about how the HOI is diagnosed; however, in rare cases, the lack of a society guideline is due to a diagnosis being extremely straightforward.

#### Appendix: Definitions of Key Terms

| Term | Definition |
|---|---|
| Data structure | A data structure refers to how information is stored, organized, and managed within the EHR or administrative claims. It includes the layout and format of data, impacting how easily information can be accessed, retrieved, and analyzed. For an FFP assessment, the focus is on the difficulty of retrieving data from a particular structure. For example, structured data (e.g., diagnosis codes) is designed for easier retrieval whereas unstructured data (i.e., clinical free text) is not optimized. |
| Structured data | Data that are organized in a predefined, systematic way, making them easy to access and interpret. For example, laboratory results can be stored in a database table where one column contains the type of laboratory test and another column contains the corresponding result value. This format allows for straightforward data retrieval and analysis. |
| Semi-structured data | Data that are not entirely unstructured but lacks a rigid, predefined format. They are often stored in free text templates, such as questionnaires or procedure reports. For example, a procedure report may appear as free text but is organized in a structured manner, with the physician "filling in the blanks." Extracting information from semi-structured data typically involves using simple NLP techniques to identify specific template locations and retrieve the corresponding values. |
| Unstructured data | Data that are not organized in a predefined manner, lacking clear divisions or specific areas for locating desired elements. For example, progress notes in medical records often start with a free text description of a patient's appearance, symptoms, and concerns. To extract meaningful information from such data, NLP algorithms are typically required to search through the entire text. |
| Features | Features (analogous to independent variables in statistical analysis) are variables constructed from healthcare data, such as diagnosis codes, demographic information, clinical measurements, or unstructured data (e.g., counts of terms in clinical notes). These features serve as inputs for machine learning models designed to identify specific phenotypes. For example, when developing a computable phenotype for anaphylaxis, features could include diagnosis codes for anaphylaxis and allergic reactions as well as counts of terms like “epinephrine” in clinical notes. The statistical model will assign importance weights to each feature and use these weights to predict whether an individual has anaphylaxis or not. |
| “Send out” laboratory tests | “Send out” labs are tests that a primary laboratory or healthcare system sends out to a more specialized lab or “reference laboratory” to perform due to a lack of necessary equipment or expertise. These tests are typically more specialized tests that are rarely performed or require unusual or specialized equipment or reagents. Examples include genetic testing, certain cancer-related tests, and specialized infectious disease tests. Results are then returned to the original lab/healthcare system but may be returned in a format that is not easily integrated into the EHR. For instance, results may be faxed or scanned into the outpatient EHR and may have inconsistent file names. |
| Dimensionality | Dimensionality refers to the number of features or attributes used as input for a computable phenotype algorithm, particular in the context of statistical modeling. High dimensionality refers to the situation where there are more features than observations, which causes numerous |

| | |
|---|---|
| | statistical challenges including overfitting, increased computational complexity, and reduced interpretability. |

### Fitness-for-Purpose (FFP) Assessment: HOI

| | |
|---|---|
| **Health Outcome of Interest:** | HOI |
| **Date:** | Month DD, YYYY |
| **Subject:** | Health Outcome of Interest Feasibility Assessment for HOI |

Sentinel

#### Executive Summary

**Overall Assessment:** *EASY/MODERATE/HARD* – brief summary

**Orientation to HOI:** TO DO

**Existing Computable Algorithms:** TO DO

**Clinical Complexity:** *LOW/MEDIUM/HIGH* – brief explanation

**Data Complexity:** *LOW/MEDIUM/HIGH* – brief explanation

**Conclusion / Fitness-for-Purpose Recommendation:** TO DO

**Limitations:** This fitness-for-purpose recommendation is based on literature reviews as of MONTH YEAR. We assume an ideal performance equates to a PPV of at least 80%.

### Detailed Discussion

#### Orientation to HOI

HOI is …

#### Literature Review of Previous Validation Studies / Algorithms:

- TO DO

#### Data Elements:

| Necessary EHR Data | Useful EHR Data |
|---|---|

**Clinical Complexity:** *LOW/MEDIUM/HIGH – brief why?*
TO DO

The FFP clinical complexity worksheet has been included in Appendix 3.

**Data Complexity:** *LOW/MEDIUM/HIGH – brief why?*
TO DO

The FFP data complexity worksheet has been included in Appendix 4.

#### Conclusion / Fitness-for-Purpose (FFP) Recommendation

TO DO

#### References

TO DO

#### Appendix 1: Society Guidelines for HOI

Focus on US society guidelines if available. If they are not available, expand to consider European or international guidelines.

#### Appendix 2: ICD Diagnostic Codes Relevant to HOI

This list of ICD-9 and -10 codes was compiled from XXX, with additional codes provided by XXX. It is not intended to be exhaustive, but provided for reference purposes.

ICD-9

| Code | Description | Source? |
| --- | --- | --- |

ICD-10

Code

| Code | Description | Source? |
| --- | --- | --- |

#### Appendix 3: Clinical Complexity Worksheet

| Question or Characteristic | Easier | More Difficult |
|---|---|---|
| Medical specialty society or other clinical society guidelines | | |
| One or a few guidelines, clear, generally in agreement | ☐ Yes | |
| No guideline available[1] | | ☐ Yes |
| Multiple guidelines, conflicting or inconsistent | | ☐ Yes |
| Diagnosis is based on: | | |
| Objective measures or tests (e.g., lab results or imaging) | ☐ Yes | |
| Subjective findings (symptoms, physical exam findings) | | ☐ Yes |
| Diagnosis is based on 1 or 2 "gold standard" test results that are straightforward to interpret | ☐ Yes | |
| "Gold standard" is problematic or difficult to interpret; for instance, results may be ambiguous or the sensitivity of the test may be very low, leading to uncertainty | | ☐ Yes |
| Diagnosis requires considerable clinical judgement and/or interpretation of multiple pieces of data | | ☐ Yes |
| If different clinicians were given identical information, the agreement between them is likely to be: | | |
| High | ☐ Yes | |
| Low | | ☐ Yes |
| Does making the diagnosis require data from multiple points in time, such as multiple encounters with the patient or longitudinal lab results? | | ☐ Yes |
| Does making the diagnosis require integrating data from many different tests or sources? E.g., results from multiple lab tests, or different types of data (lab results, imaging, signs, symptoms) | | ☐ Yes |
| Can the diagnosis be made using 1-2 relatively common lab results that are available in structured or numeric form with clear, consistent cut off values? (e.g., neutropenia) | ☐ Yes | |
| Is it necessary to exclude competing diagnoses? (e.g., DILI) | | ☐ Yes |
| Does making the diagnosis require identifying and defining a baseline state and then a subsequent state? (e.g., Hepatitis B reactivation) | | ☐ Yes |

[1]The lack of a society guideline typically means greater clinical complexity because it indicates less certainty about how the HOI is diagnosed; however, in rare cases, the lack of a society guideline is due to a diagnosis being extremely straightforward.

### Appendix 4: Data Complexity Worksheet

| Question or Characteristic | Easier | More Difficult |
|---|---|---|
| What type(s) of data elements are needed? | | |
| Laboratory test: |  |  |
| routine, results in structured form (e.g., creatinine, for acute kidney injury) | ☐ Yes |  |
| less common (may be "send out" or stored as PDF) |  | ☐ Yes |
| results may be free text or unstructured |  | ☐ Yes |
| Imaging/procedure result: |  |  |
| more common finding, standardized language (e.g., pericardial effusion) | ☐ Yes |  |
| less common finding, more variable language |  | ☐ Yes |
| Clinical free text: |  |  |
| unstructured, signs or symptoms found in free text, (e.g., self-report of symptoms in a clinical note) |  | ☐ Yes |
| How many different data elements are required to determine the diagnosis? | | |
| 1-2 | ☐ Yes |  |
| 3+ |  | ☐ Yes |
| Is natural language processing required? | ☐ No | ☐ Yes |
| Timing and number of laboratory or imaging results: | | |
| Does making the diagnosis require values from repeated laboratory or imaging results over time? |  | ☐ Yes |
| Does the diagnosis rely on integrating results from several laboratory or imaging tests? (e.g., Hepatitis B reactivation) |  | ☐ Yes |
| Does the diagnosis require comparing current lab or imaging results to a baseline? |  | ☐ Yes |
| *If "Yes"*: is it likely the baseline lab or imaging results may be found in a different healthcare system? |  | ☐ Yes |
| Are the desired data elements likely to be found in an image (e.g., a scanned PDF) rather than in structured data elements? (e.g., send-out labs, pulmonary function tests, EKGs) | | ☐ Yes |
| Are multiple algorithms required to identify a composite outcome? (e.g., serious infection) | | ☐ Yes |
| Is the interpretation of laboratory test results complex (e.g., hepatitis B reactivation)? | | ☐ Yes |
| For laboratory results: is this a commonly ordered lab that is expected to be available for most exposed patients? | ☐ Yes | |
| Are statistical models that integrate information from numerous features (e.g., sign, symptom, lab result) required to identify the outcome with adequate accuracy (e.g., anaphylaxis)? | | ☐ Yes |

### Composite Outcomes:
### Overall Assessment of Fitness-for-Purpose

#### Introduction

A composite outcome is a health outcome of interest (HOI) that represents a group of related conditions or events. Computable phenotype assessments for this type of HOI cannot proceed in the same way as an HOI that represents a single health event or condition. Composite outcomes require rigorous assessment of all components to determine whether or not they occurred. Component HOIs (i.e., individual members of the group of related conditions or events) each contribute their own clinical and data complexity to the composite. An overall fitness-for-purpose (FFP) recommendation for a composite HOI will weigh and synthesize all individual components' clinical and data complexity to reach a conclusion.

This document provides guidance about how to complete the final steps in an FFP assessment for a composite HOI. It assumes that separate assessments for each component HOI have already been performed using the FFP Assessment Protocol and summarized using the FFP HOI Template.

#### Methods

**Step 1.** Use the “Fitness-for-Purpose Assessment” template to draft the composite HOI FFP report, adding each component HOI FFP Assessment as a chapter. Individual HOI assessment chapters will include sections for i) Description of HOI, ii) Previous Validation Studies, iii) Data Elements, iv) Clinical Complexity, v) Data Complexity, and vi) Conclusion/FFP Recommendation. Any references or appendices associated with the component HOI chapters are placed at the end of the composite report (and not included with or after the individual chapters).

**Step 2.** Summarize the clinical complexity, data complexity and overall difficulty assessment for each component HOI in a table in the “Detailed Discussion” section for the composite HOI in the FFP report.

As an example, this information for the four component HOIs within major bleed is shown in the table below.

| Major Bleed Component HOIs | Clinical Complexity | Data Complexity | Overall Difficulty |
|---|---|---|---|
| Intracranial hemorrhage | Low | Low/Medium | Moderate |
| Hemorrhage requiring transfusion | Low | Medium | Moderate |
| Intraarticular hemorrhage | Medium | Medium | Moderate |
| Fatal hemorrhage | Low | High | Hard |

**Step 3.** Weigh the following considerations to reach a conclusion about the overall FFP for the composite HOI. Similar to the overall assessment for individual HOIs, the

overall difficulty assessment for a composite HOI does not follow a simple formula for integrating component determinations.

##### I. How many component HOIs comprise the composite HOI?

Generally speaking, the more component HOIs included in a composite, the more work will be involved in developing a computable phenotype. Recall that when we assess data complexity for a single HOI, one of the considerations is the number of data elements required (i.e., laboratory or imaging results). A greater number of elements increases data complexity (refer to the FFP worksheet). A comparable circumstance now occurs with a composite HOI whereby the component HOIs will each contribute to the cumulative effort. Having more components also increases the risk that one of them will pose unexpected challenges during development or that the resulting phenotype will fail to achieve the desired accuracy.

##### II. Should one or more of the component HOIs receive priority?

Typically, the conclusion regarding an overall assessment could be straightforward when composite HOIs all contribute equally to the FFP determination. Usually, the overall composite will have the same difficulty level as its hardest component.

However, in some circumstances, there may be reason to prioritize (or deprioritize) one or more of the component HOIs. People who conduct or use the computable phenotype may decide that some of the components are higher priority than others (e.g., because they are more common or more serious). They may decide that it is acceptable to omit a component that is particularly hard or infeasible. Under these circumstances, the overall difficulty assessment of the composite HOI will be driven by the remaining component HOIs. De-prioritizing one component may reduce the overall difficulty of a composite HOI and make it more feasible to pursue.

As an example, consider the case of major bleed. One component is fatal hemorrhage. This component is challenging to measure because out-of-hospital deaths are not captured well by either administrative claims data or EHRs. Some studies may wish to conduct linkage to the National Death Index, but this can be costly and time consuming, and there is a long time lag that may not allow for timely surveillance activities. While death is inarguably an important outcome, it is the most difficult to ascertain accurately of the components within the major bleed HOI. By deprioritizing the fatal hemorrhage component (or allowing it to be ascertained imperfectly via claims and/or EHR data), a team could make this composite outcome much more feasible to pursue. This is appealing because major bleed is an important outcome relevant to many types of medication for which safety surveillance is needed. Thus, it may be possible for the major bleed composite HOI to receive a lower overall difficulty rating than the most difficult component (fatal hemorrhage).

##### III. What is the prevalence of each component HOI?

Consider the prevalence of the individual HOIs that comprise the composite. There may be dramatic differences in prevalence, with some being much more common than others. In a composite outcome, the overall prevalence will be determined largely by the prevalence of the most common component(s). One or more components may predominate, largely washing out any impact from the rarest components.

Prevalence rates of component HOIs in the population could be a reason to weigh component HOIs differentially. You may choose to give lower priority to a rarely occurring component HOI, because its presence will make a relatively small impact on the presence of the composite. You might decide that it is acceptable to identify it with less accuracy using less labor-intensive approaches. In this case, its difficulty would contribute less weight to an overall determination for the composite HOI.

As an example, suppose a composite HOI includes two EASY and two HARD components to phenotype, but the HARD components have lower prevalence (making up <5% of cases), and the EASY components together make up 90% of cases. It is possible that your overall determination for the composite will be EASY. The rationale would be that you could focus the computable phenotype development on the two EASY components and exclude labor-intensive efforts to identify the HARD ones (perhaps relying on imperfect ascertainment using ICD-9 and 10 codes) since their prevalence is so low that they are unlikely to contribute substantially to the cases (and thus less likely to impact results of a future analysis).

##### IV. Is there overlap in the work related to each component HOI?

Typically, the workflows for individual component HOIs will be added together to produce the total work involved in creating a computable phenotype. Additionally, the more disparate these workflows are (e.g., using different types of data or approaches), the more likely the work needed will add up to create a higher overall difficulty level for the composite.

However, it is possible that the work needed to create each component HOI is overlapping or interrelated. Will there be efficiencies when you are doing the work, such that what you do for one component can be applied when creating the phenotype for a second component?

As an example, consider a composite HOI of disorders of blood cells comprised of neutropenia (low neutrophils), thrombocytopenia (low platelets), and pancytopenia (low cell counts for multiple cell lines), each of which relies on a complete blood count (CBC) laboratory test result for diagnosis, thus simplifying overall data needs. It is likely that the workflow needed to extract each component (white blood cell count, platelet count, etc.) from the lab reports will be very similar for each component, and thus the overall difficulty of creating phenotypes for several components will not be much larger/greater than for a single component.

**Step 4.** Synthesize information using the summary table from Step 1 and the considerations outlined above to synthesize the information regarding the component HOIs.

#### Example: Major Bleed

Major bleed is a composite HOI defined as: (1) an intracranial hemorrhage (a bleed within the brain) *OR* (2) an extracranial hemorrhage a) requiring red blood cell or whole blood transfusion, *or* b) involving critical anatomical sites, specifically, intraarticular (in a joint cavity), pericardial (around the heart) *or* retroperitoneal (abdominal), or c) that was fatal (resulting in death within 30 days of the bleeding event).

Major bleed consists of six component HOIs. This creates the potential for six separate computable phenotyping workflows and thus substantial work. If there was some anticipated overlap in the workflows, the overall workload may be somewhat reduced.

If the components were given equal weight, then the major bleed composite would receive a HARD overall assessment because one of the components - fatal hemorrhage - has a HARD assessment (see Table).

However, if intracranial hemorrhage was deemed to be the most important component and the fatal hemorrhage component was downgraded in its priority because of the difficulty in ascertaining it from available data, the overall assessment for major bleed could be MODERATE. This demonstrates how it may be possible for the major bleed composite HOI to receive a MODERATE overall assessment (Table) despite one component (fatal hemorrhage) having a HARD overall rating.

If hemorrhage requiring transfusion was the most prevalent of the hemorrhages of interest, then the major bleed composite could receive a MODERATE overall assessment, again by deprioritizing the fatal hemorrhage component.

For this and other rationales explained above, major bleed can be concluded to have an overall FFP assessment of MODERATE.

#### Final Note

It is important to note that assigning an overall difficulty of "HARD" does not mean that you are ruling out pursuing the development of a computable phenotype. It could be that the level of interest and importance of the composite HOI is so high that the decision will be made to pursue it anyway. The difficulty assessment is meant to provide guidance about the level of effort, resources and time needed. Ultimately, FDA teams will need to decide whether a "HARD" composite HOI should be prioritized for development, perhaps because of its overall public health importance, severity, or relevance to many different medical product exposures.

### Fitness-for-Purpose (FFP) Assessment: Neutropenia

| | |
|---|---|
| **Health Outcome of Interest:** | Neutropenia |
| **Date:** | June 28, 2024 |
| **Subject:** | Computable Phenotype Feasibility Assessment for Neutropenia |
| **Drug(s)/ ARIA Insufficiency Memo(s):** | Brodalumab, Memo #2016-1929 |

#### Executive Summary

**Overall Assessment:** *EASY* – A computable algorithm would require laboratory results from one commonly ordered blood test used to diagnose neutropenia.

**HOI Description:** Neutropenia is a disorder of white blood cells that can lead to an increased risk of bacterial infection. It is diagnosed based on the absolute neutrophil count (ANC) in the blood. Suggested cutoffs are 1500/$\mu$L (mild), 1000/$\mu$L (moderate), or 500/$\mu$L (severe), although other cut points can be used.

**ARIA Insufficiency Issue:** While diagnosis codes in claims data may have sufficient validity to identify this outcome in the inpatient setting, diagnosis codes in the outpatient setting have limited validity. The ARIA system does not include laboratory results for most Data Partners.

**Existing Computable Algorithms:** There are no existing validation studies of ICD-10 codes for neutropenia, and previous work validating ICD-9 codes in an inpatient setting used data from the 1980s and thus results may no longer apply to contemporary data due to substantial changes in healthcare practices and billing/coding. In outpatient settings, Knerr et al. (2017) and Kim et al. (2011) found a poor correspondence between ICD-9 codes and ANC. The best positive predictive value (PPV) and sensitivity from these studies were 63% and 26%, respectively (Knerr et al.), and 33% and 4% from Kim et al., although the sensitivity increased when looking at severe neutropenia (ANC < 500 $\mu$L). No literature on computable phenotypes has been identified.

**Clinical Complexity:** *LOW* – Neutropenia is diagnosed based on a single lab test, and interpretation of results is straightforward.

**Data Complexity:** *LOW* – Neutropenia requires standard lab results for neutrophil counts. In some patient populations and settings, this lab may be routinely monitored, but in others there may be missing data. In particular, cases could be missed in asymptomatic outpatient populations. Some effort may be involved in harmonizing data due to different lab reporting practices for neutrophil values, but the approach is likely feasible.

**Conclusion / Fitness-for-Purpose Recommendation:** Neutropenia case identification can be done by harmonizing lab values for ANC. Previous diagnosis code-based algorithms for inpatient neutropenia were validated using very old data and thus validity is unclear for contemporary data. Diagnosis code-based algorithms for outpatient neutropenia have performed poorly. Since large-scale validation studies using ICD-10 codes have not been completed, the FDA could consider a validation study of ICD-10 codes in both the inpatient and outpatient settings as well as a revalidation of ICD-9-based algorithms using more recent data.

**Limitations:** This fitness-for-purpose recommendation is based on literature reviews as of May 2024. We assume an ideal performance of an algorithm equates to a PPV of at least 80% together with acceptable sensitivity.

#### Detailed Discussion

##### Description of HOI

Neutropenia is a disorder of blood cells in which the level of neutrophils (the most abundant white blood cell [WBC]) in the blood are abnormally low, which can lead to an increased risk of bacterial infections. It is defined based on the concentration of neutrophils, measured with absolute neutrophil count (ANC) per microliter of blood. Clinical categorizations of neutropenia severity based on cutoffs in ANCs vary (Valent 2012). Commonly, mild neutropenia is defined as an ANC of 1000-1500/μL, moderate neutropenia as 500-1000/μL, and severe neutropenia as < 500/μL (Boxer 2012). For comparison, the WHO uses ANC ≤1800/μL to define neutropenia, and the American Society of Clinical Oncology defines an ANC of <1000/μL as neutropenia, <500/μL as severe neutropenia, and <100/μL as profound neutropenia (Taplitz 2018).

##### Drug from Drug-Outcome Pair:

1) Brodalumab, Memo #2016-1929

**Reason for ARIA insufficiency concern(s) related to outcome, according to FDA memo(s):** While diagnosis codes in claims data appear to have sufficient validity to identify this outcome in the inpatient setting (based on Strom 2001), diagnosis codes in the outpatient setting have limited validity (based on Kim 2011). The ARIA system does not include laboratory results for most Data Partners.

##### Previous Validation Studies:

- In the inpatient setting:
    - Strom (2001) synthesized data from three studies conducted in the 1990s which used data from the 1980s. They showed that ICD-9 diagnosis code 288.00 had excellent PPV (97%) in the inpatient setting. Cases with an inpatient neutropenia diagnosis during the early 1980s were identified from the COMPASS (Computerized On-Line Medicaid Pharmaceutical Analysis and Surveillance System) database of 1.7 million patients in Michigan. Exclusions consisted of any cancer diagnosis, receipt of cytotoxic drugs, recent deliveries (pregnancies) or hospitalization 30 days prior to the neutropenia diagnosis. Of the 198 inpatient neutropenia diagnoses, 192 (97%) were definite cases using medical records for validation. A limitation is that these data are 40 years old, and there have been substantial changes in medical practice since that time, including in medical care and coding and billing practices Thus, these validation results may not hold true in the modern era.
- In the outpatient setting:
    - Kim (2011) found that ICD-9 codes corresponded poorly with laboratory values. For instance, an algorithm using only ICD-9 code 288.0 had PPV 33%, while an algorithm using any ICD-9 code had sensitivity of 35% for

detecting severe neutropenia. PPVs for both mild and severe neutropenia were slightly higher (at the cost of sensitivity) for an algorithm combining several ICD-9 codes and dispensing of a medication that stimulates WBC production (PPV 22-56%; sensitivity 1-6%).

- Among populations with cancer receiving chemotherapy:
  - Knerr (2017) attempted to validate ICD-9 codes against laboratory data for both inpatient and outpatient settings for non-small cell lung cancer patients receiving chemotherapy. For an ANC < 1000, the ICD-9 code 288.0 had PPV of 63.3% and sensitivity of 26.3%.
  - Chen-Hardee (2006) reported a sensitivity of 80% for ICD-9 code 288.0 comparing hospitalization records with Medicare inpatient claims data among inpatients receiving chemotherapy. Since the study began with hospitalization records and then considered how many had the diagnosis code, PPV cannot be calculated.

**Clinical Complexity:** *LOW - diagnosis involves one lab test that is straightforward to interpret.*

Diagnosing neutropenia requires a complete blood count (CBC), yielding total white blood cell count (WBC); this must be followed with a differential to measure the neutrophil count. The diagnosis is based on the absolute neutrophil count (discussed in Description of HOI, above). While different guidelines suggest different cut points for defining the presence and/or severity of neutropenia, this should not pose problems for a computable phenotype, as the ANC could be extracted and then different cut points could be applied as desired.

**Data Complexity:** *LOW – requires commonly available laboratory data that are structured and should be easy to extract and harmonize.*

Laboratory results are necessary for identifying neutropenia. The key lab test used to make a diagnosis of neutropenia is a CBC with differential, from which the absolute neutrophil count is calculated. These laboratory results are expected to be structured, numeric values.

There is some minor complexity in interpreting lab values as neutrophils could be reported as counts or percent of WBC (meaning the ANC would need to be calculated using the percent and total WBC). Additionally, there are different types of neutrophils that could be reported separately (band neutrophils [immature cells] and segmented neutrophils [mature cells]). These values may be reported differently or in non-standard ways for different health care systems. Some data manipulation may be required. This is expected to be straightforward.

A challenge is the potential for missing data. In some populations (e.g., cancer patients receiving chemotherapy), neutrophil count may be monitored regularly at pre-specified intervals. Similarly, patients receiving a medication with a relevant REMS in place may receive ongoing monitoring. For other medications and populations, there may be no specific monitoring and labs may only be checked when clinically warranted, meaning there could be substantial missing data. This makes it likely that cases could be missed, which would probably affect primarily outpatient or asymptomatic cases.

**Data Sources:**

| Necessary EHR Data | Useful EHR Data |
|---|---|
| Lab reports (ANC or WBC and percent of neutrophils) | - |

**Conclusion / Fitness-for-Purpose (FFP) Recommendation:**

Neutropenia is easily identified from readily available laboratory data, which may require some harmonization. For healthcare settings that do not have laboratory data (e.g., many Sentinel Data Partners), it is unknown whether diagnosis code-based algorithms can reliably identify neutropenia. The definitive validation study for inpatient neutropenia was based on data from the 1980s. Results may no longer be applicable in the modern era given major changes in how healthcare is delivered, including changes in monitoring, diagnosis and treatment and also coding and billing. Studies of diagnosis code-based algorithms for outpatient neutropenia have shown poor performance. If the goal is to use only claims data, a contemporary validation study is needed to evaluate the performance of diagnosis code-based algorithms in both inpatient and outpatient settings. One important limitation of any study is that neutropenia can only be identified if a laboratory test has been performed; there may be many cases of neutropenia that are clinically undetected. It is plausible that the cases that are missed are predominantly mild and asymptomatic, which may be acceptable as long as more serious cases (the most clinically relevant) are detected with sufficient validity.

Prior to investing in the development of a computable phenotype, the FDA could consider a validation study of ICD-10 codes (in both the inpatient and outpatient settings) as well as a revalidation of ICD-9-based algorithms using more recent data. This is recommended because of major changes in clinical practice over the past 40 years, as discussed above. Such a study could be conducted within the Sentinel Innovation Center's Real-World Evidence Data Enterprise (RWE DE) network's Development Network, using available lab data and aiming to compare ICD-9 and 10 codes against varying ANC cut points. If ICD codes provide sufficient accuracy, a claims-based algorithm could be constructed that would allow studies to be carried out readily within ARIA, using data for larger populations not limited to those with available lab results. If validity of diagnosis codes were found to be low, then the FDA could proceed with developing computable phenotypes using laboratory results available in the RWE DE Commercial EHR+Claims Network.

#### Selected References

Boxer, L.A. (2012) ‘How to approach neutropenia’, Hematology / the Education Program of the American Society of Hematology. American Society of Hematology. Education Program, 2012(1), pp. 174–182.

Chen-Hardee, S., Chrischilles, E. A., Voelker, M. D., Brooks, J. M., Scott, S., Link, B. K., & Delgado, D. (2006). Population-based assessment of hospitalizations for neutropenia from chemotherapy in older adults with non-Hodgkin’s lymphoma (United States). Cancer causes & control, 17, 647-654.

Guo, L.L. et al. (2024) ‘Characterizing the limitations of using diagnosis codes in the context of machine learning for healthcare’, BMC medical informatics and decision making, 24(1), p. 51.

Kim, S.Y. et al. (2011) ‘Accuracy of identifying neutropenia diagnoses in outpatient claims data’, Pharmacoepidemiology and drug safety, 20(7), pp. 709–713.

Knerr, S., Hu, E.Y. and Zeliadt, S.B. (2017) ‘Incidence of Neutropenia in Veterans Receiving Lung Cancer Chemotherapy: A Comparison of Administrative Coding and Electronic Laboratory Data’, EGEMS (Washington, DC), 5(1), p. 1269.Strom, B.L. (2001) ‘Data validity issues in using claims data’, Pharmacoepidemiology and drug safety, 10(5), pp. 389–392.

Strom, B.L. "Data validity issues in using claims data." Pharmacoepidemiology and drug safety 10.5 (2001): 389-392.

Taplitz, R.A. et al. (2018) ‘Outpatient Management of Fever and Neutropenia in Adults Treated for Malignancy: American Society of Clinical Oncology and Infectious Diseases Society of America Clinical Practice Guideline Update’, Journal of clinical oncology: official journal of the American Society of Clinical Oncology, 36(14), pp. 1443–1453.

Valent, P. Low blood counts: immune mediated, idiopathic, or myelodysplasia. Hematology Am Soc Hematol Educ Program. 2012;2012:485-91. doi: 10.1182/asheducation-2012.1.485. PMID: 23233623.

#### Appendix 1: Clinical Guidelines for Neutropenia in the United States

Guidelines for the diagnosis and management of neutropenia in adults are issued by the European Hematology Association and the EuNet-INNOCHRON COST Action

See: Fioredda F, Skokowa J, Tamary H, Spanoudakis M, Farruggia P, Almeida A, Guardo D, Höglund P, Newburger PE, Palmblad J, Touw IP, Zeidler C, Warren AJ, Dale DC, Welte K, Dufour C, Papadaki HA. The European Guidelines on Diagnosis and Management of Neutropenia in Adults and Children: A Consensus Between the European Hematology Association and the EuNet-INNOCHRON COST Action. Hemasphere. 2023 Mar 30;7(4):e872. doi: 10.1097/HS9.0000000000000872. Erratum in: Hemasphere. 2023 Apr 27;7(5):e897. PMID: 37008163; PMCID: PMC10065839.

#### Appendix 2: ICD-9 and ICD-10 Diagnostic Codes Relevant to Neutropenia

This list of ICD-9 and 10 codes was compiled from various relevant literature, with additional codes provided by Terrence Lee at the FDA. It is not intended to be exhaustive but provided for reference purposes.

ICD-9

| Code | Description | Kim et al. 2011 | Knerr et al. 2017 |
|---|---|---|---|
| 288.0 | Neutropenia | | x |
| 288.00 | Neutropenia, unspecified | x | x |
| 288.01 | Congenital neutropenia | | |
| 288.02 | Cyclic neutropenia | | |
| 288.03 | Drug-induced neutropenia | x | x |
| 288.04 | Neutropenia due to infection | | |
| 288.09 | Other neutropenia | x | |
| 288.5 | Leukocytopenia | x | x |
| 288.59 | Other decreased WBC | x | |
| 288.8 | Specified disease of WBC | x | X |
| 776.7 | Transient neonatal neutropenia | | |

ICD-10

| Code | Description | Guo et al. 2014 |
|---|---|---|
| D70.0 | Congenital agranulocytosis | x |
| D70.3 | Neutropenia due to infection | |
| D70.4 | Cyclic neutropenia | |
| D70.8 | Other neutropenia | |
| D70.9 | Neutropenia, unspecified | |
| P61.5 | Transient neonatal neutropenia | |

### Fitness-for-Purpose (FFP) Assessment: Pericardial Effusion

| | |
|---|---|
| **Health Outcome of Interest:** | Pericardial Effusion |
| **Date:** | July 1, 2024 |
| **Subject:** | Computable Phenotype Feasibility Assessment for Pericardial Effusion |
| **Drug(s)/ ARIA Insufficiency Memo(s):** | 1. Omegaven (fish oil) 10% inj. emulsion, Memo #2018-1533 |

### Executive Summary

**Summary:** *MODERATE* - Pericardial effusion seems to show good suitability and relatively high likelihood of success for computable phenotyping by using natural language processing (NLP) applied to free text imaging reports, which are semi-structured.

**HOI Description:** Pericardial effusion is the buildup of fluid within the pericardial sac (the thin membrane that surrounds the heart).

**ARIA Insufficiency Issue:** There is concern that determining whether pericardial effusions were clinically related to the exposure of interest, omegaven, would require access to clinical narratives.

**Existing Computable Algorithms:** While there are no validated algorithms using ICD-9/10 diagnostic codes to identify pericardial effusion, keyword-based NLP algorithms show good performance across a number of studies, including Silvert *et al*. (2023), Jiang *et al*. (2018), and Nath (2016).

**Clinical Complexity:** *LOW* – Pericardial effusion is either present or absent, and it is readily detected by transthoracic echocardiography and some other forms of imaging (such as computed tomography or magnetic resonance imaging). There is not expected to be considerable variability or disagreement between clinicians in identifying pericardial effusion.

**Data Complexity:** *MEDIUM* – identifying pericardial effusions from electronic health records (EHR) data would require applying NLP to imaging reports such as echocardiography reports or possibly computed tomography (CT) reports. These reports are semi-structured free text, making NLP more straightforward than for other types of free text. ICD codes may not have good validity.

**Conclusion / Fitness-for-Purpose Recommendation:** Information on the accuracy of using diagnosis codes to identify pericardial effusions is limited. It might be of value for FDA to conduct additional validation studies focused on ICD-9/10 code-based algorithms. It is likely feasible to develop a computable phenotype for identifying pericardial effusion from certain reports within the EHR using NLP. An NLP algorithm would need to be developed and validated following the approaches described in the literature. Additional stages of algorithm development could then be carried out to address other aspects including severity, etiology (to help support causality), and specific populations (e.g., the neonatal population referenced in the original FDA memo).

**Limitations:** This fitness-for-purpose recommendation is based on literature reviews as of March 2024. We assume an ideal performance of an algorithm equates to a PPV of at least 80% with adequate sensitivity.

### Detailed Discussion

##### Description of HOI

Pericardial effusion is the buildup of extra fluid in the pericardial sac, the thin membrane that surrounds the heart. Pericardial effusions can occur after heart surgery or can result from infections, autoimmune disorders such as systemic lupus erythematosus or rheumatoid arthritis, cancer, taking certain medications, or injuries. Symptoms may resemble other medical conditions or heart problems and can include chest pain that is often described as a sharp pain in the middle or left chest, a low-grade fever, irregular heartbeat, shortness of breath, heart palpitations, or syncope as well as non-specific symptoms such as irritability, loss of appetite, or fatigue.

##### Drug from Drug-Outcome Pair:

1) Omegaven (fish oil) 10% inj. emulsion, Memo #2018-1533 - for pediatric patients, largely preterm neonates, with parenteral nutrition-associated cholestasis (PNAC)

##### Reason for ARIA insufficiency concern(s), according to FDA memo(s):

Omegaven is a fish oil-based lipid injection emulsion for pediatric patients with PNAC. Soybean oil-based lipid injection emulsions have been associated with fatal pericardial effusions,[1] and there is concern that this might result from Omegaven as well. Reviewers raised concerns including that "autopsy findings would likely be needed to definitively confirm that pleural or pericardial effusions were the cause or contributing factor in the deaths." The concern was that ARIA could not provide access to the needed clinical narratives.

##### Previous Validation Studies:

A single study reporting findings from 1223 patients with AL amyloidosis across 10 years of data from the Mayo Clinic suggested ICD codes alone would miss almost 2 out of 3 cases of pericardial effusion (Silvert 2023). They looked across fifteen signs and symptoms, including pericardial effusion, using ICD 9/10 codes, NLP, and registry data, but generally reported the combined performance across all signs and symptoms. A random sample of 10 notes which NLP indicated as containing pericardial effusion were manually reviewed and all were found to be correctly classified. Apart from these ten, false positives were not reported nor did the article provide information distinguishing between true negatives and false negatives. In

[1] Per the FDA Memo #2018-1533, "Almost all the reported deaths were in infants who received 100% soybean oil-based lipid emulsion at an infusion rate well above the recommended maximum rate of 0.15 g/kg/hr in preterm and low birthweight infants who have poor clearance of intravenous lipid emulsions."

spite of these limitations, we have calculated an upper bound on sensitivity of 35% for ICD codes, 80% for NLP, and 88% for NLP+ICD codes.[2]

Information about the performance of ICD-10 codes is available in an abstract-only publication. The study used ICD codes to identify pericardial effusion from EHR data for a single year (2018) at one clinic, the Cleveland Clinic Pericardial Disease Center. They reported a PPV of 98% and sensitivity of 76.9% (Furqan et al 2019). These results were validated by manual review. To our knowledge, a peer-reviewed article was never published from this work, so the methodological details were not available, limiting the usefulness and reliability of this work.

<u>NLP approaches using EHR data</u>

There are no validated NLP algorithms targeting pericardial effusion. Keyword-based algorithms, however, are described in three papers, one that focused on detecting AL amyloidosis symptoms (Silvert et al. 2023), another that sought to determine risk of small cell lung cancer (Jiang et al. 2018), and a third that extracted complete data from echocardiogram reports (Nath 2016). The last publication references a tool called EchoInfer which, while not updated in the last 8 years, could provide a useful starting point in developing an NLP algorithm.  This suggests that NLP approaches to identifying pericardial effusion are feasible using echocardiograms and radiology but will take effort to develop and validate.

**Clinical Complexity:** *LOW – a pericardial effusion is either present or absent. We do not anticipate substantial disagreement or variation between clinicians in making the diagnosis.*

For determining the presence vs. absence of a pericardial effusion, clinical complexity is deemed to be low. Pericardial effusions are identified clinically using transthoracic echocardiography (the gold standard) or other imaging studies (such as computed tomography). There are clear and validated diagnostic guidelines. Most of the guidelines focus on guiding further evaluation to determine the underlying cause of the effusion. This seems relevant to the FDA memo because of the expressed concern about determining causality.

Validated diagnostic guidelines are available from the European Society of Cardiology (ESC; Adler et al. 2015). ESC guidelines include recommendations for clinical workups that vary by the suspected etiology of the pericardial effusion (Spangler 2019). The workup is directed at identifying the underlying cause of the effusion, rather than the presence or absence of an effusion. The initial evaluation includes a clinical history and physical examination; electrocardiogram (ECG); chest

[2] These were calculated by assuming that all patients not identified by ICD codes, NLP, or registry were correctly classified as negative (which is an assumption the paper largely makes).

radiography, echocardiography, CT scanning, cardiovascular magnetic resonance imaging (CMRI), positron-emission tomography (PET) scanning, or PET/CT scanning, as indicated; and initial laboratory work. Pericardiocentesis should be performed on all patients with cardiac tamponade or suspected purulent pericarditis.

In addition to presence vs. absence of an effusion, there are other aspects of this HOI that may be of interest, including whether it is symptomatic, clinical severity (e.g., was the effusion considered life-threatening), and the underlying cause (relevant for the FDA memo, determining whether it consisted of lipids, supporting causality.) Attempting to assess these aspects of the HOI from the EHR would add greatly to the assessed clinical complexity and would require multiple stages of algorithm development.

**Data Complexity:** *MEDIUM – NLP algorithm would be needed to identify pericardial effusion from echocardiography reports; relies on free text but limited types of medical records; ICD-9/10 codes may not have good validity.*

Information about pericardial effusion would need to be extracted via NLP from free-text reports of procedures, primarily echocardiograms but potentially also imaging studies (computed tomography [CT], magnetic resonance imaging [MRI]). Any time that NLP is needed it introduces complexity and indicates a complexity level of MEDIUM or HARD. Imaging and echocardiogram reports have internal structure and tend to use a limited vocabulary and some repetitive or stereotyped language, making them more amenable to NLP than less structured reports (such as clinical notes). Prior reports of successful NLP efforts (see above) support the idea that data complexity would be medium rather than hard. In some cases, desired reports are found in the EHR as PDFs, which adds complexity.

A significant proportion of patients with pericardial effusion are asymptomatic, and many findings of pericardial effusion on imaging or echocardiogram are incidental, with the imaging performed for other reasons (Adler 2015).

| Necessary EHR Data | Useful EHR Data |
|---|---|
| Echocardiography report | Radiology reports (CT, MRI) |

**Conclusion / Fitness-for-Purpose (FFP) Recommendation – *MODERATE DIFFICULTY***

Information on the accuracy of using diagnosis codes to identify pericardial effusions is limited and contradictory. It might be of value for FDA to conduct additional validation studies focused on ICD-9/10 code-based algorithms, including for subgroups of particular interest (such as neonates or pediatric populations) where the clinical context, underlying illnesses, and treatment patterns may differ substantially from the general population.

A computable algorithm could be developed by applying NLP to reports in the EMR, predominantly echocardiogram reports but also imaging reports. These semi-structured free text notes are more amenable to NLP than less structured EHR notes. We conclude that NLP approaches would be feasible but will take effort to develop and validate. An initial approach could be broadly directed at identification of the presence/absence of pericardial effusions. Additional stages of algorithm development could then be carried out to address severity, etiology (to help support causality), and specific populations (such as the neonatal population referenced in the original FDA memo).

#### References

Adler Y, et al. (2015). 2015 ESC Guidelines for the diagnosis and management of pericardial diseases: The Task Force for the Diagnosis and Management of Pericardial Diseases of the European Society of Cardiology (ESC)Endorsed by: The European Association for Cardio-Thoracic Surgery (EACTS). Eur Heart J. 2015 Nov 7;36(42):2921-2964. doi: 10.1093/eurheartj/ehv318. Epub 2015 Aug 29. PMID: 26320112; PMCID: PMC7539677.

Bach D. S. (2015) 2015 ESC Guidelines for Pericardial Disease. Available at: https://www.acc.org/latest-in-cardiology/ten-points-to-remember/2015/10/30/12/01/2015-esc-guidelines-for-the-diagnosis-and-management-of-pericardial-diseases (Accessed: 19 March 2024).

Cheng, C.-Y. et al. (2023) 'Development and validation of a deep learning pipeline to measure pericardial effusion in echocardiography', Frontiers in Cardiovascular Medicine, 10. Available at: https://doi.org/10.3389/fcvm.2023.1195235.

Furqan, M.M. et al. (2019) 'Abstract 15826: Misclassification of Pericardial Disease Diagnosis; International Classification of Diseases-10 Coding Requires Validation', Circulation, 140(Suppl_1), pp. A15826–A15826.

Jiang R et al. Incidence of pre-treatment and off-treatment pericardial and pleural effusions by line of therapy in patients with small cell lung cancer: An analysis of electronic health records data. JCO 36, e20569-e20569(2018).doi:10.1200/JCO.2018.36.15_suppl.e20569

Meystre, S. et al. (2017) 'Enhancing Comparative Effectiveness Research With Automated Pediatric Pneumonia Detection in a Multi-Institutional Clinical Repository: A PHIS+ Pilot Study', Journal of medical Internet research, 19(5), p. e162.

Nath, C., Albaghdadi, M.S. and Jonnalagadda, S.R. (2016) 'A Natural Language Processing Tool for Large-Scale Data Extraction from Echocardiography Reports', PloS one, 11(4), p. e0153749.

Niehues, S.M. et al. (2021) 'Deep-Learning-Based Diagnosis of Bedside Chest X-ray in Intensive Care and Emergency Medicine', Investigative radiology, 56(8), pp. 525–534.

Shakti, D. et al. (2014) 'Idiopathic pericarditis and pericardial effusion in children: contemporary epidemiology and management', Journal of the American Heart Association, 3(6), p. e001483.

Silvert, E. et al. (2023) 'Identifying signs and symptoms of AL amyloidosis in electronic health records using natural language processing, diagnosis codes, and manually abstracted registry data', American journal of hematology, 98(9), pp. E255–E258.

Spangler, S. et al (2019). Acute Pericarditis Workup. Available at: https://emedicine.medscape.com/article/156951-workup (Accessed: 19 March 2024).

#### Appendix 1: Clinical Guidelines for Pericardial Effusion

Guidelines for the diagnosis and management of pericardial effusion are issued by the American College of Cardiology (ACC) who recommend following the guidance of the European Society of Cardiology (ESC).

| Reference | Society Guideline |
|---|---|
| Adler et al. (2015) | 2015 ESC Guidelines for the diagnosis and management of pericardial diseases. |
| Bach et al. (2015) | ACC recommendation to follow 2015 ESC guidelines. |

### Appendix 2: ICD-9 and ICD-10 Diagnostic Codes Relevant to Pericardial Effusion

This list of ICD-9 and -10 codes was compiled from various relevant literature. It is not intended to be exhaustive but provided for reference purposes.

ICD-9

| Code | Description | Source |
|---|---|---|
| 420.90 | Effusion, pericardium, acute | Silvert (2023) |
| 423.3 | Cardiac tamponade | |

ICD-10

| Code | Description | Source |
|---|---|---|
| I31.3 | Pericardial effusion (non-inflammatory) | |
| I30.9 | Acute pericarditis, unspecified | |
| I31.4 | Cardiac tamponade | |

Note: Procedure codes for pericardiocentesis and pericardial window surgical procedure would also be relevant and useful; researching them is beyond the scope of this report but will be a useful step for a future team charged with developing an algorithm.

### Fitness-for-Purpose (FFP) Assessment:
### Drug-Induced Liver Injury (DILI)

| | |
|---|---|
| **Health Outcome of Interest:** | Drug-Induced Liver Injury (DILI) |
| **Date:** | June 28, 2024 |
| **Subject:** | Computable Phenotype Feasibility Assessment for Drug-Induced Liver Injury |
| **Drug(s)/ ARIA Insufficiency Memo(s):** | 1. Tolvaptan, Memo #2017-2184<br>2. Zinbryta™ (Daclizumab – High Yield Process), Memo #2016-588<br>3. SKYRIZI (risankizumab-rzaa), Memo #2022-838 |

#### Executive Summary

**Overall Assessment:** *HARD* - Computable algorithms are unlikely to reliably capture DILI.

**HOI Description:** DILI is defined as any injury to the liver caused by a medication or herbal/dietary supplement.

**ARIA Insufficiency Issue:** Laboratory results for liver tests, including alanine transferase (ALT) and alkaline phosphatase (ALP), must be compared to baseline laboratory values obtained prior to starting a medication. The ARIA system does not include laboratory results for most Data Partners. ICD-9 code-based identification of cases of severe acute liver injury has been shown to be poor.

**Existing Computable Algorithms:** Weng and Overby (2012) present the most complete computable phenotyping algorithm, which used labs, medications, ICD-9 codes, and Natural Language Processing (NLP) of clinical notes. Performance measures were low with PPVs ranging from 16-39% across three sites and sensitivity estimated at 5.5%. Studies using ICD-10 codes in Europe found higher performance for certain toxic liver disease codes (K71) when validated against laboratory values, but confidence intervals were wide and there is no information about sensitivity.

**Clinical Complexity:** *HIGH* – because numerous other potential causes of liver injury must be ruled out to reach a diagnosis, and there is no consensus on clinical diagnostic criteria.

**Data Complexity:** *HIGH* – primarily because computable phenotypes are needed to identify/exclude many other possible causes of liver injury and also because longitudinal laboratory values would be needed. Developing a computable phenotyping algorithm would likely involve diagnosis codes, laboratory data, and features derived from NLP.

**Conclusion / Fitness-for-Purpose Recommendation:** Previous diagnosis code-based algorithms from the ICD-9 era have performed poorly, and computable algorithms are unlikely to reliably capture DILI. However, computable phenotyping may add value to case identification when ICD-10 diagnostic codes are integrated for liver injury and for conditions being ruled out, which could more precisely identify potential cases. Since large-scale validation studies using ICD-10 codes have not yet been completed, FDA could consider a validation study of ICD-10 codes. Alternatively, a computable phenotype could be developed for case finding, with chart review used to validate potential cases.

**Limitations:** This fitness-for-purpose recommendation is based on literature reviews as of April 2024. We assume an ideal performance of an algorithm equates to a PPV of at least 80% together with acceptable sensitivity.

### Detailed Discussion

**<u>Description of HOI</u>**

Drug-induced liver injury (DILI) is defined as injury to the liver caused by a medication, including prescription medications, over-the-counter medications, and herbal/dietary supplements. A diagnosis is based in part on liver biochemistry tests, with abnormal values indicated by either (1) aspartate aminotransferase (AST) or alanine aminotransferase (ALT) >5 times the upper limit of normal (ULN) and/or alkaline phosphatase (ALP) >2 times ULN (Danan & Teschke, 2016) (or pretreatment baseline if baseline is abnormal) on two separate occasions at least 24 hours apart; (2) total serum bilirubin >2.5 mg/dl with elevated serum AST, ALT, or ALP; or (3) international normalized ratio (INR) >1.5 with elevated serum AST, ALT, or ALP (Fontana et al., 2023; Senior 2014). The signs and symptoms of DILI include fever, nausea, vomiting, anorexia, pruritis (itching), abdominal pain, jaundice, coagulopathy, confusion, or coma. Patients may present with hepatomegaly (enlarged liver) on physical examination. DILI (and liver injury more generally) can be characterized as hepatocellular (necrosis or damage to hepatocytes), cholestatic (abnormal biliary secretion due to biliary obstruction), or mixed hepatocellular and cholestatic injury. The ratio of ALT to ALP or R-value is used to characterize the pattern of liver injury with R > 5 labeled as hepatocellular, R < 2 cholestatic, and 2 < R < 5 mixed (Chalasani et al., 2021). Idiosyncratic DILI is uncommon, occurring in only 1 in 1000 to 1 in a million exposed individuals.

**<u>Information from FDA memos:</u>**

**Drug from Drug-Outcome Pair:**

1) Tolvaptan, Memo #2017-2184
2) Zinbryta™ (Daclizumab – High Yield Process), Memo #2016-588
3) SKYRIZI (risankizumab-rzaa), Memo #2022-838

**Reason for ARIA insufficiency concern(s) related to outcome, according to FDA memo(s):** Laboratory results for alanine transferase (ALT) and alkaline phosphatase (ALP) will be needed. The ARIA system does not include laboratory results for most Data Partners. ICD-9 code-based identification of cases of severe acute liver injury has been shown to be poor.

**Previous Validation Studies:**

In summary, there is currently no existing high-performing computable phenotype and no validated high-performing algorithm based on ICD-9 or ICD-10 diagnosis codes.

- <u>Previously described computable phenotype</u>: Weng and Overby (2012) present the most complete algorithm described in the literature which was deployed across three healthcare systems. The algorithm relies first on labs (to diagnose liver injury and confirm normal measurements prior to drug exposure) and medications (to determine recent exposure to a new drug) to select a cohort of possible cases. Both diagnosis codes (ICD-9) and Natural Language Processing (extracting relevant codes from the Unified Medical Language System [UMLS] using a tool such as MedLEE) applied to discharge summaries or all clinical notes are then used to exclude all patients with documented cancer, gallbladder disease, pancreatic disease, heart failure, alcohol abuse, liver damage, toxic effects, HIV infection, rheumatoid arthritis, sarcoidosis, systemic lupus, viral hepatitis, organ transplantation, therapeutic operations for the liver, chronic liver injury, and overdoses. Weng and Overby attempted to exclude chronic cases from their definition. They reported PPVs ranging from 16% to 39% with an estimated sensitivity (at one site) of 5.5%.[1] They also recommend that hospital discharge summaries (rather than all clinical text) be used to improve PPV.[2]

- European studies have sought to validate the performance of **ICD-10 codes**, primarily the code for "toxic liver disease" (K71). Validation of the codes was performed by checking for concurrent elevated liver enzymes at the time of diagnosis (and, depending on the study, ensuring the individuals were receiving a particular medication). Then, depending on the study, they included the potential case only when there were no elevated levels in a prior time period (e.g., the 24 months prior to the elevation) or when the elevation lasted less than a year. When insufficient lab data were available, the cases were not evaluated. The total number of potential cases was small, even when the study looked across a decade, with sample size typically <50. The studies presented results separately by individual ICD-10 code and, while a single code may appear to perform with high PPV, the confidence intervals are very wide due to small numbers of cases. A major limitation is that these algorithms did not attempt to exclude other potential causes of liver injury. Also, information on sensitivity is typically not reported.
    - Forns *et al*. (2019) reported PPVs for inpatient codes ranged from 60.0% to 84.2%, and adding codes from the outpatient setting gave a range from 34.8% to 83.3%.
    - Giunta *et al*. (2024) performed a validation study in a population in Sweden receiving antifungal drug treatment using ICD-10 codes for toxic liver disease and reported a PPV of 53.8% (CI 33.4-73.4%).
    - Timmer *et al*. (2019) report a PPV of at most 62.7%.

**Clinical Complexity:** *HIGH – clinical diagnosis requires temporal association with a drug and exclusion of other potential causes of liver injury.*

Once acute liver injury has been identified based on abnormal laboratory results, DILI is a diagnosis of exclusion, meaning that other potential causes of liver injury must be ruled out (Senior & Guo 2018, 420-5). A diagnosis relies on a detailed medical

[1] Overby *et al.* (2013a) describes error analysis in which 'many results were correct for the algorithm but inappropriate given the DILI case definition' (133). E.g., 'Overdose patients were inappropriate because the cause of DILI was already known for these patients.'
[2] Overby *et al.* (2013b), however, note that chart abstractors observed descriptions in other note types (namely, outpatient notes).

history including medication exposure and the pattern and course of liver biochemistry tests before and after drug discontinuation. While society guidelines for DILI exist in the US (i.e., AASLD [Fontana *et al*. 2023], ACG [Chalasani *et al*. 2021]), there is no single unified set of criteria, and available diagnostic algorithms for clinicians lack clarity and proven accuracy (Chalasani *et al*. 2021). Scales are available that codify causality of drug toxicity, but there is high variability between observers in classifying cases (Lucena *et al*. 2001).

**Data Complexity:** *HIGH – data required to identify/exclude many of the separate potential causes of liver injury are complex, covering multiple domains including free text (requiring NLP).* The amount and complexity of data required for a computable phenotype for DILI would be substantial. Both inpatient and outpatient EHR data will be needed, and data from these settings may not be accessible within the same health system. For instance, patients admitted to a hospital with DILI may have relevant data about their current status in that healthcare system, but key baseline data may be stored in the EHR from their primary care setting, which may be in a different healthcare system. Longitudinal lab data will be needed to allow comparison of baseline laboratory values to values during the acute illness. NLP algorithms will be required to extract information from free text, particularly physician notes, to rule out some potential alternative causes. For some of these potential causes, there may be a very wide range of words or phrases that could be used (e.g., use of supplements). The needed information is likely to be stored in multiple locations in the EHR. Information about some potential causes (e.g., heavy alcohol use) may not be available in the EHR (because patients may not seek care or physicians may not ask about it) or if they are, may not be recorded in structured variables. ICD codes for DILI as well as for other exclusionary causes may not have good validity.

**Data Sources to Identify Liver Injury:**

| Necessary EHR Data | Useful EHR Data[3] |
|---|---|
| Lab reports (liver chemistry tests) | Radiology reports (hepatobiliary imaging) |
| | Pathology reports (liver biopsy) |

**Data Elements to Rule Out Other Causes of Liver Injury:**

*Note: Having any of these conditions does not automatically rule out that a patient also has DILI.*

**Relatively easy:**

| Other potential cause | Rationale for easy rating (data needed) |
|---|---|
| Viral infection (e.g., viral hepatitis, mononucleosis) | Could use diagnostic codes or laboratory tests related to viral infection (e.g., serologic tests; HBV DNA.) |

[3] Chalasani *et al.* (2021)

| | |
|---|---|
| Liver cancer | Could use diagnostic codes to identify cancer in the liver |
| Budd-Chiari syndrome | Could use diagnostic codes |
| Wilson disease (hepatolenticular degeneration) | Could use diagnostic codes or results of genetic testing |
| Pregnancy-related liver conditions *(e.g., acute fatty liver of pregnancy, intrahepatic cholestasis of pregnancy)* | Could use diagnostic codes |
| Other prescription medications known to cause liver injury | Could use dispensings or orders for implicated medications |
| Ischemic hepatitis | Could use diagnostic codes and labs |
| Sclerosing cholangitis | Could use diagnostic codes and labs |
| Hemochromatosis | Could use diagnostic codes and labs |
| Primary biliary cholangitis | Could use diagnostic codes and labs |
| Biliary obstruction | Could use diagnostic codes (if natural language processing on imaging reports needed to identify accurately, then would be hard) |
| Gallbladder disease | Could use diagnostic codes |
| Pancreatic disease | Could use diagnostic codes and labs |
| Organ transplantation | Could use diagnostic codes |
| Heart failure | A fairly accurate algorithm can likely be identified using diagnostic codes, possibly also including labs. |

**Relatively hard:**

| Other potential cause | Rationale for hard rating (data needed) |
|---|---|
| Alcohol use disorder | Could use diagnostic codes, but this condition is under-reported, under-recognized and under-coded. Diagnosis codes may not reflect current ongoing alcohol use. Would likely need natural language processing of clinical notes to determine with enough accuracy, and information might be absent from notes. |
| Herbal supplements | Need natural language processing of clinical notes to determine use of herbal supplements, and a very wide variety of herbal supplements may be of interest. |
| Metabolic associated fatty liver disease (MAFLD; formerly nonalcoholic fatty liver disease) | Could use diagnostic code for MAFLD while raising the cutoff level of ALT elevation to 5x ULN. MAFLD itself is a diagnosis of |

| | |
|---|---|
| | exclusion, making the assessment of causality challenging. |
| Alpha-1-antitrypsin deficiency | Could use diagnostic codes and common labs, but also need a genetic test and natural language processing on imaging reports |
| Specific autoimmune conditions that can cause liver injury (rheumatoid arthritis, sarcoidosis, systemic lupus erythematosus) | Could use diagnostic codes and labs, but difficult to diagnose accurately |
| Autoimmune hepatitis | Many lab tests that are commonly done to identify, but can be difficult to interpret |
| Lupus hepatitis | Many lab tests that are commonly done to identify, but can be difficult to interpret |

**Conclusion / Fitness-for-Purpose (FFP) Recommendation – *HIGH DIFFICULTY***

Previous diagnosis code-based algorithms from the ICD-9 era have performed poorly, and it will be difficult to develop a computable algorithm to capture DILI. Challenges include high clinical complexity due to the need to exclude a large number of alternative causes and the lack of agreement across clinical guidelines. Data complexity is high due to the need for a large number of data elements across diverse data types, primarily to rule out competing causes of liver injury. The assessment of liver injury itself would require longitudinal lab data, which might be located in different healthcare settings and systems. Thus it seems very challenging to develop a computable phenotype for DILI that could have high PPV and sensitivity. There is potential for computable phenotyping to be useful for enhanced case identification, with subsequent medical record review to validate cases.

#### Selected References

Bui, C.L. et al. (2014) 'Validation of acute liver injury cases in a population-based cohort study of oral antimicrobial users', *Current drug safety*, 9(1), pp. 23–28.

Chalasani, N.P., Maddur, H., Russo, M.W., Wong, R.J., Reddy, K.R; on behalf of the Practice Parameters Committee of the American College of Gastroenterology (2021). ACG Clinical Guideline: Diagnosis and Management of Idiosyncratic Drug-Induced Liver Injury. *The American Journal of Gastroenterology* 116(5):p 878-898, May 2021. | DOI: 10.14309/ajg.0000000000001259

Danan G, Teschke R. RUCAM in Drug and Herb Induced Liver Injury: The Update. International Journal of Molecular Sciences. 2016; 17(1):14. https://doi.org/10.3390/ijms17010014

Fontana, R.J.; Liou, I.; Reuben, A.; Suzuki, A.; Fiel, M.I; Lee, W., Navarro, V. (2023). AASLD practice guidance on drug, herbal, and dietary supplement–induced liver injury. Hepatology 77(3):p 1036-1065, March 2023. | DOI: 10.1002/hep.32689

Forns, J. et al. (2019) 'Validity of ICD-9 and ICD-10 codes used to identify acute liver injury: A study in three European data sources', *Pharmacoepidemiology and drug safety*, 28(7), pp. 965–975.

Giunta, D.H. et al. (2024) 'Validation of diagnoses of liver disorders in users of systemic azole antifungal medication in Sweden', *BMC gastroenterology*, 24(1), p. 21.

Lo Re V, 3rd, et al. (2012). *Validity of Diagnostic Codes to Identify Cases of Severe Acute Liver Injury in the Mini-Sentinel Distributed Database*. Available at: https://www.sentinelinitiative.org/sites/default/files/surveillance-tools/validations-literature/Mini-Sentinel_Validation-of-Severe-Acute-Liver-Injury-Cases.pdf (Accessed 19 March 2024).

Lo Re, V, 3rd et al. (2013) 'Validity of diagnostic codes to identify cases of severe acute liver injury in the US Food and Drug Administration's Mini-Sentinel Distributed Database', Pharmacoepidemiology and drug safety, 22(8), pp. 861–872.

Lucena MI, Camargo R, Andrade RJ, Perez-Sanchez CJ, Sanchez De La Cuesta F. Comparison of two clinical scales for causality assessment in hepatotoxicity. Hepatology. 2001 Jan;33(1):123-30. doi: 10.1053/jhep.2001.20645. PMID: 11124828.

Overby, C.L. et al. (2013a) 'A collaborative approach to developing an electronic health record phenotyping algorithm for drug-induced liver injury', Journal of the American Medical Informatics Association: JAMIA, 20(e2), pp. e243–52.

Overby, C.L. et al. (2013b) 'Evaluation considerations for EHR-based phenotyping algorithms: A case study for drug-induced liver injury', AMIA Joint Summits on Translational Science proceedings. AMIA Joint Summits on Translational Science, 2013, pp. 130–134.

Senior, J.R. Evolution of the Food and Drug Administration approach to liver safety assessment for new drugs: current status and challenges. Drug Saf. 2014 Nov;37

Suppl 1(Suppl 1):S9-17. doi: 10.1007/s40264-014-0182-7. PMID: 25352324; PMCID: PMC4212154.

Senior, J. & Guo, T. (2018). 'Hy's Law and eDISH for Clinical Studies'. In: Chen, M., Will, Y. (eds) *Drug-Induced Liver Toxicity*. Methods in Pharmacology and Toxicology. Humana, New York, NY. https://doi.org/10.1007/978-1-4939-7677-5_20

Timmer, A. *et al.* (2019) 'Validity of hospital ICD-10-GM codes to identify acute liver injury in Germany', *Pharmacoepidemiology and drug safety*, 28(10), pp. 1344–1352.

Udo, R. *et al.* (2016) 'Validity of diagnostic codes and laboratory measurements to identify patients with idiopathic acute liver injury in a hospital database', *Pharmacoepidemiology and drug safety*, 25 Suppl 1, pp. 21–28.

Wang, X, Xiaowei X, Weida T, Qi L, and Zhichao L. 2022. "DeepCausality: A General AI-Powered Causal Inference Framework for Free Text: A Case Study of LiverTox." *Frontiers in Artificial Intelligence* 5 (December): 999289.

Weng C and Overby C. (2012). *Drug Induced Liver Injury*. PheKB. Available from: https://phekb.org/phenotype/135.

### Appendix 1: Clinical Guidelines for Drug-Induced Liver Injury in the United States

- American Association for the Study of Liver Diseases (AASLD): Practice guidance on drug, herbal, and dietary supplement-induced liver injury (Fontana 2023)
    - Clinically significant DILI is commonly defined as any one of the following: (1) serum AST or ALT >5× upper limit of normal (ULN) or ALP >2× ULN (or pretreatment baseline if baseline is abnormal) on two separate occasions at least 24 h apart; (2) total serum bilirubin >2.5 mg/dl along with elevated serum AST, ALT, or ALP level; or (3) INR >1.5 with elevated serum AST, ALT, or ALP.
- American College of Gastroenterology (ACG): Clinical guideline for the diagnosis and management of idiosyncratic drug-induced liver injury (Chalasani *et al.* 2021)
    - The R-value is defined as serum alanine aminotransferase (ALT)/upper limit of normal (ULN) divided by serum alkaline phosphatase (Alk P)/ULN. By common convention, R > 5 is labeled as hepatocellular DILI, R < 2 is labeled as cholestatic DILI, and 2 < R < 5 is labeled as mixed DILI.

### Appendix: ICD-9 and ICD-10 Diagnostic Codes Relevant to Liver Injury

This list of ICD-9 and -10 codes was compiled from various relevant literature. It is not intended to be exhaustive but provided for reference purposes.

ICD-9[4]

| Code | Description |
|---|---|
| 570 | Acute and subacute necrosis of the liver |
| 572.2 | Hepatic coma |
| 572.4 | Hepatorenal syndrome |
| 572.8 | Sequelae of liver disease |
| V42.7 | Liver replaced by transplant |
| 573.3 | Toxic (non-infectious) hepatitis |
| 573.8 | Other specified disorder of the liver |

ICD-10[5]

| Code | Description |
|---|---|
| K71.0 | Toxic liver disease with cholestasis |
| K71.1 | Toxic liver disease with hepatic necrosis |
| K71.10 | Toxic liver disease with hepatic necrosis without coma |
| K71.11 | Toxic liver disease with hepatic necrosis with coma |
| K71.2 | Toxic liver disease with acute hepatitis |
| K71.3 | Toxic liver disease with chronic persistent hepatitis |
| K71.4 | Toxic liver disease with chronic lobular hepatitis |
| K71.5 | Toxic liver disease with chronic active hepatitis |
| K71.50 | Toxic liver disease with chronic active hepatitis without ascites |
| K71.51 | Toxic liver disease with chronic active hepatitis with ascites |
| K71.6 | Toxic liver disease with hepatitis, not elsewhere classified |
| K71.7 | Toxic liver disease with hepatitis with fibrosis and cirrhosis of the liver |
| K71.8 | Toxic liver disease, unspecified |

[4] Evaluated by Lo Re V, 3rd, *et al.* (2012)

[5] https://www.clinicalkey.com/#!/content/derived_clinical_overview/76-s2.0-B978032375576400288X

### Fitness-for-Purpose (FFP) Assessment: Serious Infections

| | |
|---|---|
| **Health Outcome of Interest:** | Serious Infections |
| **Date:** | September 16, 2024 |
| **Subject:** | Computable Phenotype Feasibility Assessment |
| **Drug(s)/ ARIA Insufficiency Memo(s):** | 1. Zinbryta (Daclizumab – High Yield Process), Memo #2016-588<br>2. Sutimlimab, Memo #2022-67 |
| **Resolution:** | **The serious infections HOI is not ready for evaluation** |

#### Summary

The workgroup has determined that the “serious infections” health outcome of interest (HOI) is not amenable to a fitness-for-purpose assessment without several areas of ambiguity first being resolved, specifically, the definition of “serious” as well as specifying which infections are of primary interest.

#### Challenges to be Addressed:

Serious infections is a composite HOI. Overall, there is not a clear or widely-accepted definition for what constitutes a “serious infections”. Studies have taken heterogeneous approaches to define the concept, including variability in which and how many infections are included. In some definitions, one or more of the component infections are themselves ill-defined.

- **Society or regulatory guidelines**: No definition of serious infections has been published by a professional society, nor is there regulatory guidance about how this outcome should be defined.
- **Significance of “serious”**: There is no consensus on what features or characteristics indicate that an infection is “serious.” Some but not all studies or algorithms consider “serious” as requiring hospitalization. For example, Lo Re *et al*. (2021) required an inpatient hospitalization in their definition, whereas an international consensus definition for adults aged 65 years or older developed by a panel of physicians defined “serious” based on “a high risk of complications, functional decline, and/or mortality, requiring a prompt diagnostic and therapeutic approach in the appropriate care setting” (Struyf *et al*. 2020). The latter would be difficult to operationalize, particularly with electronic health data.
- **Selection and number of infections**: Studies have varied greatly in their choice of which infections and how many to include. For example, Lo Re et al. specified seven different infections or types of infection (e.g., pneumonia, meningitis, gastrointestinal infections), while Zhou *et al*. (2023) published an algorithm including 58 different infections. A systematic review of validation studies by Barber *et al*. (2013) also demonstrates this point: four articles cited as addressing “serious infections” in a broad sense include different numbers and types of infections in their definition. It is not clear what criteria or evidence should guide the choice of infections. In addition, some of the published definitions include components that are themselves vague and ill-defined. For instance, the Lo Re *et al*. (2021) algorithm for identifying serious infections includes “gastrointestinal infections” -- a category that encompasses a large number of heterogeneous infections caused by different organisms and with differing prognoses.

- **Other considerations**: Studies have taken a variety of approaches concerning how to handle nosocomial (hospital-acquired) infections with respect to "serious infections". For example, on one end of the spectrum, Lo Re *et al*. (2021) takes great pains to exclude these while Zhou *et al*. (2023) includes nosocomial infections in their definition of "serious infections". Most other studies attempt to exclude them, but either does not apply as stringent criteria as Lo Re (e.g., Schneeweiss *et al*. [2007]) and Grijalva *et al*. [2008]), or does not provide a definition (e.g., Jackson *et al*. [2003]).

The issues described above would need to be considered and clarified before a fitness-for-purpose assessment could be carried out.

#### References

Barber, C., Lacaille, D. and Fortin, P. R. (2013) Systematic review of validation studies of the use of administrative data to identify serious infections: Administrative data to identify infections, Arthritis care & research. Wiley, 65(8), pp. 1343–1357.

Feldman, C. H. et al. (2017) Comparative rates of serious infections among patients with systemic lupus erythematosus receiving immunosuppressive medications, Arthritis & rheumatology. NIH Public Access, 69(2), pp. 387–397.

Grijalva, C. G. et al. (2008) Computerized definitions showed high positive predictive values for identifying hospitalizations for congestive heart failure and selected infections in Medicaid enrollees with rheumatoid arthritis, Pharmacoepidemiology and drug safety. NIH Public Access, 17(9), pp. 890–895.

Jackson, L. A. et al. (2003) Effectiveness of pneumococcal polysaccharide vaccine in older adults, NEJM. 348(18), pp. 1747–1755.

Lacaille, D. et al. (2008) Use of nonbiologic disease-modifying antirheumatic drugs and risk of infection in patients with rheumatoid arthritis, Arthritis and rheumatism 59(8), pp. 1074–1081.

Lo Re, V., 3rd et al. (2021) Validity of ICD-10-CM diagnoses to identify hospitalizations for serious infections among patients treated with biologic therapies, Pharmacoepidemiology and drug safety. Wiley, 30(7), pp. 899–909.

Schneeweiss, S. et al. (2007) Veteran's affairs hospital discharge databases coded serious bacterial infections accurately, Journal of clinical epidemiology. Elsevier BV, 60(4), pp. 397–409.

Simard, J. F. et al. (2021) Infection hospitalisation in systemic lupus in Sweden, Lupus science & medicine. BMJ, 8(1), p. e000510.

Struyf T, Tournoy J, Verbakel JY, Van den Bruel A. International Consensus Definition of a Serious Infection in a Geriatric Patient Presenting to Ambulatory Care. J Am Med Dir Assoc. 2020 May;21(5):578-582.e1. doi: 10.1016/j.jamda.2020.01.015. Epub 2020 Feb 25. PMID: 32111485.

Zhou, V. Y. et al. (2023) Risk of severe infections after the introduction of biologic DMARDs in people with newly diagnosed rheumatoid arthritis: a population-based interrupted time-series analysis, Rheumatology (Oxford, England). 62(12), pp. 3858–3865.

### Fitness-for-Purpose (FFP) Assessment: Venous Thromboembolism

| | |
|---|---|
| **Health Outcome of Interest:** | Venous Thromboembolism |
| **Date:** | December 13, 2024 |
| **Subject:** | Computable Phenotype Feasibility Assessment for Venous Thromboembolism |
| **Drug(s)/ ARIA Insufficiency Memo(s):** | 1. Abrocitinib (Cibinqo), Memo #2021-1436<br>2. Upadacitinib (RINVOQ), Memo #2021-276 |

#### Executive Summary

**Summary:** *MODERATE* – Venous thromboembolism (VTE) appears to have good feasibility and high likelihood of success for computable phenotyping via natural language processing (NLP) applied to imaging reports.

**HOI Description:** VTE refers to a blood clot in the veins. There are 2 main types of VTE: deep vein thrombosis (DVT), where a blood clot forms in a deep vein, typically in the upper or lower extremities, and pulmonary embolism (PE), when a clot occurs in arteries in the lungs.

**ARIA Insufficiency Issue:** Concern that administrative data such as diagnosis codes do not have sufficient specificity, positive predictive value (PPV), or sensitivity to identify VTE accurately outside of settings with high incidence and awareness of heightened risk, e.g., following orthopedic surgery.

**Existing Computable Algorithms:** There is a large literature showing that algorithms using ICD-10 diagnosis codes do not have sufficient PPV when used in relatively healthy populations that are not post-surgery or already hospitalized when the VTE occurs. Evidence from multiple studies shows that basic natural language processing applied to radiology report text can identify VTE with high accuracy.

**Clinical Complexity:** *LOW* – For patients who present with relevant symptoms, the imaging needed for diagnosis is commonly ordered and relatively easy to conduct. It is relatively straightforward to determine from this imaging whether VTEs are present or absent. There is no evidence of substantial disagreement between clinicians.

**Data Complexity:** *MEDIUM* – Due to inadequacy of ICD codes in identifying VTE, NLP algorithms are needed to identify VTE from imaging reports. This approach uses free text from radiology reports, which are semi-structured and have relatively standardized language. Separate NLP algorithms would need to be developed for the 2 types of VTE (DVT and PE), which slightly increases data complexity.

**Conclusion / Fitness-for-Purpose Recommendation:** Diagnosing VTE is straightforward from imaging studies so clinical complexity is low, but the need for NLP to extract information from radiology reports makes data complexity medium, resulting in moderate difficulty. ICD codes do not identify VTEs with high enough validity in relatively healthy, general populations for use in post-marketing safety surveillance. Evidence from relatively recent studies in North America shows that several basic NLP algorithms applied to imaging reports have been very successful, demonstrating feasibility of this approach.

**Limitations:** This fitness-for-purpose recommendation is based on literature reviews as of September 2024. We assume an ideal performance equates to a PPV of at least 80%.

#### Detailed Discussion

##### Description of HOI

Venous thromboembolism (VTE) refers to the presence of a blood clot in the veins. The two specific types of VTE are: 1) deep vein thrombosis (DVT), a condition where a blood clot forms in a deep vein, commonly in the upper or lower extremities and 2) pulmonary embolism (PE), when a clot affects arteries in the lungs. Major risk factors for VTE include hospitalization, immobilization, and surgery. VTEs can be detected and treated in the inpatient, emergency department, and outpatient settings.

##### Drug from Drug-Outcome Pair:

1) abrocitinib (Cibinqo), Memo #2021-1436
2) upadacitinib (RINVOQ), Memo #2021-276

**Reason for ARIA insufficiency concern(s) related to outcome, according to FDA memo(s):** Abrocitinib and upadacitinib are Janus kinase inhibitor medications being considered in these memos for treatment of moderate-to-severe-atopic dermatitis, a condition which is most common in youth, but also occurs in adults. Both memos emphasize the importance of sensitivity and specificity in identifying the outcome of VTE. They note that "adequate verification typically requires standardized clinical review of primary patient records" following a "sensitive code search". Although diagnosis-code based algorithms (which would be used in ARIA) may have high enough specificity (and PPV) for VTE in certain clinical settings (e.g., following orthopedic surgery), there is concern that they may not have sufficient specificity and PPV in populations with lower background rates of VTE (e.g., the atopic dermatitis population). There is also no evidence that algorithms have sufficient sensitivity.

##### Previous Validation Studies:

*ICD codes & administrative data*

Because there are many validation studies available, we focus this summary on ICD-10 codes, the version in current use in the US. For algorithms using ICD-10 diagnosis codes to identify VTE with a gold standard of manual chart review, a recent systematic review and meta-analysis found a mean PPV of 74.1%, but PPVs in individual studies were very inconsistent, ranging from 1.3% to 100% (Liu et al., 2024). The mean sensitivity for VTE was 76.5%, but sensitivity in individual studies also had a wide range (20.0% to 100%). For PE specifically, the mean PPV was 66.2% (range: 0.0% to 100%), and the mean sensitivity was 82.0% (range: 0.0% to 100%). For DVT specifically, the mean PPV was 71.6% (range: 1.3% to 100%), and the mean sensitivity was 53.2% (range: 20.0% to 100%). Limitations in the relevance to this computable phenotype feasibility assessment include that several of the included studies were 1) from outside the US, and 2) limited to inpatient VTEs or even post-surgical patients, which tend to have higher PPV than can be expected in a relatively healthy general population. Additionally, although mean PPV and sensitivity were moderate to high, this was not consistent across studies, with many studies having low PPV and sensitivity.

The study most relevant for Sentinel (because it included recent US data from a variety of care settings) is a multisite study using 2016–2020 insurance claims data in a population with rheumatoid arthritis on immunosuppressive medications (Kim et al, 2023). The authors assessed the validity of VTE ICD-10 codes in inpatient, emergency department, and outpatient settings. Their algorithm, including ICD-10 codes for VTE along with a claim for an anticoagulant medication post-VTE, had a PPV of 75.5% (95% CI: 68.7-82.3%), with chart review as the gold standard. The authors did not report sensitivity. Even with the added requirement for an anticoagulant medication claim, the algorithm did not identify VTE with sufficient accuracy. Overall, we found that algorithms using only claims data did not have adequate PPV and sensitivity to identify VTE accurately in populations similar to the atopic dermatitis population (i.e., relatively healthy populations).

*Natural language processing (NLP)*

NLP appears to identify VTE much more accurately than claims data based on consistently high PPV and sensitivity observed across many studies. A recent meta-analysis of NLP approaches including 8 studies found a pooled PPV of 91% (95% CI, 87-94%), with PPVs for individual studies ranging from 78% to 97%. They found a pooled sensitivity of 93% (95% CI, 88-96%), with sensitivities for individual studies ranging from 79% to 98% (Lam et al., 2024). Potential limitations include that this meta-analysis included some studies that only assessed PE, and some studies focused on inpatient cases (some specifically on post-surgical patients). Overall, it appears that NLP algorithms are sufficient for both types of VTE: a study across a few US institutions using data from 2018-2022 found their NLP algorithms to be highly accurate for both DVT (sensitivity: 98.0%, specificity: 99.4%) and PE (sensitivity: 99.5%, specificity: 99.8%) (Scott et al., 2023). A study in a pediatric population in Canada had similarly high validity (Gálvez et al., 2017), suggesting an algorithm in a largely adolescent atopic dermatitis population could also perform well.

**Clinical Complexity:** *LOW – VTEs are either present or absent, which is relatively straightforward to determine from imaging. There is no evidence of substantial difficulty in diagnosing these conditions or disagreement between clinicians.*

There are standard clinical consensus diagnostic guidelines published by the American Society for Hematology (Lim et al, 2018), which are aligned with guidelines by the United Kingdom's National Institute for Health and Care Excellence (2023). These indicate that there is a gold standard diagnostic imaging test for each type of

VTE, specifically duplex ultrasonography for DVT and CT pulmonary angiogram for PE. (A ventilation-perfusion scan will occasionally be used to diagnose PE, but this is less common). For patients who present with relevant symptoms, a diagnostic imaging test is commonly ordered and relatively easy to conduct. Interpreting these images appears to be relatively straightforward, and agreement between different physicians has been reported to be good (Shaham et al., 2006; Schwartz et al., 2002). There is no evidence of competing diagnoses that are particularly challenging to differentiate from VTE.

If some people with VTE have no symptoms or minor symptoms that resolve without treatment, these VTEs will be missed by any approach using routine clinical data. This is not considered a major limitation because these events are not likely to be clinically significant.

**Data Complexity:** *MEDIUM – Due to inadequacy of ICD-10 codes for identifying VTE, NLP algorithms are needed to identify VTE from imaging reports. Imaging reports are semi-structured and use relatively standardized language.*

VTE is technically a composite outcome, consisting of 2 sub-outcomes (DVT and PE). Although composite outcomes often will have increased complexity, here, there is not a large impact on complexity because there are only 2 sub-outcomes, both well-defined, and the data elements required are similar for the two sub-outcomes. Data complexity is slightly increased by needing separate algorithms for the 2 sub-outcomes.

The information about DVT or PE would need to be extracted from reports from imaging studies. For DVT, this would be duplex ultrasonography and for PE it would most commonly be CT pulmonary angiography, but occasionally a ventilation-perfusion scan. The requirement of NLP increases the data complexity. Radiology reports are semi-structured (not unstructured, like most clinical notes). As such, abstracting text from radiology reports is a simpler task for NLP than clinical notes because they have a relatively narrow clinical content, simpler vocabulary, and are more consistently formatted. Supporting that this approach is feasible, several basic NLP algorithms that utilize these reports have been highly successful. Some NLP studies of VTE have posted their programming code on a public repository, such as GitHub (e.g., Verma et al., 2023), which could provide a foundation for future efforts within Sentinel.

**Data Sources:**

| Necessary EHR Data | Useful EHR Data |
|---|---|
| Imaging reports (duplex ultrasound, CT pulmonary angiography, ventilation-perfusion scan) | |

**Conclusion / Fitness-for-Purpose (FFP) Recommendation – *MODERATE DIFFICULTY***

Although clinical complexity is low, with diagnosis being straightforward from imaging studies, data complexity drives the MODERATE rating. Specifically, ICD-10 codes do not consistently identify VTEs with high enough validity in more general populations where VTE incidence is low.

Evidence from recent studies in North America indicates that basic NLP algorithms using imaging reports to identify VTE have been very successful, suggesting that this is likely a feasible approach. However, NLP algorithms take considerable effort to develop and validate, and separate algorithms would need to be developed for DVT and PE, hence the moderate rating.

#### References

Gálvez JA, Pappas JM, Ahumada L, Martin JN, Simpao AF, Rehman MA, Witmer C. The use of natural language processing on pediatric diagnostic radiology reports in the electronic health record to identify deep venous thrombosis in children. J Thromb Thrombolysis. 2017 Oct;44(3):281-290. doi: 10.1007/s11239-017-1532-y. PMID: 28815363.

Lam, B. D. et al. (2024) 'Machine learning natural language processing for identifying venous thromboembolism: systematic review and meta-analysis', Blood advances. American Society of Hematology, 8(12), pp. 2991–3000.

Liu B, Hadzi-Tosev M, Kerolos Eisa, Liu Y, Lucier KJ, Garg A, Li S, Xu E, Mithoowani S, Ikesaka R, Heddle NM, Rochwerg B, Ning S, Accuracy of venous thromboembolism ICD-10 codes: A systematic review and meta-analysis, Thrombosis Update, Volume 14, 2024, pp. 2666-5727, doi: 10.1016/j.tru.2023.100154.

Kim S, Martin C, White J, Carlyle M, Bui B, Gao S, Salinas CA. Validation of an Algorithm to Identify Venous Thromboembolism in Health Insurance Claims Data Among Patients with Rheumatoid Arthritis. Clin Epidemiol. 2023 Jun 1;15:671-682. doi: 10.2147/CLEP.S402360. PMID: 37284517; PMCID: PMC10241253.

Schwarz T, Schmidt B, Schmidt B, Schellong SM. Interobserver agreement of complete compression ultrasound for clinically suspected deep vein thrombosis. Clin Appl Thromb Hemost. 2002 Jan;8(1):45-9. doi: 10.1177/107602960200800106. PMID: 11991239.

Scott, D. M. et al. (2023) 'Natural Language Processing tool accurately identifies acute venous thromboembolism', Thrombosis research. Elsevier BV, 229, pp. 252–254.

Shaham D, Heffez R, Bogot NR, Libson E, Brezis M. CT pulmonary angiography for the detection of pulmonary embolism: interobserver agreement between on-call radiology residents and specialists (CTPA interobserver agreement). Clin Imaging. 2006 Jul-Aug;30(4):266-70. doi: 10.1016/j.clinimag.2006.01.001. PMID: 16814143.

Venous thromboembolic diseases: diagnosis, management and thrombophilia testing. London: National Institute for Health and Care Excellence (NICE); 2023 Aug 2. PMID: 32374563.

Verma AA, Masoom H, Pou-Prom C, Shin S, Guerzhoy M, Fralick M, Mamdani M, Razak F. Developing and validating natural language processing algorithms for radiology reports compared to ICD-10 codes for identifying venous thromboembolism in hospitalized medical patients. Thromb Res. 2022 Jan;209:51-58. doi: 10.1016/j.thromres.2021.11.020. Epub 2021 Nov 27. PMID: 34871982.

### Appendix 1: Society Guidelines for Venous Thromboembolism in the United States

Guidelines for the diagnosis and management of pericardial effusion are issued by the American College of Cardiology (ACC) who recommend following the guidance of the European Society of Cardiology (ESC).

| Source | Definition |
|---|---|
| American Society of Hematology 2018<br><br>(Lim et al. 2018) | Imaging is the gold standard for diagnosis, specifically:<br>• DVT: duplex ultrasonography<br>• PE: computed tomography pulmonary angiography or ventilation-perfusion scanning |

#### Appendix 2: ICD Diagnostic Codes Relevant to Venous Thromboembolism

This list of ICD-10 codes is not intended to be exhaustive but rather provided for reference purposes.

ICD-10

| Code | Description | Source |
|---|---|---|
| I26.0, I26.02, I26.09, I26.9, I26.92, I26.99 | PE | Kim *et al*., 2023 |
| I80.10, I80.11, I80.12, I80.13, I80.201, I80.201, I80.203, I80.209, I80.211, I80.212, I80.213, I80.219, I80.221, I80.222, I80.223, I80.229, I80.231, I80.232, I80.233, I80.239, I80.291, I80.292, I80.293, I80.299, I80.3, I82.401, I82.402, I82.403, I82.409, I82.411, I82.412, I82.413, I82.419, I82.421, I82.422, I82.423, I82.429, I82.431, I82.432, I82.433, I82.439, I82.441, I82.442, I82.443, I82.449, I82.491, I82.492, I82.493, I82.499, I82.4y1, I82.4y2, I82.4y3, I82.4y9, I82.4z1, I82.4z2, I82.4z3, I82.4z9 | Lower extremity DVT | Kim *et al*., 2023 |
| I82.210, I82.290, I82.601, I82.602, I82.603, I82.609, I82.621, I82.622, I82.623, I82.629, I82.a11, I82.a12, I82.a13, I82.a19, I82.b11, I82.b12, I82.b13, I82.b19, I82.c11, I82.c12, I82.c13, I82.c19 | Upper extremity DVT | Kim *et al*., 2023 |
| I80.8, I80.9, I81, I82.0, I82.1, I82.220, 182.3, I82.890, I82.90 | Other VT | Kim *et al*., 2023 |

### Fitness-for-Purpose (FFP) Assessment: Major Bleed

| | |
|---|---|
| **Health Outcome of Interest:** | Major Bleed (composite outcome) |
| **Date:** | February 11, 2025 |
| **Subject:** | Health Outcome of Interest Feasibility Assessment for HOI |
| **Drug(s)/ ARIA Insufficiency Memo(s):** | 1. PRADAXA (dabigatran etexilate), Memo #2020-1989 |

### Executive Summary

**Overall Assessment:** *MODERATE* – Major bleed is a composite HOI. ICD codes may be sufficient for identifying some but not all of the component HOIs, requiring that other approaches and data be used, including NLP applied to various types of EHR data. If fatal hemorrhage including out of hospital deaths is essential to the algorithm, the difficulty increases to HARD.

**ARIA Insufficiency Issue:** Claims based algorithms are available for major bleed in adults but may not be sufficient. Results of laboratory testing that are needed for this outcome, including blood coagulation tests, are not currently available in Sentinel ARIA.

**Orientation to HOI**: Major bleed is defined as follows: (1) an intracranial hemorrhage (a bleed within the brain) including those from closed head trauma and irrespective of fatality status OR (2) an extracranial hemorrhage a) requiring red blood cell or whole blood transfusion, or b) involving critical anatomical sites, specifically, intraarticular (in a joint space), pericardial (around the heart) or retroperitoneal (abdominal), or c) that was fatal (resulting in death within 30 days of the bleeding event).

**Existing Computable Algorithms:** The use of ICD codes may be sufficient for identifying intracranial hemorrhage, but other component HOIs appear to be more difficult. Reported definitions for other bleeding events, including gastrointestinal, urogenital, and "other", generally show poor positive predictive value (PPV) using ICD codes, even when combined with lab data. In developing a computable phenotype, prior studies utilizing NLP suggest that information could be gleaned from clinical notes, but this too could prove challenging.

**Clinical Complexity:** *LOW/MEDIUM* – Some component HOIs are straightforward to diagnose (extracranial hemorrhage requiring transfusion, fatal bleed) and/or have society guidelines available with a gold standard and high agreement between clinicians (intracranial hemorrhage), while other components do not (intraarticular hemorrhage).

**Data Complexity:** *MEDIUM* – ICD codes are available for intracranial and intraarticular bleeds, and codes for intracranial bleeds have good validity. EHR data needed include multiple types (imaging reports for intracranial hemorrhage; procedure codes and orders data for hemorrhage requiring transfusion; clinical notes and lab reports for intraarticular hemorrhage). Identifying most of the components would require NLP. Because mortality may not be observable in the EHR or recorded in claims data with high enough sensitivity, and national mortality data sources have limitations due to time lags, data complexity would be otherwise high unless fatal bleed can be prioritized lower relative to the other component HOIs.

**Conclusion / Fitness-for-Purpose Recommendation:** *MODERATE* - The major bleed composite HOI is a composite with six component HOIs. There is some but not substantial overlap in the work that would be needed to identify the individual component HOIs. The MODERATE overall assessment was reached only because the fatal hemorrhage component was downgraded in its priority for inclusion in the computable phenotype owing to the difficulty in ascertaining it.

**Limitations:** This fitness-for-purpose recommendation is based on literature reviews as of December 2024. We assume an ideal performance equates to a PPV of at least 80%.

#### Detailed Discussion

**Reason for ARIA insufficiency concern(s) related to outcome, according to FDA memo(s):** Claims-based algorithms are available for major bleed in adults, but have low accuracy, especially low PPV. No validated algorithms in pediatric patients are available based on a search of published medical literature. Without validation the performance of any algorithm is unknown. Results of clinical laboratory testing used to identify this outcome, including blood coagulation tests, are not currently available in Sentinel ARIA.

**Drug from Drug-Outcome Pair:**

1. PRADAXA (dabigatran etexilate), Memo # 2020-1989

**Orientation to HOI:**

There is substantial heterogeneity in the medical literature as to how occurrences of "major bleed" are classified (See Appendix 1 and Mehran et al. 2011; Wells et al. 2019). Based on consultation with the FDA, this memo will focus on a composite definition of major bleed as follows: (1) an intracranial hemorrhage (a bleed within the brain) including those from closed head trauma and irrespective of fatality status OR (2) an extracranial hemorrhage a) requiring red blood cell or whole blood transfusion, or b) involving critical anatomical sites, specifically, intraarticular (in a joint cavity), pericardial (around the heart) or retroperitoneal (abdominal), or c) that was fatal (resulting in death within 30 days of the bleeding event) (Figure 1). Because the definition of major bleed represents a composite outcome, i.e., a group of conditions, these components will need to be evaluated individually to assess their clinical and data complexity.

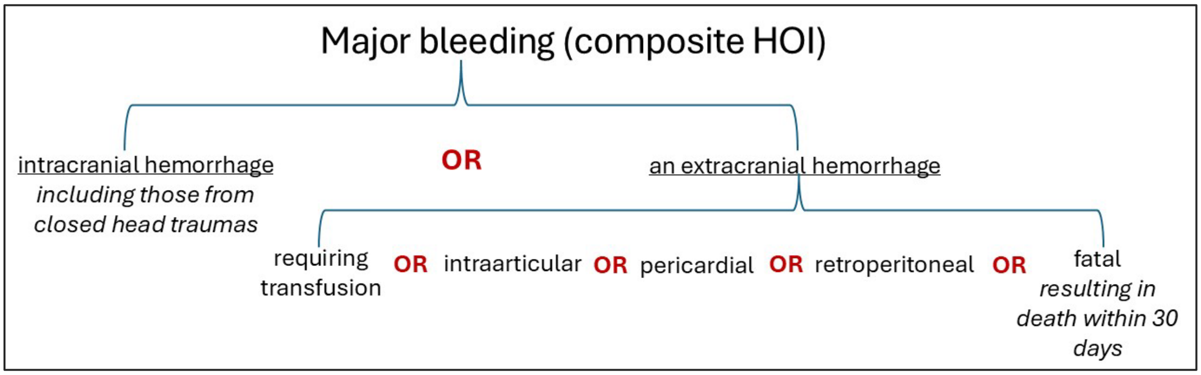

***Figure 1. Schematic of Major Bleed Composite HOI with Components.***

Based on discussion with the FDA and due to time constraints, we focus our assessments of the following individual component HOIs which are described in detail in sections below pertaining to the components:

- Intracranial hemorrhage (see Component 1)
- Extracranial hemorrhage requiring red blood cell or whole blood transfusion (see Component 2)
- Intraarticular hemorrhage (see Component 3)
- Fatal hemorrhage (see Component 4)

For the major bleed composite HOI to be complete, evaluations of pericardial and retroperitoneal bleeds should be conducted in the future and integrated in this FFP assessment.

**Literature Review of Previous Validation Studies/Algorithms:**

In general, validation studies of ICD-9 or ICD-10 codes for bleeds at anatomical sites relevant to the major bleed definition have yielded variable levels of performance. The following studies focus on identifying broad, composite definitions of bleeding. Validation studies specific to a component HOI are presented in the relevant section.

- Joos (2019) reviewed notes from anticoagulated patients at the University of Utah hospital (n=487) and found high sensitivity (91.4%) but low PPV (52.5%) for ICD-10 codes for a composite outcome of "bleeding events", encompassing gastrointestinal bleeding, intracranial hemorrhage, hematoma, genitourinary bleeding, epistaxis, and retroperitoneal bleeding, among others.
- Oger (2019) developed a similar ICD-10 algorithm for "major bleeding" (grouped into intracranial hemorrhage, gastrointestinal hemorrhage, and other miscellaneous bleeds) and applied it to admissions at a French hospital in 2014 (16,012 hospital stays). They used an automated, previously-validated approach (sensitivity of 96%) using diagnostic codes and emergency therapies. The goal of their work was to eliminate hospital stays that were likely to be unrelated to bleeding events. Applying this algorithm generated a dataset of 1,959 hospitalizations which was then manually reviewed by a medical expert who classified 736 as major bleed events. On the assumption that all stays apart from these 736 were not major bleed events, the study reported an overall sensitivity of 65%.
- Ruigomez (2018) looked at a primary care database including Swedish, Spanish, and German populations aged 2 to 89 years who had received an anticoagulant prescription. Using manual review of patient notes, they similarly found that ICD-10 codes relevant to major bleed had PPVs of 96.9% (intracranial) and 70.5% (gastrointestinal). Sensitivity was not reported.
- Yap (2024) led a study exploring the utility of using ICD-10 codes in combination with laboratory data to identify major bleed events (as defined by the International Society on Thrombosis and Haemostasis [Schulman 2005]) in an inpatient setting in Singapore. Using a validation set of 1000 patients, the researchers analyzed three separate algorithms: 1) ICD-10 codes

only (mapped from 189 Snomed CT codes); 2) a "sensitivity-optimized" algorithm (>= 3 hemoglobin [Hgb] lab tests in 7 days with adjacent tests showing rise/fall in Hgb levels by >= 1g/dL); 3) a "PPV-optimized" algorithm (same as "sensitivity-optimized", but with additional requirement of >= 2g/dL rise/fall between any of the tests in the 7-day window). PPV and sensitivity for the algorithms were: 1) ICD-10: 1.00, 0.16; 2) Sensitivity-optimized: 0.34, 0.94; 3) PPV-optimized: 0.52, 0.88. Note that PPV was fairly low even for the "PPV-optimized" version of the algorithm.

Natural language processing (NLP) has not been applied specifically to the outcome of "major bleed", though it has been applied to other bleeding definitions with some success.

- Taggart (2018) developed both rule-based and machine learning models to identify "clinically relevant bleeding event[s]" during hospitalization. The concept of "clinically relevant" was not defined in the manuscript. Using a training set of 990 notes and test set of 660 notes, performance for the rule-based approach included a PPV of 0.627 and a Sensitivity of 0.911. The best machine learning model had a PPV of 0.707 and a Sensitivity of 0.712.
- Li (2019) trained a series of machine learning models to detect "important bleeding events" in clinical narratives. "Important bleeding events" was not defined. Every note was annotated by two physicians (Cohen's kappa: 0.9182). Classification occurred at the sentence level using 878 clinical notes. PPVs ranged from 0.848-0.954 with sensitivities from 0.855-0.912. They noted that one of the primary challenges in this approach was identifying statements that bleeding was not present.

While these validation studies suggest that the use of ICD codes may be sufficient for identifying intracranial hemorrhage, other component HOIs appear to be more difficult. Reported definitions for other types of bleeding events, including gastrointestinal, urogenital, and "other", generally show poor PPV using ICD codes, even when combined with lab data. While these classifications do not map directly to our definition of "major bleed", they hint at the challenge of identifying bleeding events other than intracranial bleeds. In developing a computable phenotype, the NLP studies suggest there is some information that could be gleaned from clinical notes, but this too could prove challenging.

**Data Elements:**

| Necessary EHR Data | Useful EHR Data |
|---|---|
| Imaging reports | Inpatient nursing notes |
| Diagnosis codes | Imaging reports |
| Procedure codes | Laboratory data |

##### Clinical Complexity:

Major bleed is a composite outcome in which meeting any one of the multiple criteria (i.e., four component outcomes) fulfills the definition for this HOI. Assessing clinical complexity begins with disaggregating the composite into its components (intracranial hemorrhage; hemorrhage requiring red blood cell or whole blood transfusion; intraarticular hemorrhage; pericardial hemorrhage; retroperitoneal hemorrhage; or fatal hemorrhage). Overall clinical complexity is determined by the clinical complexity of the individual components. See Table 1 below. The reasoning behind the assessments for each type of bleeding is presented in the relevant sections of the report which follow this summary.

##### Data Complexity:

As with clinical complexity, overall data complexity is determined by the data complexity of the individual components. See Table 1 below.

***Table 1. Clinical and Data Complexity Summary Table.***

| Component HOI | Clinical Complexity | Data Complexity | Overall |
|---|---|---|---|
| Intracranial hemorrhage | Low | Low/Medium | Moderate |
| Hemorrhage requiring transfusion | Low | Medium | Moderate |
| Intraarticular hemorrhage | Medium | Medium | Moderate |
| Fatal hemorrhage | Low | High | Hard |

##### Conclusion / Fitness-for-Purpose (FFP) Recommendation:

Because the major bleed composite HOI can be met by any one of the component HOIs, the overall difficulty for developing a computable phenotype could be determined by the most difficult component. In this case, the major bleed composite that includes fatal hemorrhage would receive a HARD overall assessment (Table1) because the fatal hemorrhage component would be HARD to create a phenotype.

If it is satisfactory to accept imperfect identification of deaths (focusing on those available from the initial hospitalization for the bleed, or at a rehospitalization within the same healthcare system), then the assessment for the fatal hemorrhage component may be downgraded to MODERATE difficulty and the overall assessment of major bleed would be MODERATE.

Alternatively, if the FDA prioritizes the component HOIs such that one of the other components is the most important to phenotype (and fatal hemorrhage is less important to phenotype), then the major bleed composite could receive a MODERATE overall assessment. This choice might be reasonable if one of the other components is likely to be much more common and to account for a larger proportion of the major bleed events.

#### Component 1: Assessment of Intracranial Hemorrhage

**Description of HOI:**

Intracranial hemorrhage (a bleed in the brain) is a composite outcome comprised of four types of hemorrhage which are identified based on their anatomical location within the brain (Tenny and Thorell, 2024):

- epidural hemorrhage,
- subdural hemorrhage [(SDH), also referred to as a subdural hematoma],
- subarachnoid hemorrhage (SAH), and
- intraparenchymal hemorrhage [also referred to as an intracerebral hemorrhage (ICH)].

Epidural and subdural hemorrhage are most commonly associated with a head injury, while a subarachnoid hemorrhage can result from both traumatic and non-traumatic events; an example of a non-traumatic cause would be the rupture of an aneurysm. An intraparenchymal hemorrhage can result from a number of causes including hypertension, arteriovenous malformation (AVM), amyloid angiopathy, aneurysm rupture, tumor, coagulopathy, infection and vasculitis in addition to trauma. Non-traumatic ICH comprises 10-15% of all strokes (Rajashekar and Liang, 2023).

**Previous Validation Studies:**

- Overall, validation studies indicate ICD-10 codes are able to identify intracranial hemorrhage with relatively high accuracy. However, only two US-based studies were identified. Differences between the US health system and those of the non-US studies introduce questions about the extent these studies will generalize to the US. Furthermore, all studies stem from the ICD-10 era and it is therefore not known how ICD-9 codes would perform. Columbo et al. (2024) assessed the accuracy of ICD-10 codes using data from two prospective population-based cohort studies [the ARIC Study (4 US communities) and the REGARDS Study (national sample from the US)]. Participants in both studies were aged 65 years or older without prior stroke who had linked Medicare claims data. Diagnostic codes were compared against gold standard incident stroke events (n=110) identified via active surveillance and adjudicated by expert review. For nontraumatic subarachnoid hemorrhage and nontraumatic intracerebral hemorrhage using data from the first study: PPV: 65%, sensitivity (SE): 82% (primary diagnosis code), PPV: 57%, SE: 95% (any diagnosis code); using data from the second study: PPV: 65%, SE: 71% (primary diagnosis code), PPV: 62%, SE: 78% (any diagnosis code).
- Shehab et al. (2018) assessed performance of ICD-10 codes for identifying bleeding events resulting in emergency department (ED) visits and hospitalizations among outpatient Medicare beneficiaries prescribed anticoagulants between October 1, 2015 and September 30, 2016 (study sample of 1166 records). Structured medical record review was the gold standard for validating the presence of anticoagulant-related bleeding. Among ED visit cases, four codes for intracranial bleeding each individually

Sentinel

demonstrated 100% PPV. Grouped together, the performance of the intracranial bleeding codes varied by code position: codes in any position had PPV 85%, sensitivity 91%, while codes in primary position had PPV 91%, sensitivity 71%.

- Hsieh et al. 2021 evaluated ICD-10 codes for identifying subarachnoid hemorrhage (SAH) or intracerebral hemorrhage (ICH) within Taiwan's National Health Insurance claims database among patients hospitalized for stroke at two hospitals (n=806) using stroke registry databases as the gold standard. SAH in any diagnostic field had a PPV = 88% and SE = 95%; ICH in any diagnostic field had a PPV = 91% and SE = 99%; PPV increased for both diagnostic codes if diagnostic field position was limited to primary.
- Hald et al. 2018 assessed the validity of intracerebral hemorrhage (ICH) diagnoses (ICD-10 era) among a sample of 500 patients from the Danish National Patient Registry during 2010–2015, using discharge summaries and brain imaging reports as the gold standard, and reported an PPV = 88%.

**Clinical Complexity:** *LOW – society guidelines are available and in agreement, diagnosis is based on a gold standard test (imaging), and diagnostic agreement between clinicians is good.*

Consensus guidelines for subdural hematoma in the US are available from ARISE (Aneurysm/AVM/cSDH Roundtable Discussion With Industry and Stroke Experts) (Kap et al. 2024). A computed tomography (CT) scan is considered the standard for diagnosis.

Diagnostic guidelines for intracerebral hemorrhage in the US are available from the American Heart Association/American Stroke Association (Greenberg et al. 2022). Neuroimaging (CT or MRI) is required for a definitive diagnosis (Rordorf and McDonald, 2024).

Studies show generally good levels of agreement between clinicians diagnosing a brain bleed. A Dutch study showed good interobserver agreement of the presence of any ICH on MRI between six neuroradiologists [kappa = 0.78 (95% CI, 0.71–0.85)] (Van der Ende et al. 2023). In a US study, agreement was excellent between physicians who reviewed medical records and all other available information to diagnose SAH (kappa = 0.82) and ICH (kappa = 0.96) (Berger et al 1996). However, a British study found poor overall agreement for an ICH diagnosis on CT (kappa=0.35) (Lovelock et al. 2009).

Because the HOI was defined to include intracranial hemorrhages of both traumatic and non-traumatic etiologies, there is not a need to try to identify whether hemorrhages resulted from trauma (which would increase the clinical complexity).

**Data Complexity:** *LOW/MEDIUM – ICD codes are available and have good validity; EHR data is limited to one type, brain imaging, but would require NLP.*

ICD-10 codes for subdural hematomas and intracerebral hemorrhages have been demonstrated to have good to very good PPV and moderate to high sensitivity depending on the way they are defined. Some studies show slightly lower PPV for codes in any position (not limited to primary). Subdural hematomas and intracerebral hemorrhages are identified clinically by neuroimaging, either brain magnetic resonance imaging (MRI) or computed tomography (CT) (McBride, 2024; Grossman, 2003). Data from imaging reports are semi-structured, which is more complex to extract and use than structured laboratory test results. Natural language processing (NLP) would be required to extract information from imaging records to identify SDH and ICH, which increases data complexity.

**Data Sources:**

| Necessary EHR Data | Useful EHR Data |
|---|---|
| Imaging reports | |
| Diagnostic codes | |

**Conclusion/Fitness-for-Purpose Recommendation:** *MODERATE*

Development of a computable algorithm for intracranial bleeding should be moderately difficult based on low clinical complexity and low to moderate data complexity. This HOI benefits from ICD-10 codes with good validity, available consensus guidelines, a clear “gold standard” for diagnosis, and evidence of good agreement between clinicians. If ICD codes alone are deemed to have sufficient accuracy, overall difficulty would be EASY. If their accuracy is felt to be not sufficiently high, then NLP of imaging reports would be involved, resulting in difficulty of MODERATE. Additional studies in the future providing improved algorithms could change this determination.

#### Component 2: Assessment of Extracranial Hemorrhage Requiring Red Blood Cell or Whole Blood Transfusion

**Description of HOI:**

An extracranial hemorrhage requiring transfusion of red blood cells or whole blood is considered a major bleed. Guidelines about when to administer a transfusion have varied over time and according to presence of clinical conditions; recent guidelines suggest that in general, a transfusion is indicated when hemoglobin levels drop to less than 6–8 g/dL (Lotterman & Sharma, 2024; Szczepiorkowski & Dunbar, 2013). There is variability across different medical society definitions in terms of how much blood or blood products must be transfused to qualify as a major bleed, with the Bleeding Academic Research Consortium (BARC) allowing any amount of transfusion to count (Mehran et al., 2011), and the International Society on Thrombosis and Haemostasis (ISTH) requiring ≥2 units packed red blood cells (Schulman et al., 2005). For this FFP assessment, we are focusing on any transfusion of red blood cells or whole blood, other than exchange transfusions. We anticipate that most transfusions due to hemorrhage would occur in the inpatient setting, but some could also occur in the outpatient setting, most likely in the emergency department.

In assessing whether this HOI is FFP, we are focusing on red blood cell or whole blood transfusions used to treat an extracranial hemorrhage. Intracranial hemorrhages are not treated with transfusions; in an intracranial hemorrhage, blood is trapped in the intracranial space, so the problem is not loss of blood from the body, but rather pressure on the brain. Therefore, hemorrhages treated with transfusions are inherently extracranial.

**Previous Validation Studies:**

We did not find studies validating algorithms specifically identifying any extracranial hemorrhage *and* a transfusion of red blood cells or whole blood. Therefore, we are assessing the hemorrhage and transfusion pieces separately, recognizing that an ideal study would assess them jointly.

Considering analyses validating diagnostic codes for extracranial hemorrhage without requiring transfusion, studies have focused on gastrointestinal hemorrhage, the most common type of extracranial hemorrhage. Al-Ani and colleagues (2015) found a PPV of 74% (95% CI: 60–85%) for upper gastrointestinal bleeds and 90% (95% CI: 78–96%) for lower gastrointestinal bleeds in an emergency room setting in Canada. One study did include both hemorrhage and transfusion, but only added transfusion in an attempt to boost the performance of their algorithm (Park et al., 2019). The authors identified 200 patients with gastrointestinal bleeding (most common type of extracranial hemorrhage) from a single hospital in Korea. Requiring a hospitalization and a primary hemorrhage diagnostic code resulted in a PPV of 92%; adding a requirement of red blood cell transfusion also resulted in a PPV of 92%, compared with a gold standard of gastrointestinal bleed (with or without transfusion). We are not able to infer what the validity of an algorithm for extracranial hemorrhage requiring transfusion would be, since the study did not require a transfusion as part of their gold standard comparison. Overall, the PPVs for identifying gastrointestinal hemorrhage look relatively promising, and while they may not generalize to all extracranial hemorrhages requiring transfusion, we believe that a hemorrhage code along with evidence of a transfusion is likely to represent a true hemorrhage.

Regarding validity of transfusion identification, the most recent relevant study was conducted in the Veterans Administration (VA) in the ICD-9 era (2008-2013), and it assessed the performance of multiple algorithms in identifying red blood cell transfusion-related hospital admissions for anemia associated with end-stage renal disease (ESRD) using VA data linked with Medicare data (Peters et al., 2019). All included individuals had ESRD, and those with alternative causes of anemia (such as bleeding, malignancy, surgery, or hematological disorders) were excluded. Chart review was the gold standard. Findings from a few of their algorithms are described below:

- Clinical algorithm (EHR data)
  - Required a hospital admission with evidence of anemia (hemoglobin lab result <9.0 g/dL) AND evidence of a red blood cell transfusion based on orders, laboratory data, and procedure codes occurring after the hemoglobin measurement.
  - Found a PPV of 72.1% (95% CI: 66-77.4%) and sensitivity of 83.5% (67.9–92.4%).
- Claims-based algorithm 1
  - Used insurance claims, specifically, required a hospital admission with a principal discharge diagnosis of chronic kidney disease (CKD) or ESRD, a secondary diagnosis of anemia, AND a procedure code for an RBC transfusion.
  - Found a PPV of 80% (95% CI: 63.9-90.1%) and sensitivity of 38.3% (29.3-48.3%).
- Claims-based algorithm 2
  - Used insurance claims, specifically required a hospital admission with any discharge diagnosis for CKD or ESRD, any discharge diagnosis for anemia, AND a procedure code for an RBC transfusion.
  - Found a PPV of 66.1% (56.4–74.5%) and sensitivity of 80.6% (71.7–87.2%).

Most of the false positives, which lowered PPV, were due to patients who had an alternative etiology for their anemia (e.g., bleeding) noted on chart review but lacked a code for this and so were included in the population even though they did not actually have ESRD-associated anemia. Low sensitivity in claims-based algorithm 1 was more often a result of missing CKD/ESRD or anemia codes, not missed transfusions. However, some data sources that were used, such as an accounting-related source recording laboratory data that was used to identify transfusions, may be unique to the VA and not replicable elsewhere.

Overall, this study had promising results for identifying transfusions, especially given that the factors lowering their performance measures appeared to be related to identifying ESRD as the cause of the transfusion, which is not relevant to this HOI. Considering both PPV and sensitivity, the clinical algorithm (using EHR data) had the best performance.

Two studies using data from the late 1990's assessed the validity of procedure codes for transfusion at single hospitals. The study in a single Canadian hospital used chart review as the gold standard and found a sensitivity of 26.4%, a specificity of 99.7%, and a PPV of 91.7% (Quan et al., 2004). The study in a single US hospital using blood bank records as the gold standard found a sensitivity of 83% and specificity of 100% (therefore, PPV=100%) (Segal et al., 2001). Except for the low sensitivity in the Canadian study, these findings are generally promising; however, these single-center studies from 25+ years ago may not be relevant to modern practice.

**Clinical Complexity:** *LOW* – Extracranial hemorrhage requiring transfusion can be diagnosed relatively easily. A transfusion is indicated in patients with active or acute bleeding or a recent bleed with hemoglobin less than a threshold (often 6-8 g/dL) (Lotterman & Sharma, 2024). Assessing clinical complexity for transfusions is unusual because these are procedures, not a condition or health state that applies to the patient. However, there is little ambiguity in the clinical setting as to whether someone received a transfusion and what was transfused (i.e., whole blood, red cells, platelets, plasma, etc.), so we are rating this as low clinical complexity.

**Data Complexity:** *MEDIUM* – Both the extracranial hemorrhage and the transfusion would need to be identified to meet the definition of this HOI subcomponent.

Compiling a list of codes for extracranial hemorrhage would be relatively straightforward; the Al-Ani et al. and Park et al. studies included code lists for gastrointestinal hemorrhage, and other studies have compiled code lists relevant to extracranial hemorrhage, but did not conduct validation on these code lists (Hartenstein et al., 2023 [most relevant codes are overt bleeding codes, included in Appendix 2 of this report]; de Burgos-Gonzalez et al., 2021).

Regarding identifying transfusions, it is possible that procedure codes from insurance claims would be sufficient to identify transfusions. In the inpatient setting, most procedures are bundled, making it difficult to identify individual aspects of care, but we found evidence that blood products are billed separately, such that they could be identified via claims (Peters et al., 2019). We also conducted an investigation assessing the ability to identify transfusions in a single, integrated health system that does not have hospitals and found that we were able to identify a substantial number of transfusions, both outpatient and inpatient, via codes (CPT, ICD diagnostic and procedure codes, HCPCS, revenue codes). This brief investigation indicated that it is possible to identify at least some transfusions via codes on billing claims for inpatient stays, as well as outpatient encounters.

However, the study by Peters et al. (2019), described above, indicated that to achieve relatively high PPV *and* sensitivity, the clinical algorithm using EHR data was needed. Adding support to this finding, the study by Quan et al. (2004), which only used procedure codes, found that PPV was high, but sensitivity was very low. In the inpatient setting, a transfusion may be captured in the EHR via blood products being ordered (recorded in structured order fields) or via procedure codes for transfusion of whole blood or red blood cells (Menis et al., 2009; Kleinman et al., 2013; Peters et al., 2019). How the orders for blood products are entered or stored may vary across different hospitals and/or EHR systems, which could add to complexity of data extraction. To increase accuracy of an algorithm, it would be advisable to include codes from claims, as well as information from EHR data (i.e., orders). We anticipate that extracranial hemorrhage would be more easily identifiable than ESRD, but it is possible that it would be similar or more difficult, which could shift the performance of algorithms.

It is unlikely that clinical notes, such as hospital nursing notes, would need to be queried with NLP for identification of transfusion. A previous study used NLP to identify that transfusions are often recorded in free text nursing notes, but did not assess their NLP system against a gold standard (Hyun et al., 2009).

**Data Sources:**

| Necessary EHR Data | Useful EHR Data |
|---|---|
| Procedure codes | Inpatient nursing notes |
| Diagnosis codes | |
| Orders data (inpatient and outpatient) | |

**Conclusion/Fitness-for-Purpose Recommendation**: *MODERATE* – The difficulty is elevated to MODERATE due to the medium data complexity combined with low clinical complexity. The main drivers of the medium data complexity are the variety of types of inpatient (and outpatient) EHR data likely needed, and the potential need for some NLP.

#### Component 3: Assessment of Intraarticular Hemorrhage

**Description of HOI:**

Intraarticular hemorrhage (also referred to as hemarthrosis) consists of bleeding into a joint such as a knee, elbow, ankle, hip or shoulder. Intraarticular hemorrhages most commonly occur after an injury. They are also associated with bleeding disorders such as hemophilia. They can occur post-operatively such as following a knee replacement surgery or less commonly, can be seen in association with an infection.

**Previous Validation Studies:**

As of the date of this report (December 2024), no validation studies of diagnostic codes for joint bleeds are available.

**Clinical Complexity:** *MEDIUM - A diagnostic gold standard for exists, but in practice, there is not consistency between clinicians in the approach to diagnosis.*

Consensus diagnostic guidelines for diagnosing intraarticular hemorrhage were not identified. While not specified in a consensus guideline, the literature identifies arthrocentesis (joint aspiration) with examination of the synovial fluid as the standard for diagnosis of an intraarticular hemorrhage. While synovial fluid can be sent to the laboratory for formal analysis, clinicians agreed that for significant bleeds, it is obvious from visual inspection of the fluid that a bleed has occurred. Thus, a diagnosis of a joint bleed is very straightforward when results from joint aspiration are available.

Imaging modalities such as radiography, computed tomography (CT), ultrasound or magnetic resonance imaging (MRI) may also provide useful information (Lombardi and Cardenas, 2023). In some cases when joint aspiration is used to manage a swollen joint, i.e., to drain fluid and relieve swelling or because of infection, the therapy may confirm a diagnosis of an intraarticular hemorrhage, i.e., by finding blood in the joint.

It was noted that the approach to diagnosis may vary for different joints, and different joints may be encountered or managed by different physician specialties in different settings (e.g., orthopedists vs. Emergency Department physicians.) To further assess clinical complexity, we queried five medical doctors from Kaiser Permanente who encounter joint bleeds in their practices including in urgent care (UC) and orthopedics. Based on these interviews, we conclude that there is not consistency between clinicians in the approach to diagnosing hemarthroses. We did not find studies reporting on agreement between physicians who received the same information about a patient. Clinical complexity could be increased by the need to integrate data from different sources if a physician does not order a joint aspiration. However, a joint aspiration is commonly ordered.

Hemarthrosis may be present as an associated finding in the context of higher priority clinical conditions, such as a knee injury (e.g., ligamentous tear) or postoperatively. In those cases, the biggest clinical concern is not the bleeding within the joint, and it is likely that a small hemarthrosis will not be formally diagnosed, because it is not considered clinically significant, and the main focus is on managing the primary condition. This may add some clinical complexity in that it is possible for a small hemarthrosis to coexist with the primary condition; the hemarthrosis is not considered clinically meaningful and not recorded consistently in the EHR. Depending on how a computable phenotype is developed, it is possible that these clinically insignificant hemarthroses may be captured in some cases, making the phenotype outcome more “noisy” and less meaningful.

For this HOI, there is not a need to identify and exclude other competing diagnoses. Ambiguity related to competing diagnoses could be resolved by ordering a joint aspirate.

**Data Complexity:** *MEDIUM – multiple data elements are needed, including both structured and unstructured data; some NLP of EHR data is anticipated.*

ICD-10 codes are available for hemarthrosis, but their validity is not well established.

Because joint bleeds will be encountered in emergency medicine, urgent care, primary care and specialty care (orthopedics), data required from EHR would need to be inclusive of ED and outpatient visits, as well as records from more than one specialty.

Several different types of data may provide evidence of a joint bleed. Laboratory results from analysis of joint fluid could provide valuable evidence. Because some physicians diagnose a joint bleed by visual inspection of the joint aspirate, without formal laboratory analysis, key evidence may also be found in free text chart notes. Missing data may be an issue, i.e., if a joint aspirate analysis is not ordered.

Considering laboratory tests, it is unclear how such results would be formatted (i.e., as structured data vs. unstructured data), and so obtaining results might require NLP. Imaging reports from MRI and CT scans or, in some cases, X-rays may also be of value.

Ascertaining this outcome does not require data from multiple time points. The interpretation of the lab test (when it is present) appears to be straightforward (i.e., the presence or absence of blood in joint fluid).

**Data Sources:**

| Necessary EHR Data | Useful EHR Data |
|---|---|
| Clinical notes and notes from specialists (procedure notes, etc.) | Imaging reports |
| Lab reports | |

**Conclusion/Fitness-for-Purpose Recommendation:** *MODERATE*

Since no validation study of diagnostic codes for joint bleeds is available, we recommend conducting a more comprehensive validation study of hemarthrosis diagnostic codes as a first step before further consideration of the development of a computable phenotype. If diagnostic codes alone are sufficient, it could lead to a final assessment of EASY for the computable phenotype for hemarthrosis. Without knowing how well diagnostic codes perform, we would have to assess this HOI as having MODERATE difficulty for fitness for purpose for computable phenotype development because of medium levels of difficulty for both clinical and data complexity.

#### Component 4: Assessment of Fatal Hemorrhage

**Description of HOI:**

A fatal bleed is a significant loss of blood from an anatomical site to the extent that death results. Existing definitions differ in their criteria, including whether they require that bleeding was a direct or indirect cause of death as well as when death occurs in relation to the blood loss (Schenker et al. 2023). For example, some clinical trials have specified a timeframe of 30 days from the bleeding event (Xu et al. 2022) and others 7 days (Thrombolysis in Myocardial Infarction [TIMI]). The BARC defines fatal bleed as bleeding that directly causes death with no other explainable cause. Bleeding sites with potential to cause death include intracranial, gastrointestinal, retroperitoneal, pulmonary, pericardial, genitourinary, or other, and the time interval from the bleeding event to death is considered with respect to likely causality but with no specific time limit required. Using the BARC definition, an example of a fatal bleed would be a patient who receives thrombolytic therapy (a medication to dissolve a blood clot) for a myocardial infarction who later loses consciousness and dies with an autopsy showing a large intracranial hemorrhage.

For the purpose of this assessment, we focus on the feasibility and comprehensiveness of obtaining information about death from claims and/or EHR data, rather than the clinical diagnosis of death.

**Previous Validation Studies:** N/A

Because there are no ICD-9 or 10 codes for fatal bleeds specifically, this section is not applicable.

**Clinical Complexity:** *LOW – mortality is an objectively defined endpoint.*

Death itself is an unambiguous outcome. Diagnosis of a bleed preceding death may require patient information from more than one point in time and/or more than one healthcare encounter. Timing of the bleeding event with respect to death may introduce some ambiguity if it is not specified in the definition of the outcome.

**Data Complexity:** *HIGH – mortality may not be observable in medical records or recorded in claims data; national mortality data linkages include lags.*

ICD-9 and 10 codes exist for bleeding and/or hemorrhage at different anatomical sites. Many articles have provided potential code lists. In this write up, we are not focusing on examining the validity of various codes. Based on our experience, the greatest challenge relates to capture of deaths in claims or EHR data. In general, EHR and claims data do not capture the occurrence of death in a sensitive or comprehensive way. It is unknown if data that can be used to identify a fatal bleed are accessible. If a patient death occurs during the initial hospitalization for the causative bleed, or at a rehospitalization within the same healthcare system, then the death is likely to be captured. If a patient death occurs in a different hospital or out of hospital (more common for intracranial bleeds), the death is likely to be missed. Autopsies are rarely done. If one was done, it is unlikely the data from the autopsy would be included in the EHR.

From our correspondence with the FDA, we understand that currently there is not access to National Death Index (NDI) data. In the future, claims data and EHR data may be complemented with additional data sources to improve capture of mortality. Lags in NDI data available for linkage may impact the usability of death data for computable phenotyping.

**Data Sources:**

| Necessary EHR Data | Useful EHR Data |
|---|---|
| Diagnostic codes (to establish presence of bleeding) | |

**Conclusion/Fitness for Purpose Recommendation:** *HARD*

While fatal bleeds meet the criteria for low clinical complexity because mortality is an unambiguous endpoint, data complexity is high owing to the current incompleteness of data on mortality in claims or EHR sources. If a decision is made to pursue development of a computable phenotype, the challenge of accessing mortality data will have to be solved.

#### References

Al-Ani, F. et al. (2015) 'Identifying venous thromboembolism and major bleeding in emergency room discharges using administrative data', Thrombosis research. Elsevier BV, 136(6), pp. 1195–1198.

Abdullah S, Young-Min SA, Hudson SJ, Kelly CA, Heycock CR, Hamilton JD. Gross synovial fluid analysis in the differential diagnosis of joint effusion. J Clin Pathol. 2007 Oct;60(10):1144-7. doi: 10.1136/jcp.2006.043950. Epub 2007 Jan 26. PMID: 17259296; PMCID: PMC2014834.

Alonso A, Norby FL, MacLehose RF, Zakai NA, Walker RF, Adam TJ, Lutsey PL. Claims-Based Score for the Prediction of Bleeding in a Contemporary Cohort of Patients Receiving Oral Anticoagulation for Venous Thromboembolism. J Am Heart Assoc. 2021 Sep 21;10(18):e021227. doi: 10.1161/JAHA.121.021227. Epub 2021 Sep 13. PMID: 34514806; PMCID: PMC8649528.

Berger K, Kase CS, Buring JE. Interobserver agreement in the classification of stroke in the physicians' health study. Stroke. 1996 Feb;27(2):238-42. doi: 10.1161/01.str.27.2.238. PMID: 8571416.

Claxton JS, MacLehose RF, Lutsey PL, Norby FL, Chen LY, O'Neal WT, Chamberlain AM, Bengtson LGS, Alonso A. A new model to predict major bleeding in patients with atrial fibrillation using warfarin or direct oral anticoagulants. PLoS One. 2018 Sep 10;13(9):e0203599. doi: 10.1371/journal.pone.0203599. PMID: 30199542; PMCID: PMC6130859.

Coleman CI, Vaitsiakhovich T, Nguyen E, Weeda ER, Sood NA, Bunz TJ, Schaefer B, Meinecke AK, Eriksson D. Agreement between coding schemas used to identify bleeding-related hospitalizations in claims analyses of nonvalvular atrial fibrillation patients. Clin Cardiol. 2018 Jan;41(1):119-125. doi: 10.1002/clc.22861. Epub 2018 Jan 23. PMID: 29360144; PMCID: PMC6489698.

Columbo JA, Daya N, Colantonio LD, Wang Z, Foti K, Hyacinth HI, Johansen MC, Gottesman R, Goodney PP, Howard VJ, Muntner P, Schneider ALC, Selvin E, Hicks CW. Derivation and Validation of ICD-10 Codes for Identifying Incident Stroke. JAMA Neurol. 2024 Aug 1;81(8):875-881. doi: 10.1001/jamaneurol.2024.2044. PMID: 38949838; PMCID: PMC11217886.

de Burgos-Gonzalez A, Bryant V, Maciá-Martinez MA, Huerta C. A strategy for assessment and validation of major bleeding cases in a primary health care database in Spain. Pharmacoepidemiol Drug Saf. 2021; 30(12): 1696-1702. doi:10.1002/pds.5357

Delate T, Jones AE, Clark NP, Witt DM. Assessment of the coding accuracy of warfarin-related bleeding events. Thromb Res. 2017 Nov;159:86-90. doi: 10.1016/j.thromres.2017.10.004. Epub 2017 Oct 12. PMID: 29035718.

Greenberg SM, Ziai WC, Cordonnier C, Dowlatshahi D, Francis B, Goldstein JN, Hemphill JC 3rd, Johnson R, Keigher KM, Mack WJ, Mocco J, Newton EJ, Ruff IM, Sansing LH, Schulman S, Selim MH, Sheth KN, Sprigg N, Sunnerhagen KS; American Heart Association/American Stroke Association. 2022 Guideline for the Management of Patients With Spontaneous Intracerebral Hemorrhage: A Guideline From the

American Heart Association/American Stroke Association. Stroke. 2022 Jul;53(7):e282-e361. doi: 10.1161/STR.0000000000000407. Epub 2022 May 17. PMID: 35579034.

Grossman RI. Head Trauma. In: Neuroradiology: The Requisites, 2nd ed, Mosby, Philadelphia 2003. p.243.

Hald SM, Kring Sloth C, Hey SM, Madsen C, Nguyen N, García Rodríguez LA, Al-Shahi Salman R, Möller S, Poulsen FR, Pottegård A, Gaist D. Intracerebral hemorrhage: positive predictive value of diagnosis codes in two nationwide Danish registries. Clin Epidemiol. 2018;10:941-948 https://doi.org/10.2147/CLEP.S167576

Hanley J, McKernan A, Creagh MD, Classey S, McLaughlin P, Goddard N, Briggs PJ, Frostick S, Giangrande P, Wilde J, Thachil J, Chowdary P; Musculoskeletal Working Party of the UKHCDO. Guidelines for the management of acute joint bleeds and chronic synovitis in haemophilia: A United Kingdom Haemophilia Centre Doctors' Organisation (UKHCDO) guideline. Haemophilia. 2017 Jul;23(4):511-520. doi: 10.1111/hae.13201. Epub 2017 Mar 30. PMID: 28370924.

Hartenstein A, Abdelgawwad K, Kleinjung F, Privitera S, Viethen T, Vaitsiakhovich T. Identification of International Society on Thrombosis and Haemostasis major and clinically relevant non-major bleed events from electronic health records: a novel algorithm to enhance data utilisation from real-world sources. Int J Popul Data Sci. 2023 Oct 2;8(1):2144. doi: 10.23889/ijpds.v8i1.2144. PMID: 38414540; PMCID: PMC10898215.

Hoeven LRV, Bruijne MC, Kemper PF, Koopman MMW, Rondeel JMM, Leyte A, Koffijberg H, Janssen MP, Roes KCB. Validation of multisource electronic health record data: an application to blood transfusion data. BMC Med Inform Decis Mak. 2017 Jul 14;17(1):107. doi: 10.1186/s12911-017-0504-7. PMID: 28709453; PMCID: PMC5512751.

Hsieh MT, Huang KC, Hsieh CY, Tsai TT, Chen LC, Sung SF. Validation of ICD-10-CM Diagnosis Codes for Identification of Patients with Acute Hemorrhagic Stroke in a National Health Insurance Claims Database. Clin Epidemiol. 2021 Jan 14;13:43-51. doi: 10.2147/CLEP.S288518. PMID: 33469381; PMCID: PMC7813455.

Hyun S, Johnson SB, Bakken S. Exploring the ability of natural language processing to extract data from nursing narratives. Comput Inform Nurs. 2009 Jul-Aug;27(4):215-23; quiz 224-5. doi: 10.1097/NCN.0b013e3181a91b58. PMID: 19574746; PMCID: PMC4415266.

Kan P, Fiorella D, Dabus G, Samaniego EA, Lanzino G, Siddiqui AH, Chen H, Khalessi AA, Pereira VM, Fifi JT, Bain MD, Colby GP, Wakhloo AK, Arthur AS; ARISE I Academic Industry Roundtable. ARISE I Consensus Statement on the Management of Chronic Subdural Hematoma. Stroke. 2024 May;55(5):1438-1448. doi: 10.1161/STROKEAHA.123.044129. Epub 2024 Apr 22. PMID: 38648281.

Kleinman S, Busch MP, Murphy EL, Shan H, Ness P, Glynn SA; National Heart, Lung, and Blood Institute Recipient Epidemiology and Donor Evaluation Study (REDS-III). The National Heart, Lung, and Blood Institute Recipient Epidemiology and Donor Evaluation Study (REDS-III): a research program striving to improve blood donor and transfusion recipient outcomes. Transfusion. 2014 Mar;54(3 Pt 2):942-55. doi: 10.1111/trf.12468. Epub 2013 Nov 4. PMID: 24188564; PMCID: PMC4383641.

Li, R. et al. (2019) 'Detection of bleeding events in electronic health record notes using convolutional neural network models enhanced with recurrent neural network autoencoders: Deep learning approach', JMIR medical informatics. JMIR Publications Inc., 7(1), p. e10788.

Lombardi M, Cardenas AC. Hemarthrosis. [Updated 2023 Jul 31]. In: StatPearls [Internet]. Treasure Island (FL): StatPearls Publishing; 2024 Jan-. Available from: https://www.ncbi.nlm.nih.gov/books/NBK525999/

Lotterman S, Sharma S. Blood Transfusion. [Updated 2023 Jun 20]. In: StatPearls [Internet]. Treasure Island (FL): StatPearls Publishing; 2024 Jan-. Available from: https://www.ncbi.nlm.nih.gov/books/NBK499824/.

Lovelock CE, Anslow P, Molyneux AJ, Byrne JV, Kuker W, Pretorius PM, Coull A, Rothwell PM. Substantial observer variability in the differentiation between primary intracerebral hemorrhage and hemorrhagic transformation of infarction on CT brain imaging. Stroke. 2009 Dec;40(12):3763-7. doi: 10.1161/STROKEAHA.109.553933. Epub 2009 Oct 8. PMID: 19815830.

McBride W 2024, [Literature review current through: Oct 2024; topic last updated: Oct 02, 2024] Subdural hematoma in adults: Etiology, clinical features, and diagnosisin UpToDate® Available from: https://www.uptodate.com/contents/subdural-hematoma-in-adults-etiology-clinical-features-and-diagnosis?search=Intracranial%20hemorrhage%20&source=search_result&selectedTitle=4%7E150&usage_type=default&display_rank=4

Mehran R, Rao SV, Bhatt DL, Gibson CM, Caixeta A, Eikelboom J, Kaul S, Wiviott SD, Menon V, Nikolsky E, Serebruany V, Valgimigli M, Vranckx P, Taggart D, Sabik JF, Cutlip DE, Krucoff MW, Ohman EM, Steg PG, White H. Standardized bleeding definitions for cardiovascular clinical trials: a consensus report from the Bleeding Academic Research Consortium. Circulation. 2011 Jun 14;123(23):2736-47. doi: 10.1161/CIRCULATIONAHA.110.009449. PMID: 21670242.

Menis M, Burwen DR, Holness L, Anderson SA. Blood use in the ambulatory setting among elderly in the United States. Transfusion. 2009 Jun;49(6):1186-94. doi: 10.1111/j.1537-2995.2009.02114.x. Epub 2009 Feb 27. PMID: 19309470.

Oger, E., Botrel, MA., Juchault, C. et al. Sensitivity and specificity of an algorithm based on medico-administrative data to identify hospitalized patients with major bleeding presenting to an emergency department. BMC Med Res Methodol 19, 194 (2019). https://doi.org/10.1186/s12874-019-0841-6

Peters CB, Hansen JL, Halwani A, Cho ME, Leng J, Huynh T, Burningham Z, Caloyeras J, Matsuda T, Sauer BC. Validation of Algorithms Used to Identify Red Blood Cell Transfusion Related Admissions in Veteran Patients with End Stage Renal Disease. EGEMS (Wash DC). 2019 Jul 3;7(1):23. doi: 10.5334/egems.257. PMID: 31304183; PMCID: PMC6611485.

Piran S, Schulman S. Treatment of bleeding complications in patients on anticoagulant therapy. Blood. 2019 Jan 31;133(5):425-435. doi: 10.1182/blood-2018-06-820746. Epub 2018 Dec 17. PMID: 30559261.

Pruitt, P. et al. (2019) 'A natural language processing algorithm to extract characteristics of subdural hematoma from head CT reports', Emergency radiology. Springer Science and Business Media LLC, 26(3), pp. 301–306.

Quan, Hude; Parsons, Gerry A.; Ghali, William A. Validity of Procedure Codes in International Classification of Diseases, 9th revision, Clinical Modification Administrative Data. Medical Care 42(8):p 801-809, August 2004. | DOI: 10.1097/01.mlr.0000132391.59713.0d.

Rajashekar D, Liang JW. Intracerebral Hemorrhage. [Updated 2023 Feb 6]. In: StatPearls [Internet]. Treasure Island (FL): StatPearls Publishing; 2024 Jan-. Available from: https://www.ncbi.nlm.nih.gov/books/NBK553103/

Rordorf G and McDonald C, [Literature review current through: Oct 2024; topic last updated: Mar 07, 2024] Spontaneous intracerebral hemorrhage: Pathogenesis, clinical features, and diagnosis in UpToDate® Available from: https://www.uptodate.com/contents/spontaneous-intracerebral-hemorrhage-pathogenesis-clinical-features-and-diagnosis?search=Intracranial+hemorrhage+&source=search_result&selectedTitle=2%7E150&usage_type=default&display_rank=2

Ruigómez, A. et al. (2019) 'Ascertainment and validation of major bleeding events in a primary care database', Pharmacoepidemiology and drug safety. Wiley, 28(2), pp. 148–155.

Schenker C, Marx CE, Kraaijpoel N, Le Gal G, Siegal DM, Klok FA, Aujesky D, Tritschler T. Definitions of fatal bleeding in clinical studies evaluating anticoagulant treatment for venous thromboembolism: A scoping review. J Thromb Haemost. 2023 Jun;21(6):1553-1566. doi: 10.1016/j.jtha.2023.02.013. Epub 2023 Feb 28. PMID: 36858345.

Schulman S, Kearon C; Subcommittee on Control of Anticoagulation of the Scientific and Standardization Committee of the International Society on Thrombosis and Haemostasis. Definition of major bleeding in clinical investigations of antihemostatic medicinal products in non-surgical patients. J Thromb Haemost. 2005 Apr;3(4):692-4. doi: 10.1111/j.1538-7836.2005.01204.x. PMID: 15842354.

Segal JB, Ness PM, Powe NR. Validating billing data for RBC transfusions: a brief report. Transfusion. 2001 Apr;41(4):530-3. doi: 10.1046/j.1537-2995.2001.41040530.x. PMID: 11316905.

Seto, A. et al. (2009) 'Definition of major bleeding used by US anticoagulation clinics', Thrombosis research. Elsevier BV, 124(2), pp. 239–240.

Shehab, Nadine, et al. "Assessment of ICD-10-CM code assignment validity for case finding of outpatient anticoagulant-related bleeding among Medicare beneficiaries." Pharmacoepidemiology and drug safety 28.7 (2019): 951-964.

Spahn, D.R., Bouillon, B., Cerny, V. et al. The European guideline on management of major bleeding and coagulopathy following trauma: fifth edition. Crit Care 23, 98 (2019). https://doi.org/10.1186/s13054-019-2347-3

Szczepiorkowski ZM, Dunbar NM. Transfusion guidelines: when to transfuse. Hematology Am Soc Hematol Educ Program. 2013;2013:638-44.

Taggart, M. et al. (2018) 'Comparison of 2 natural language processing methods for identification of bleeding among critically ill patients', JAMA network open. American Medical Association, 1(6), p. e183451.

Tenny S, Thorell W. Intracranial Hemorrhage. [Updated 2024 Feb 17]. In: StatPearls [Internet]. Treasure Island (FL): StatPearls Publishing; 2024 Jan-. Available from: https://www.ncbi.nlm.nih.gov/books/NBK470242/

Tomaselli GF, Mahaffey KW, Cuker A, Dobesh PP, Doherty JU, Eikelboom JW, Florido R, Gluckman TJ, Hucker WJ, Mehran R, Messé SR, Perino AC, Rodriguez F, Sarode R, Siegal DM, Wiggins BS. 2020 ACC Expert Consensus Decision Pathway on Management of Bleeding in Patients on Oral Anticoagulants: A Report of the American College of Cardiology Solution Set Oversight Committee. J Am Coll Cardiol. 2020 Aug 4;76(5):594-622. doi: 10.1016/j.jacc.2020.04.053. Epub 2020 Jul 14. Erratum in: J Am Coll Cardiol. 2021 Jun 1;77(21):2760. doi: 10.1016/j.jacc.2021.04.020. PMID: 32680646.

van der Ende NAM, Luijten SPR, Kluijtmans L, Postma AA, Cornelissen SA, van Hattem AMG, Lycklama À Nijeholt GJ, Bokkers RPH, Thomassen L, Waje-Andreassen U, Logallo N, Bracard S, Gory B, Roozenbeek B, Dippel DWJ, van der Lugt A. Interobserver Agreement on Intracranial Hemorrhage on Magnetic Resonance Imaging in Patients With Ischemic Stroke. Stroke. 2023 Jun;54(6):1587-1592. doi: 10.1161/STROKEAHA.122.042145. Epub 2023 May 8. PMID: 37154054.

Wells GA, Elliott J, Kelly S, et al. Dual Antiplatelet Therapy Following Percutaneous Coronary Intervention: Clinical and Economic Impact of Standard Versus Extended Duration [Internet]. Ottawa (ON): Canadian Agency for Drugs and Technologies in Health; 2019 Mar. (CADTH Optimal Use Report, No. 9.2b.) Appendix 10, Bleeding Classification System Definitions. Available from: https://www.ncbi.nlm.nih.gov/books/NBK542934/

Xu Y, Gomes T, Wells PS, Pequeno P, Johnson A, Sholzberg M. Evaluation of definitions for oral anticoagulant-associated major bleeding: A population-based cohort study. Thromb Res. 2022 May;213:57-64. doi: 10.1016/j.thromres.2022.02.018. Epub 2022 Feb 24. PMID: 35298939.

Yap, A. J. Y. et al. (2024) 'Validation of a major and clinically relevant nonmajor bleeding phenotyping algorithm on electronic health records', Pharmacoepidemiology and drug safety. Wiley, 33(8), p. e5875.

### Appendix 1: Commonly Used Criteria for Major Bleed[1]

| Criterion *(any one meets definition)* | International Society on Thrombosis and Haemostasis (ISTH)[2] | Bleeding Academic Research Consortium (BARC) (Class IIIA-C, V) | Thrombolysis in Myocardial Infarction (TIMI) |
|---|---|---|---|
| **Mortality** | Fatal bleeding | Fatal bleeding | Fatal bleeding |
| **Hemoglobin drop** | ≥2 g/dL | ≥3 g/dL | ≥5 g/dL |
| **Site of bleed** | Intracranial<br>Tamponade<br>Intraocular<br>Intra-spinal<br>Intra-articular<br>Intra-muscular with compartment syndrome | Intracranial<br>Tamponade<br>Intraocular | Intracranial<br>Intraocular |
| **Transfusion** | ≥2 units packed red blood cells | Any transfusion | |
| **Other** | If **surgical** site bleeding: requires a second intervention, is unexpected and prolonged and/ or sufficiently large | Surgical intervention<br>Vasopressor requirement | |

[1] Adapted from Xu et al., 2022.

[2] Used by the American College of Cardiology.

### Appendix 2: ICD Diagnostic Codes Relevant to Bleeds

This list of ICD-9 and -10 codes was compiled from Hartenstein et al 2023. It is not intended to be exhaustive but is provided for reference purposes.

**Critical Organ bleed disease codes**

**ICD-9-CM**

360.43 Haemophthalmias, except current injury
362.43 Haemorrhagic detachment of retinal pigment epithelium
362.81 Retinal haemorrhage
363.6 Choroidal haemorrhage and rupture
363.61 Choroidal haemorrhage, unspecified
363.62 Expulsive choroidal haemorrhage
363.63 Choroidal rupture
363.72 Haemorrhagic choroidal detachment
364.41 Hyphema of iris and ciliary body
376.32 Orbital haemorrhage
379.23 Vitreous haemorrhage
423.0 Haemopericardium
423.3 Cardiac tamponade
430 Subarachnoid haemorrhage
431 Intracerebral haemorrhage
432 Other and unspecified intracranial haemorrhage
432.0 Non-traumatic extradural haemorrhage
432.1 Subdural haemorrhage
432.9 Unspecified intracranial haemorrhage
441.1 Thoracic aneurysm, ruptured
441.3 Abdominal aneurysm, ruptured
441.5 Aortic aneurysm of unspecified site, ruptured
441.6 Thoraco-abdominal aneurysm, ruptured
719.10 Haemarthrosis, site unspecified
719.11 Haemarthrosis, shoulder region
719.12 Haemarthrosis, upper arm
719.13 Haemarthrosis, forearm
719.14 Haemarthrosis, hand
719.15 Haemarthrosis, pelvic region and thigh
719.16 Haemarthrosis, lower leg
719.17 Haemarthrosis, ankle and foot
719.18 Haemarthrosis, other specified sites
719.19 Haemarthrosis, multiple sites
729.71 Non-traumatic compartment syndrome of upper extremity
729.72 Non-traumatic compartment syndrome of lower extremity
729.73 Non-traumatic compartment syndrome of abdomen
729.79 Non-traumatic compartment syndrome of other sites
998.11 Haemorrhage complicating a procedure

**ICD-10-CM**

H05.23 Haemorrhage of orbit
H05.231 Haemorrhage of right orbit
H05.232 Haemorrhage of left orbit
H05.233 Haemorrhage of bilateral orbit
H05.239 Haemorrhage of unspecified orbit
H21.0 Hyphema
H21.00 Hyphema, unspecified eye
H21.01 Hyphema, right eye
H21.02 Hyphema, left eye
H21.03 Hyphema, bilateral
H31.30 Unspecified choroidal haemorrhage
H31.301 Unspecified choroidal haemorrhage, right eye
H31.302 Unspecified choroidal haemorrhage, left eye
H31.303 Unspecified choroidal haemorrhage, bilateral
H31.309 Unspecified choroidal haemorrhage, unspecified eye
H31.31 Expulsive choroidal haemorrhage
H31.311 Expulsive choroidal haemorrhage, right eye
H31.312 Expulsive choroidal haemorrhage, left eye
H31.313 Expulsive choroidal haemorrhage, bilateral
H31.319 Expulsive choroidal haemorrhage, unspecified eye
H31.41 Haemorrhagic choroidal detachment
H31.411 Haemorrhagic choroidal detachment, right eye
H31.412 Haemorrhagic choroidal detachment, left eye
H31.413 Haemorrhagic choroidal detachment, bilateral
H31.419 Haemorrhagic choroidal detachment, unspecified eye
H35.6 Retinal haemorrhage
H35.60 Retinal haemorrhage, unspecified eye
H35.61 Retinal haemorrhage, right eye
H35.62 Retinal haemorrhage, left eye
H35.63 Retinal haemorrhage, bilateral
H35.73 Haemorrhagic detachment of retinal pigment epithelium
H35.731 Haemorrhagic detachment of retinal pigment epithelium, right eye
H35.732 Haemorrhagic detachment of retinal pigment epithelium, left eye
H35.733 Haemorrhagic detachment of retinal pigment epithelium, bilateral
H35.739 Haemorrhagic detachment of retinal pigment epithelium, unspecified eye
H43.1 Vitreous haemorrhage
H43.10 Vitreous haemorrhage, unspecified eye
H43.11 Vitreous haemorrhage, right eye
H43.12 Vitreous haemorrhage, left eye
H43.13 Vitreous haemorrhage, bilateral
H44.81 Haemophthalmos
H44.811 Haemophthalmos, right eye
H44.812 Haemophthalmos, left eye
H44.813 Haemophthalmos, bilateral

H44.819 Haemophthalmos, unspecified eye
I23.0 Haemopericardium as current complication following acute myocardial infarction
I31.2 Haemopericardium, not elsewhere classified
I31.4 Cardiac tamponade
I60 Non-traumatic subarachnoid haemorrhage
I60.0 Non-traumatic subarachnoid haemorrhage from carotid siphon and bifurcation
I60.00 Non-traumatic subarachnoid haemorrhage from unspecified carotid siphon and bifurcation
I60.01 Non-traumatic subarachnoid haemorrhage from right carotid siphon and bifurcation
I60.02 Non-traumatic subarachnoid haemorrhage from left carotid siphon and bifurcation
I60.1 Non-traumatic subarachnoid haemorrhage from middle cerebral artery
I60.10 Non-traumatic subarachnoid haemorrhage from unspecified middle cerebral artery
I60.11 Non-traumatic subarachnoid haemorrhage from right middle cerebral artery
I60.12 Non-traumatic subarachnoid haemorrhage from left middle cerebral artery
I60.2 Non-traumatic subarachnoid haemorrhage from anterior communicating artery
I60.3 Non-traumatic subarachnoid haemorrhage from posterior communicating artery
I60.30 Non-traumatic subarachnoid haemorrhage from unspecified posterior communicating artery
I60.31 Non-traumatic subarachnoid haemorrhage from right posterior communicating artery
I60.32 Non-traumatic subarachnoid haemorrhage from left posterior communicating artery
I60.4 Non-traumatic subarachnoid haemorrhage from basilar artery
I60.5 Non-traumatic subarachnoid haemorrhage from vertebral artery
I60.50 Non-traumatic subarachnoid haemorrhage from unspecified vertebral artery
I60.51 Non-traumatic subarachnoid haemorrhage from right vertebral artery
I60.52 Non-traumatic subarachnoid haemorrhage from left vertebral artery
I60.6 Non-traumatic subarachnoid haemorrhage from other intracranial arteries
I60.7 Non-traumatic subarachnoid haemorrhage from unspecified intracranial artery
I60.8 Other non-traumatic subarachnoid haemorrhage
I60.9 Non-traumatic subarachnoid haemorrhage, unspecified
I61 Non-traumatic intracerebral haemorrhage
I61.0 Non-traumatic intracerebral haemorrhage in hemisphere, subcortical
I61.1 Non-traumatic intracerebral haemorrhage in hemisphere, cortical
I61.2 Non-traumatic intracerebral haemorrhage in hemisphere, unspecified
I61.3 Non-traumatic intracerebral haemorrhage in brain stem
I61.4 Non-traumatic intracerebral haemorrhage in cerebellum
I61.5 Non-traumatic intracerebral haemorrhage, intraventricular
I61.6 Non-traumatic intracerebral haemorrhage, multiple localised
I61.8 Other non-traumatic intracerebral haemorrhage
I61.9 Non-traumatic intracerebral haemorrhage, unspecified

I62 Other and unspecified non-traumatic intracranial haemorrhage
I62.0 Non-traumatic subdural haemorrhage
I62.00 Non-traumatic subdural haemorrhage, unspecified
I62.01 Non-traumatic acute subdural haemorrhage
I62.02 Non-traumatic subacute subdural haemorrhage
I62.03 Non-traumatic chronic subdural haemorrhage
I62.1 Non-traumatic extradural haemorrhage
I62.9 Non-traumatic intracranial haemorrhage, unspecified
I71.1 Thoracic aortic aneurysm, ruptured
I71.3 Abdominal aortic aneurysm, ruptured
I71.5 Thoraco-abdominal aortic aneurysm, ruptured
I71.8 Aortic aneurysm of unspecified site, ruptured
M25.0 Haemarthrosis
M25.00 Haemarthrosis, unspecified joint
M25.01 Haemarthrosis, shoulder
M25.011 Haemarthrosis, right shoulder
M25.012 Haemarthrosis, left shoulder
M25.019 Haemarthrosis, unspecified shoulder
M25.02 Haemarthrosis, elbow
M25.021 Haemarthrosis, right elbow
M25.022 Haemarthrosis, left elbow
M25.029 Haemarthrosis, unspecified elbow
M25.03 Haemarthrosis, wrist
M25.031 Haemarthrosis, right wrist
M25.032 Haemarthrosis, left wrist
M25.039 Haemarthrosis, unspecified wrist
M25.04 Haemarthrosis, hand
M25.041 Haemarthrosis, right hand
M25.042 Haemarthrosis, left hand
M25.049 Haemarthrosis, unspecified hand
M25.05 Haemarthrosis, hip
M25.051 Haemarthrosis, right hip
M25.052 Haemarthrosis, left hip
M25.059 Haemarthrosis, unspecified hip
M25.06 Haemarthrosis, knee
M25.061 Haemarthrosis, right knee
M25.062 Haemarthrosis, left knee
M25.069 Haemarthrosis, unspecified knee
M25.07 Haemarthrosis, ankle and foot
M25.071 Haemarthrosis, right ankle
M25.072 Haemarthrosis, left ankle
M25.073 Haemarthrosis, unspecified ankle
M25.074 Haemarthrosis, right foot
M25.075 Haemarthrosis, left foot
M25.076 Haemarthrosis, unspecified foot
M25.08 Haemarthrosis, other specified site
M79.A Non-traumatic compartment syndrome
M79.A1 Non-traumatic compartment syndrome of upper extremity

M79.A11 Non-traumatic compartment syndrome of right upper extremity
M79.A12 Non-traumatic compartment syndrome of left upper extremity
M79.A19 Non-traumatic compartment syndrome of unspecified upper extremity
M79.A2 Non-traumatic compartment syndrome of lower extremity
M79.A21 Non-traumatic compartment syndrome of right lower extremity
M79.A22 Non-traumatic compartment syndrome of left lower extremity
M79.A29 Non-traumatic compartment syndrome of unspecified lower extremity
M79.A3 Non-traumatic compartment syndrome of abdomen
M79.A9 Non-traumatic compartment syndrome of other sites

**Overt Bleed Disease Codes**

**ICD-9-CM**

246.3 Haemorrhage and infarction of thyroid
287.8 Other specified haemorrhagic conditions
287.9 Unspecified haemorrhagic conditions
374.81 Haemorrhage of eyelid
377.42 Haemorrhage in optic nerve sheaths
456.0 Oesophageal varices with bleeding
456.20 Oesophageal varices in diseases classified elsewhere, with bleeding
459.0 Haemorrhage, unspecified
530.21 Ulcer of oesophagus with bleeding
530.7 Gastro-oesophageal laceration-haemorrhage syndrome
530.82 Oesophageal haemorrhage
531.0 Acute gastric ulcer with haemorrhage
531.00 Acute gastric ulcer with haemorrhage, without mention of obstruction
531.01 Acute gastric ulcer with haemorrhage, with obstruction
531.2 Acute gastric ulcer with haemorrhage and perforation
531.20 Acute gastric ulcer with haemorrhage and perforation, without mention of obstruction
531.21 Acute gastric ulcer with haemorrhage and perforation, with obstruction
531.4 Chronic or unspecified gastric ulcer with haemorrhage
531.40 Chronic or unspecified gastric ulcer with haemorrhage, without mention of obstruction
531.41 Chronic or unspecified gastric ulcer with haemorrhage, with obstruction
531.6 Chronic or unspecified gastric ulcer with haemorrhage and perforation
531.60 Chronic or unspecified gastric ulcer with haemorrhage and perforation, without mention of obstruction
531.61 Chronic or unspecified gastric ulcer with haemorrhage and perforation, with obstruction
532.0 Acute duodenal ulcer with haemorrhage
532.00 Acute duodenal ulcer with haemorrhage, without mention of obstruction
532.01 Acute duodenal ulcer with haemorrhage, with obstruction
532.2 Acute duodenal ulcer with haemorrhage and perforation
532.20 Acute duodenal ulcer with haemorrhage and perforation, without mention of obstruction
532.21 Acute duodenal ulcer with haemorrhage and perforation, with obstruction

532.4 Chronic or unspecified duodenal ulcer with haemorrhage
532.40 Chronic or unspecified duodenal ulcer with haemorrhage, without mention of obstruction
532.41 Chronic or unspecified duodenal ulcer with haemorrhage, with obstruction
532.6 Chronic or unspecified duodenal ulcer with haemorrhage and perforation
532.60 Chronic or unspecified duodenal ulcer with haemorrhage and perforation, without mention of obstruction
532.61 Chronic or unspecified duodenal ulcer with haemorrhage and perforation, with obstruction
533.0 Acute peptic ulcer of unspecified site with haemorrhage
533.00 Acute peptic ulcer of unspecified site with haemorrhage, without mention of obstruction
533.01 Acute peptic ulcer of unspecified site with haemorrhage, with obstruction
533.2 Acute peptic ulcer of unspecified site with haemorrhage and perforation
533.20 Acute peptic ulcer of unspecified site with haemorrhage and perforation, without mention of obstruction
533.21 Acute peptic ulcer of unspecified site with haemorrhage and perforation, with obstruction
533.4 Chronic or unspecified peptic ulcer of unspecified site with haemorrhage
533.40 Chronic or unspecified peptic ulcer of unspecified site with haemorrhage, without mention of obstruction
533.41 Chronic or unspecified peptic ulcer of unspecified site with haemorrhage, with obstruction
533.6 Chronic or unspecified peptic ulcer of unspecified site with haemorrhage and perforation
533.60 Chronic or unspecified peptic ulcer of unspecified site with haemorrhage and perforation, without mention of obstruction
533.61 Chronic or unspecified peptic ulcer of unspecified site with haemorrhage and perforation, with obstruction
534.0 Acute gastrojejunal ulcer with haemorrhage
534.00 Acute gastrojejunal ulcer with haemorrhage, without mention of obstruction
534.01 Acute gastrojejunal ulcer with haemorrhage, with obstruction
534.2 Acute gastrojejunal ulcer with haemorrhage and perforation
534.20 Acute gastrojejunal ulcer with haemorrhage and perforation, without mention of obstruction
534.21 Acute gastrojejunal ulcer with haemorrhage and perforation, with obstruction
534.4 Chronic or unspecified gastrojejunal ulcer with haemorrhage
534.40 Chronic or unspecified gastrojejunal ulcer with haemorrhage, without mention of obstruction
534.41 Chronic or unspecified gastrojejunal ulcer with haemorrhage, with obstruction
534.6 Chronic or unspecified gastrojejunal ulcer with haemorrhage and perforation
534.60 Chronic or unspecified gastrojejunal ulcer with haemorrhage and perforation, without mention of obstruction
534.61 Chronic or unspecified gastrojejunal ulcer with haemorrhage and perforation, with obstruction
535.01 Acute gastritis, with haemorrhage
535.11 Atrophic gastritis, with haemorrhage

535.21 Gastric mucosal hypertrophy, with haemorrhage
535.31 Alcoholic gastritis, with haemorrhage
535.41 Other specified gastritis, with haemorrhage
535.51 Unspecified gastritis and gastroduodenitis, with haemorrhage
535.61 Duodenitis, with haemorrhage
535.71 Eosinophilic gastritis, with haemorrhage
537.83 Angiodysplasia of stomach and duodenum with haemorrhage
537.84 Dieulafoy lesion (haemorrhagic) of stomach and duodenum
562.02 Diverticulosis of small intestine with haemorrhage
562.03 Diverticulitis of small intestine with haemorrhage
562.12 Diverticulosis of colon with haemorrhage
562.13 Diverticulitis of colon with haemorrhage
568.81 Haemoperitoneum (non-traumatic)
569.3 Haemorrhage of rectum and anus
569.85 Angiodysplasia of intestine with haemorrhage
569.86 Dieulafoy lesion (haemorrhagic) of intestine
578 Gastrointestinal haemorrhage
578.0 Haematemesis
578.1 Blood in stool
578.9 Haemorrhage of gastrointestinal tract, unspecified
593.81 Vascular disorders of kidney
596.7 Haemorrhage into bladder wall
599.71 Gross haematuria
602.1 Congestion or haemorrhage of prostate
620.7 Haematoma of broad ligament
621.4 Haematometra
623.6 Vaginal haematoma
624.5 Haematoma of vulva
626.2 Excessive or frequent menstruation
626.5 Ovulation bleeding
626.6 Metrorrhagia
626.7 Postcoital bleeding
626.8 Other disorders of menstruation and other abnormal bleeding from female genital tract
626.9 Unspecified disorders of menstruation and other abnormal bleeding from female genital tract
627.1 Postmenopausal bleeding
629.0 Haematocele, female, not elsewhere classified
729.92 Non-traumatic haematoma of soft tissue
782.7 Spontaneous ecchymoses
784.7 Epistaxis
784.8 Haemorrhage from throat
786.3 Haemoptysis
786.30 Haemoptysis, unspecified
786.31 Acute idiopathic pulmonary haemorrhage in infants [AIPHI]
786.39 Other haemoptysis
790.01 Precipitous drop in haematocrit

Sentinel

**ICD-10-CM**

D68.32 Haemorrhagic disorder due to extrinsic circulating anticoagulants
H47.02 Haemorrhage in optic nerve sheath
H47.021 Haemorrhage in optic nerve sheath, right eye
H47.022 Haemorrhage in optic nerve sheath, left eye
H47.023 Haemorrhage in optic nerve sheath, bilateral
H47.029 Haemorrhage in optic nerve sheath, unspecified eye
H60.32 Haemorrhagic otitis externa
H60.321 Haemorrhagic otitis externa, right ear
H60.322 Haemorrhagic otitis externa, left ear
H60.323 Haemorrhagic otitis externa, bilateral
H60.329 Haemorrhagic otitis externa, unspecified ear
H92.2 Otorrhagia
H92.20 Otorrhagia, unspecified ear
H92.21 Otorrhagia, right ear
H92.22 Otorrhagia, left ear
H92.23 Otorrhagia, bilateral
I85.01 Oesophageal varices with bleeding
I85.11 Secondary oesophageal varices with bleeding
J94.2 Haemothorax
K20.81 Other oesophagitis with bleeding
K20.91 Oesophagitis, unspecified with bleeding
K22.11 Ulcer of oesophagus with bleeding
K22.6 Gastro-oesophageal laceration-haemorrhage syndrome
K25.0 Acute gastric ulcer with haemorrhage
K25.2 Acute gastric ulcer with both haemorrhage and perforation
K25.4 Chronic or unspecified gastric ulcer with haemorrhage
K25.6 Chronic or unspecified gastric ulcer with both haemorrhage and perforation
K26.0 Acute duodenal ulcer with haemorrhage
K26.2 Acute duodenal ulcer with both haemorrhage and perforation
K26.4 Chronic or unspecified duodenal ulcer with haemorrhage
K26.6 Chronic or unspecified duodenal ulcer with both haemorrhage and perforation
K27.0 Acute peptic ulcer, site unspecified, with haemorrhage
K27.2 Acute peptic ulcer, site unspecified, with both haemorrhage and perforation
K27.4 Chronic or unspecified peptic ulcer, site unspecified, with haemorrhage
K27.6 Chronic or unspecified peptic ulcer, site unspecified, with both haemorrhage and perforation
K28.0 Acute gastrojejunal ulcer with haemorrhage
K28.2 Acute gastrojejunal ulcer with both haemorrhage and perforation
K28.4 Chronic or unspecified gastrojejunal ulcer with haemorrhage
K28.6 Chronic or unspecified gastrojejunal ulcer with both haemorrhage and perforation
K29.01 Acute gastritis with bleeding
K29.21 Alcoholic gastritis with bleeding
K29.31 Chronic superficial gastritis with bleeding
K29.41 Chronic atrophic gastritis with bleeding
K29.51 Unspecified chronic gastritis with bleeding

K29.61 Other gastritis with bleeding
K29.71 Gastritis, unspecified, with bleeding
K29.81 Duodenitis with bleeding
K29.91 Gastroduodenitis, unspecified, with bleeding
K31.811 Angiodysplasia of stomach and duodenum with bleeding
K50.011 Crohn's disease of small intestine with rectal bleeding
K50.111 Crohn's disease of large intestine with rectal bleeding
K50.811 Crohn's disease of both small and large intestine with rectal bleeding
K50.911 Crohn's disease, unspecified, with rectal bleeding
K51.011 Ulcerative (chronic) pancolitis with rectal bleeding
K51.211 Ulcerative (chronic) proctitis with rectal bleeding
K51.311 Ulcerative (chronic) rectosigmoiditis with rectal bleeding
K51.411 Inflammatory polyps of colon with rectal bleeding
K51.511 Left-sided colitis with rectal bleeding
K51.811 Other ulcerative colitis with rectal bleeding
K51.911 Ulcerative colitis, unspecified with rectal bleeding
K55.21 Angiodysplasia of colon with haemorrhage
K57.01 Diverticulitis of small intestine with perforation and abscess with bleeding
K57.11 Diverticulosis of small intestine without perforation or abscess with bleeding
K57.13 Diverticulitis of small intestine without perforation or abscess with bleeding
K57.21 Diverticulitis of large intestine with perforation and abscess with bleeding
K57.31 Diverticulosis of large intestine without perforation or abscess with bleeding
K57.33 Diverticulitis of large intestine without perforation or abscess with bleeding
K57.41 Diverticulitis of both small and large intestine with perforation and abscess with bleeding
K57.51 Diverticulosis of both small and large intestine without perforation or abscess with bleeding
K57.53 Diverticulitis of both small and large intestine without perforation or abscess with bleeding
K57.81 Diverticulitis of intestine, part unspecified, with perforation and abscess with bleeding
K57.91 Diverticulosis of intestine, part unspecified, without perforation or abscess with bleeding
K57.93 Diverticulitis of intestine, part unspecified, without perforation or abscess with bleeding
K62.5 Haemorrhage of anus and rectum
K63.81 Dieulafoy lesion of intestine
K66.1 Haemoperitoneum
K92.0 Haematemesis
K92.1 Melena
K92.2 Gastrointestinal haemorrhage, unspecified
K94.01 Colostomy haemorrhage
K94.11 Enterostomy haemorrhage
K94.21 Gastrostomy haemorrhage
K94.31 Oesophagostomy haemorrhage
M79.81 Non-traumatic haematoma of soft tissue
N30.01 Acute cystitis with haematuria
N30.11 Interstitial cystitis (chronic) with haematuria

N30.21 Other chronic cystitis with haematuria
N30.31 Trigonitis with haematuria
N30.41 Irradiation cystitis with haematuria
N30.81 Other cystitis with haematuria
N30.91 Cystitis, unspecified with haematuria
N42.1 Congestion and haemorrhage of prostate
N83.7 Haematoma of broad ligament
N85.7 Haematometra
N92.0 Excessive and frequent menstruation with regular cycle
N92.1 Excessive and frequent menstruation with irregular cycle
N92.3 Ovulation bleeding
N92.4 Excessive bleeding in the pre-menopausal period
N93 Other abnormal uterine and vaginal bleeding
N93.0 Postcoital and contact bleeding
N93.1 Pre-pubertal vaginal bleeding
N93.8 Other specified abnormal uterine and vaginal bleeding
N93.9 Abnormal uterine and vaginal bleeding, unspecified
N95.0 Postmenopausal bleeding
R04.0 Epistaxis
R04.1 Haemorrhage from throat
R04.2 Haemoptysis
R04.81 Acute idiopathic pulmonary haemorrhage in infants
R04.89 Haemorrhage from other sites in respiratory passages
R04.9 Haemorrhage from respiratory passages, unspecified
R23.3 Spontaneous ecchymoses
R31.0 Gross haematuria
R58 Haemorrhage, not elsewhere classified
R71.0 Precipitous drop in haematocrit

### Fitness-for-Purpose (FFP) Assessment:
### Hepatitis B Reactivation

| | |
|---|---|
| **Health Outcome of Interest:** | Hepatitis B Virus Reactivation |
| **Date:** | July 26, 2024 |
| **Subject:** | Computable Phenotype Feasibility Assessment for Hepatitis B Virus Reactivation |
| **Drug(s)/ ARIA Insufficiency Memo(s):** | 1. Anthracyclines class, Memo #2016-2374<br>2. Direct Acting Antiviral Exposure, Memo #2016-2257 |

#### Executive Summary

**Summary:** *MODERATE* – Computable algorithm would require longitudinal laboratory results for four different laboratory tests, whose interpretation can be complex. The availability of these lab results in different settings would need to be explored.

**HOI Description:** Hepatitis B Virus reactivation (HBV-R) is a clinical syndrome characterized either by 1) the reappearance of serum HBV DNA or development of Hepatitis B surface antigen (HBsAg) in individuals with a previous resolved HBV infection, or 2) a substantial increase in serum HBV DNA in individuals with inactive chronic hepatitis B (CHB). HBV-R can occur spontaneously but is often associated with a compromised immune system (e.g., immunosuppressive therapies, autoimmune disease, or organ transplantation).

**ARIA Insufficiency Issue:** There is concern that HBV-R can occur following exposure to anthracyclines or direct-acting antivirals. It can be difficult to distinguish between a new HBV infection and HBV reactivation, and the low HBV prevalence in the general population (<1%) would result in very few reactivations.

**Existing Computable Algorithms:** There are no ICD-9/10 codes for HBV-R, and ICD-9/10 codes cannot distinguish inactive HBV or resolved HBV. Chen et al. (2024) used laboratory results to compute prevalence of HBV-R after initiation of ibrutinib between 2014 and 2019 in national data from the Veterans Administration. In this study, 2219 of 4130 patients were missing key baseline laboratory results prior to initiation. Thirteen patients were identified with HBV-R.

**Clinical Complexity:** *MEDIUM* – HBV-R requires longitudinal results from multiple laboratory tests to identify an increase in HBV DNA compared to baseline or reappearance of Hepatitis B surface antigen (HBsAg+) after prior resolution of infection. Interpretation of these results can be complex.

**Data Complexity:** *MEDIUM* – There are no ICD codes specific to HBV-R. Additionally, HBV laboratory results may be difficult to extract and standardize. Data from multiple laboratory tests are required to establish HBV status at baseline and a second set of laboratory results is needed to demonstrate reactivation. Missing data may be problematic.

**Conclusion / Fitness-for-Purpose Recommendation:** ICD 9/10 codes alone are insufficient, so access to laboratory results would be necessary. This could be challenging due to incomplete or difficult to interpret laboratory results, so it is recommended to explore the availability and interpretability of these laboratory results.

**Limitations:** This fitness-for-purpose recommendation is based on literature reviews as of June 2024. We assume an ideal performance of an algorithm equates to a PPV of at least 80% with adequate sensitivity.

#### Detailed Discussion

##### Description of HOI

Hepatitis B Virus reactivation (HBV-R) is a clinical syndrome characterized either by 1) the reappearance of serum HBV DNA or development of Hepatitis B surface antigen positivity (HBsAg+) in individuals with a previous resolved HBV infection, or 2) a substantial increase in serum HBV DNA in individuals with inactive chronic hepatitis B (CHB). HBV-R can occur spontaneously but is often associated with a compromised immune system such as when using immunosuppressive therapies for cancer, autoimmune disease, or organ transplantation. The definition of HBV-R varies between society guidelines, and there is no standardized criterion for the level of increase in HBV DNA that qualifies as reactivation, but at least a 2-log or 100-fold increase is often used in the medical literature (see Appendix 1).

**Drug from Drug-Outcome Pair:**

1) Anthracyclines class (Doxorubicin, Danorubicin, Epirubicin), Memo #2016-2374
2) Direct-Acting Antivirals for Treatment of Chronic Hepatitis C, Memo #2016-2257

**Reason for ARIA insufficiency concern(s) related to outcome, according to FDA memo(s):** There is concern that HBV-R can occur following exposure to anthracyclines or direct-acting antivirals. Not only is ARIA unable to distinguish between a new HBV infection and an HBV reactivation, but there may be an insufficient population in which to complete the study due to low prevalence of HBV infection in the general population (<1%).[1]

**Previous Validation Studies:**

ICD-9 or -10 codes for HBV-R, specifically, do not exist. Furthermore, ICD-9 and -10 codes do not distinguish inactive or resolved HBV.

- Chen *et al.* (2024) used laboratory results to compute prevalence of HBV-R after initiation of ibrutinib between 2014 and 2019 using national data from the Veterans Administration (VA). Beginning with 4130 patients, 1670 were excluded due to having no antibodies to Hepatitis B core antigen (anti-HBc-), indicating no prior exposure to HBV. Next, 2219 were excluded due to missing anti-HBc results. An additional 60 patients were excluded due to being on antivirals against HBV, leaving 181 patients. Of these, 16 patients were identified as having one of the following after ibrutinib initiation: 1) a change in HBsAg/HBV DNA to positive status, 2) at least a 3-fold increase in alanine aminotransferase levels, or 3) a new prescription of an antiviral. When these 16

[1] The CDC provides a 2021 estimate of 13,300 acute HBV infections and 14,229 chronic HBV infections in the US. (Source: https://www.cdc.gov/hepatitis/statistics/2021surveillance/hepatitis-b.htm, Accessed 6 June 2024)

Sentinel

patients' charts were manually reviewed, 13 had physician-documented HBV-R (verified against a 2018 US guideline), of which 7 were directly attributed to ibrutinib according to physician documentation (6 were attributed to another drug). This approach evaluating labs and medications compared to the gold standard of clinician-documented HBV-R due to ibrutinib would give an estimated PPV of 54% (7/13) and a sensitivity of 44% (7/16).

- A number of studies have tried to validate ICD-9/10 codes for HBV (*not* HBV-R specifically). While a couple of studies over a decade old found promising results using ICD-9 codes (e.g., Mahajan et al. 2013 reports PPV and sensitivity of 90% and 58%, respectively, for 2 diagnosis codes 6 months apart), performance of ICD-10 codes in the U.S. is unknown, and it is not clear how a computable phenotype would be developed to determine HBV-R. Validation of these algorithms was done by using laboratory results.

**Clinical Complexity:** *MEDIUM – Requires more than one set of laboratory results for a patient over time, and there is not agreement among clinical guidelines.*

Complexity is based on the need for clinicians to interpret multiple different HBV lab results to establish HBV status and to compare results at multiple points in time. History of HBV infection would need to be established at baseline using a combination of several test results and in particular, to identify individuals with resolved HBV or chronic inactive HBV. Baseline HBV DNA levels are needed to establish baseline viral load. Lab results at a second point in time would be needed to establish reactivation, with somewhat different criterion applied to people with prior resolved HBV vs. those with chronic inactive HBV. Different professional societies have issued guidelines for identifying HBV reactivation but with differing criteria (Appendix 1), which adds to complexity and difficulty with standardization.

**Data Complexity:** *MEDIUM – Requires longitudinal data that may be missing or incomplete; data may not all be structured and easy to extract.*

Complexity is based on the need for repeated measures of multiple different HBV lab results. Some of these may be numeric results that are easy to extract, but others may exist as structured free text or in PDF form. Laboratory test results are required to establish HBV status prior to starting a therapy, and a second set of lab results is required to demonstrate reactivation.

Determining HBV-R would involve an approach that includes **three phases**:

1. Identification of a history of HBV infection;
2. Characterization as either
   a. chronic inactive HBV, or
   b. resolved HBV (evidence of viral suppression);
3. Evidence of an HBV reactivation: increase in HBV DNA levels (for either HBV status) or HBsAg+ lab results (only for resolved HBV).

For HBV-R to be associated with medication use, the reactivation in (3) would need to be temporally related to initiation of a new medication.

**Phase 1**: Presence of any history of HBV could be identified in various ways, such as ICD diagnosis code for HBV infection, dispensings of medications used to treat HBV (e.g., including entecavir, tenofovir, lamivudine, adefovir, and telbivudine) or codes for lab services (e.g., presence of repeated HBV laboratory testing, indicating monitoring), as well as from laboratory results.

**Phase 2**: Identification would require lab results for HBsAg, anti-HBc, and anti-HBs, and/or HBV DNA (see Appendices).

**Phase 3**: Identified by an elevated level of HBV DNA or, by some guidelines, the seroconversion to HBsAg+.

Missing data are likely and add to complexity. Baseline results may not be available for many individuals. Current US clinical guidelines recommend testing all patients prior to initiation of immunosuppressive therapies (e.g., Reddy *et al*. 2014, Huang *et al*. 2020). Current CDC guidelines issued in 2023 recommend that all adults should undergo HBV screening at least once (Conners et al. 2023). While these promise increased availability of HBV screening results, Chen *et al*. (2024) found that in the VA, over half of those beginning ibrutinib treatment between 2014 and 2019 lacked baseline anti-HBc lab results.[2] In addition, Shah *et al*. (2018) note that even though guidelines recommend screening prior to immunosuppressive therapies, actual adherence to screening guidelines ranged from 13% to 61% from 2003-2011 in the VA. It is likely that a number of individuals at risk for HBV-R would not be identified. The availability of laboratory results during follow-up is also not well characterized. Individuals may receive laboratory testing in different healthcare systems than the one providing their treatment, leading to missing data.

**Data Sources:**

| Necessary EHR Data | Useful EHR Data |
|---|---|
| Lab results (HBsAg, anti-HBc, anti-HBs, HBV DNA) | Medications |
| | ICD-9/10 codes |

**Conclusion / Fitness-for-Purpose (FFP) Recommendation:** *MODERATE DIFFICULTY*

A computable algorithm would require longitudinal laboratory results for four laboratory tests which need to be interpreted in combination. This is likely to be feasible and is easier than some approaches such as those requiring Natural Language Processing of free text. However, the availability of these lab results, and ease of extraction, would need to be explored.

There are no ICD-9 or -10 codes for HBV-R. If ICD-10 codes were to be used to help determine baseline HBV status as part of a computable phenotype, these codes

[2] Current guidance at the VA is to screen for HBV in all patients prior to starting immunosuppressive therapy (see: https://www.hepatitis.va.gov/hbv/reactivation-prevention.asp).

would require validation. Their sensitivity may also be low. It is therefore recommended to use laboratory results.  A challenge is the potential for missing laboratory results, given several studies showing relatively low adherence to guidelines recommending screening prior to initiation of certain therapies.

If a decision is made to pursue development of a computable phenotype, an important recommended step is to explore the availability, ease of extraction, and interpretability of key laboratory results.

#### References

Chen, T.-Y. et al. (2024) ‘Risk of hepatitis B reactivation in patients receiving ibrutinib: The national veterans affairs cohort’, Open forum infectious diseases. Oxford University Press, 11(3), p. ofae008.

Conners, E. E. et al. (2023) ‘Screening and testing for hepatitis B Virus Infection: CDC recommendations - United States, 2023’, Recommendations and reports: Morbidity and mortality weekly report. MMWR Recomm Rep, 72(1), pp. 1–25.

Hwang, J. P. et al. (2020) ‘Hepatitis B virus screening and management for patients with cancer prior to therapy: ASCO Provisional Clinical Opinion update’, Journal of clinical oncology: official journal of the American Society of Clinical Oncology. American Society of Clinical Oncology (ASCO), 38(31), pp. 3698–3715.

Lam, L. et al. (2023) ‘Performance of algorithms for identifying patients with chronic hepatitis B or C infection in the french health insurance claims databases using the ANRS CO22 HEPATHER cohort’, Journal of viral hepatitis, 30(3), pp. 232–241.

Lee, P.-C. et al. (2020) ‘Risk of HBV reactivation in patients with immune checkpoint inhibitor-treated unresectable hepatocellular carcinoma’, Journal for immunotherapy of cancer. BMJ, 8(2), p. e001072.

Mahajan, R. et al. (2013) ‘Use of the International Classification of Diseases, 9th revision, coding in identifying chronic hepatitis B virus infection in health system data: implications for national surveillance’, Journal of the American Medical Informatics Association: JAMIA. Oxford University Press (OUP), 20(3), pp. 441–445.

Reddy, K. R. et al. (2015) ‘American Gastroenterological Association Institute guideline on the prevention and treatment of hepatitis B virus reactivation during immunosuppressive drug therapy’, Gastroenterology. Elsevier BV, 148(1), pp. 215–9; quiz e16-7.

Shah, R. et al. (2018) ‘Hepatitis B virus screening and reactivation in a national VA cohort of patients with inflammatory bowel disease treated with tumor necrosis factor antagonists’, Digestive diseases and sciences. Springer Nature, 63(6), pp. 1551–1557.

Sheu, M.-J. et al. (2020) ‘Validity of ICD-10-CM codes used to identify patients with chronic hepatitis B and C virus infection in administrative claims data from the Taiwan National Health Insurance Outpatient Claims Dataset’, Clinical epidemiology. Informa UK Limited, 12, pp. 185–192.

Smalls, D. J. et al. (2019) ‘Hepatitis B virus reactivation: Risk factors and current management strategies’, Pharmacotherapy. Wiley, 39(12), pp. 1190–1203.

### Appendix 1: Clinical Guidelines for Hepatitis B Virus Reactivation

| | Definition for Reactivation of Chronic HBV | Definition for Reactivation of Resolved HBV |
|---|---|---|
| **Baseline serologies** | **HBsAg⁺, anti-HBc⁺** | **HBsAg⁻, anti-HBc⁺** |
| AASLD 2018 guidelines | If HBV DNA at baseline is unknown: ≥10,000 IU/mL<br>If HBV DNA at baseline is known and previously undetectable: ≥1,000 IU/mL<br>If HBV DNA at baseline is known and previously detectable: ≥100-fold increase | Development of detectable DNA<br>*OR*<br>Development of HBsAg (also known as reverse seroconversion) |
| AGA 2015 guidelines | If HBV DNA at baseline is unknown: undefined<br>If HBV DNA at baseline is known and previously undetectable: *de novo* detectable DNA<br>If HBV DNA at baseline is known and previously detectable: ≥10-fold increase | Development of detectable DNA<br>*OR*<br>Development of HBsAg (also known as reverse seroconversion)<br>*OR*<br>Development of HBeAg |
| APASL 2016 guidelines | If HBV DNA at baseline is unknown: ≥20,000 IU/mL<br>If HBV DNA at baseline is known and previously undetectable: ≥100 IU/mL<br>If HBV DNA at baseline is known and previously detectable: ≥100-fold increase | Development of detectable DNA<br>*OR*<br>Development of HBsAg (also known as reverse seroconversion) |
| EASL 2017 guidelines | Undefined | Undefined |

AASLD, American Association for the Study of Liver Diseases
AGA, American Gastroenterological Association
APASL, Asian Pacific Association for the Study of the Liver
EASL, European Association for the Study of the Liver
HBeAg, Hepatitis B e-antigen
HBsAg, Hepatitis B surface antigen
anti-HBc, antibody to Hepatitis B core antigen

#### Appendix 2: ICD-9 and ICD-10 Diagnostic Codes Relevant to Hepatitis B Virus Reactivation

ICD-9 or -10 codes for HBV-R, inactive HBV, and resolved HBV do not exist.

ICD-9 or -10 codes for Hepatitis B infection may be useful depending on the approach to the computable phenotype. This list of ICD-9 and -10 codes was compiled from various relevant literature. It is not intended to be exhaustive but is provided for reference purposes.

ICD-9

| Code | Description |
| --- | --- |
| 70.2 | Viral hepatitis B with hepatic coma, acute or unspecified, without mention of hepatitis delta |
| 70.21 | Viral hepatitis B with hepatic coma, acute or unspecified, with hepatitis delta |
| 70.22 | Viral hepatitis B with hepatic coma, chronic, without mention of hepatitis delta |
| 70.23 | Viral hepatitis B with hepatic coma, chronic with hepatitis delta |
| 70.31 | Viral hepatitis B without mention of hepatic coma, acute or unspecified, with hepatitis delta |
| 70.32 | Viral hepatitis B without mention of hepatic coma, chronic, without mention of hepatitis delta |
| 70.33 | Viral hepatitis B without mention of hepatic coma, chronic with hepatitis delta |
| 70.42 | Hepatitis delta without mention of active hepatitis B disease with hepatic coma |
| 70.52 | Hepatitis delta without mention of active hepatitis B disease or hepatic coma |
| V02.61 | Hepatitis B carrier |

ICD-10

| Code | Description |
| --- | --- |
| B16.0 | Acute hepatitis B with delta-agent without hepatic coma |
| B16.1 | Acute hepatitis B with delta-agent with hepatic coma |
| B16.2 | Acute hepatitis B without delta-agent with hepatic coma |
| B16.9 | Acute hepatitis B without delta-agent and without hepatic coma |
| B17.0 | Acute delta-(super) infection of hepatitis B carrier |
| B18.0 | Chronic viral hepatitis B with delta-agent |
| B18.1 | Chronic viral hepatitis B without delta-agent |
| B19.1 | Unspecified viral hepatitis B with hepatic coma |

| | |
|---|---|
| B19.2 | Unspecified viral hepatitis B without hepatic coma |

### Appendix 3

##### Table 1. Glossary of Terms

| Term | Abbreviation | Comment |
|---|---|---|
| Hepatitis B Virus | HBV | |
| HBV surface antigen | HBsAg | Viral protein found on the surface of HBV. Detection of HBsAg indicates that the patient is infected (either acute or chronic infection). |
| HBV e antigen | HBeAg | Viral protein whose presence indicates active viral replication. |
| HBV core antibody, Total | anti-HBc | Antibody to HBV. Testing detects both IgM (short-term immunity) and IgG (long-term immunity) antibodies. Positive results indicate prior or ongoing HBV infection. This test is the only way to differentiate vaccination from infection. Appears ~5 weeks after HBV exposure. |
| HBV surface antibody | anti-HBs | Antibody to HBV. Positive results indicate recovery from HBV infection OR successful HBV vaccination. |
| HBV e antibody | anti-HBe | Antibody to HBeAg. |
| Hepatitis B Virus DNA | HBV DNA | A reliable marker of active HBV replication. Viral load test uses PCR to detect and quantify HBV DNA fragments. HBV DNA levels are detectable by 30 days following infection, generally reach a peak at the time of acute hepatitis, and gradually decrease and disappear when the infection resolves spontaneously. |
| Immunoglobulin M antibody | anti-HBc IgM | HBV antibody, the detection of which indicates a new acute infection. |

##### Table 2. Interpretation of screening for status of HBV infection which consists of the 'triple test'. Adapted from Conners (2023)

| HBsAg | anti-HBc | anti-HBs | Interpretation(s) |
|---|---|---|---|
| negative | negative | negative | No current or prior HBV infection |
| negative | negative | positive | Prior HBV vaccination |
| **negative** | **positive** | **positive** | Prior HBV infection* |
| positive | positive | negative | Current HBV infection (likely chronic)* |
| negative | positive | negative | 1) acute HBV (window period)*<br>2) chronic HBV (HBsAg not detectable)*<br>3) recovery from acute HBV (anti-HBs not detectable)*<br>4) False positive anti-HBc |

**At risk for reactivation*

Likely workup for HBV reactivation:

1. HBV screening results of HBsAg-, anti-HBc+, anti-HBs+ prior to medication initiation.

2. Follow-up surveillance with HBsAg+ and/or HBV DNA.

**Figure 1. Typical serologic course of acute hepatitis B to recovery. X-axis is 'weeks after exposure'. (Source: https://www.cdc.gov/mmwr/preview/mmwrhtml/rr5708a1.htm)**

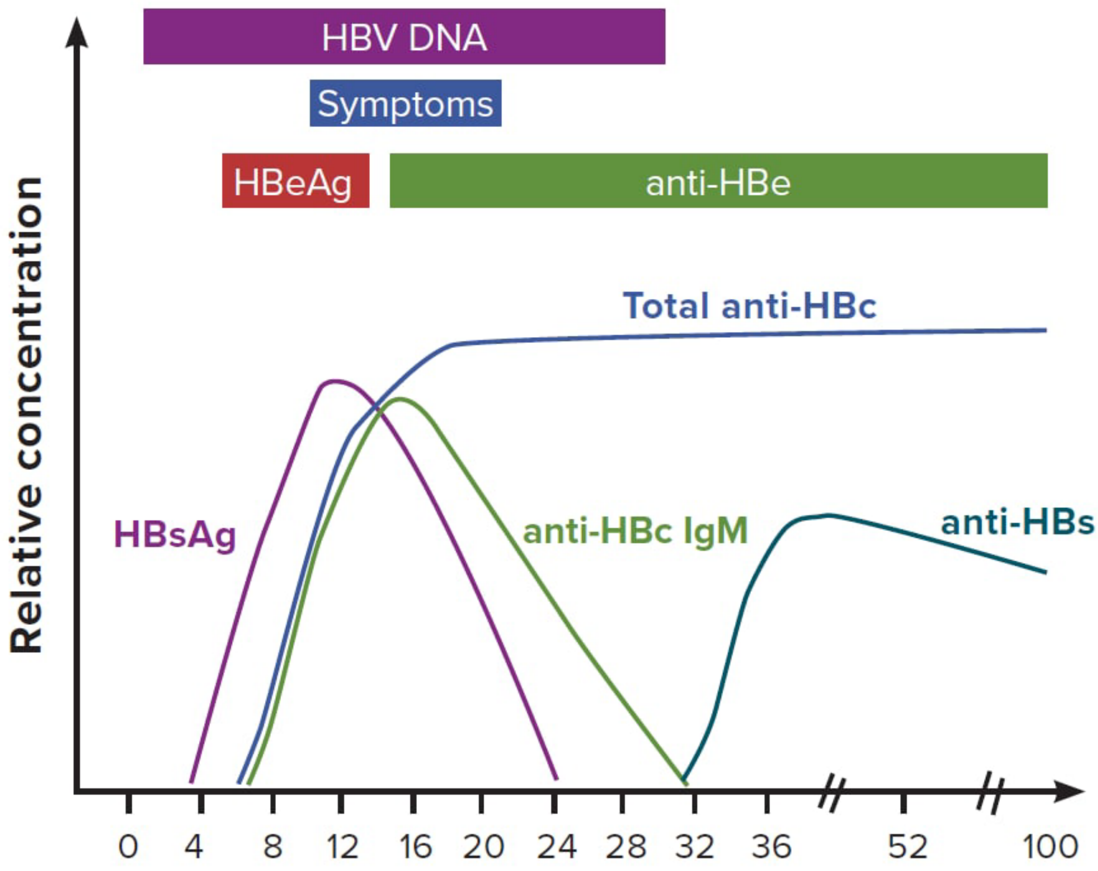

**Figure 2. Typical serologic course of the progression to chronic hepatitis B (Source: https://www.cdc.gov/mmwr/preview/mmwrhtml/rr5708a1.htm)**

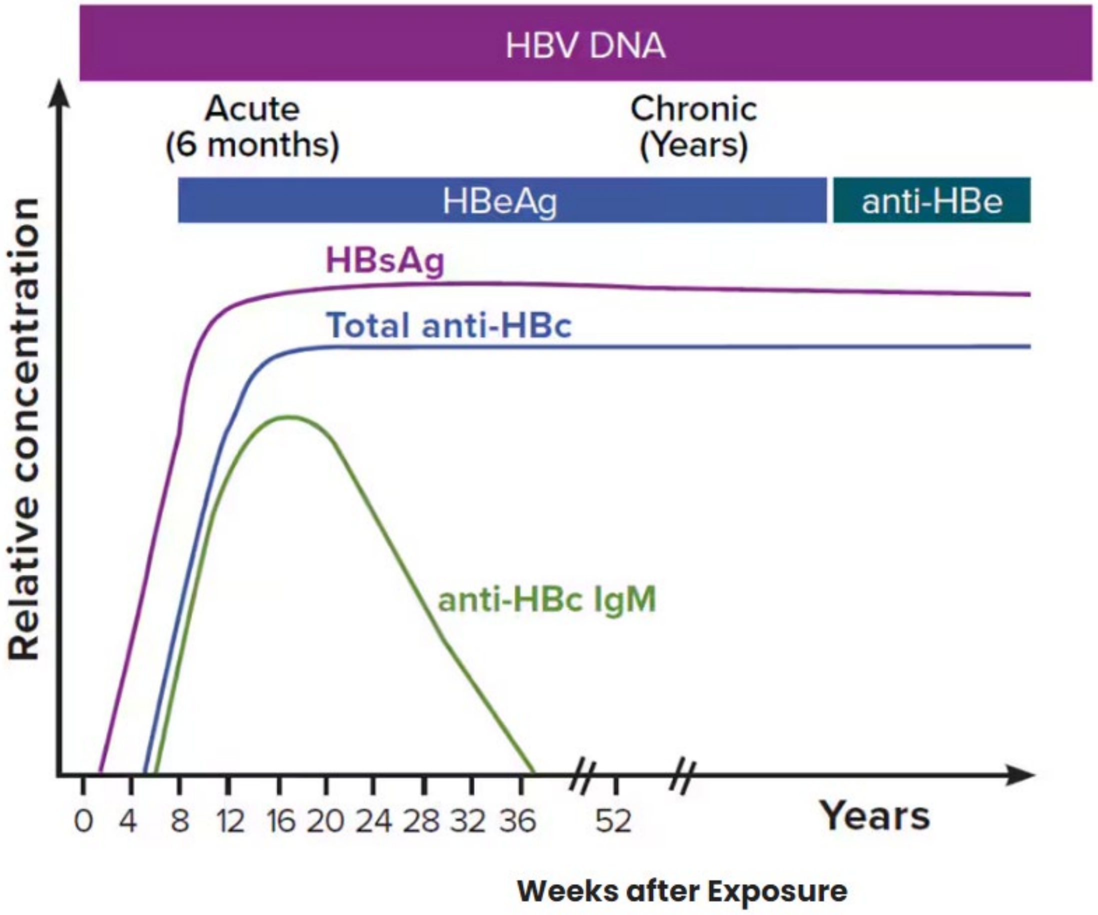

### Fitness-for-Purpose (FFP) Assessment: Hemolytic Anemia

| | |
|---|---|
| **Health Outcome of Interest:** | Hemolytic Anemia |
| **Date:** | January 16, 2025 |
| **Subject:** | Health Outcome of Interest Feasibility Assessment for Hemolytic Anemia |
| **Drug(s)/ ARIA Insufficiency Memo(s):** | 1. Arakoda (tafenoquine), Memo #2018-1054<br>2. Krintafel (tafenoquine), Memo #2018-1054 |

#### Executive Summary

**Overall Assessment:** *MODERATE* – While there is evidence that a claims-based algorithm may be sufficient to identify hemolytic anemia, validation in US data is needed. If the algorithm fails to meet performance characteristics, development of a computable phenotype synthesizing electronic health record (EHR) data from at least five different laboratory results would be required, resulting in MODERATE difficulty.

**ARIA Insufficiency Issue(s):** Diagnosis codes for hemolytic anemia have not been sufficiently validated in administrative data, and laboratory results required for making a diagnosis may not be available in the Sentinel Common Data Model.

**Orientation to HOI:** Hemolytic anemia is an anemia due to the destruction (i.e., hemolysis) of red blood cells (RBCs) in which the production of new RBCs is unable to keep up with the pace of their destruction.

**Existing Computable Algorithms:** ICD-10 validation studies in Denmark (Hansen 2016) and France (Maquet 2019) showed high positive predictive value (PPV) of ICD-10 codes for both acquired and inherited hemolytic anemia when validated against laboratory data: 87% overall (83% for acquired) and 95% overall (93% for acquired), respectively. No studies have validated these algorithms in US data.

**Clinical Complexity:** *MEDIUM* – Hemolytic anemia requires diagnosis of anemia and evidence of hemolysis in the absence of other explanatory factors. This requires laboratory results to verify anemia, identify an unexplained high reticulocyte count, and find evidence of RBC destruction from an increase in unconjugated bilirubin, an increase in lactate dehydrogenase (LDH), or low haptoglobin levels.

**Data Complexity:** *MEDIUM* – A computable phenotype for hemolytic anemia must identify anemia, an unexplained high reticulocyte count, and evidence of RBC destruction. Additionally, the computable phenotype would need to ensure that there were no other explanatory factors or at least rule out some possible contributing conditions. This would require extraction, harmonization and manipulation of multiple laboratory test results.

**Conclusion / Fitness-for-Purpose Recommendation:** A validation study of hemolytic anemia algorithms using ICD-10 codes should be conducted in a US population to evaluate performance. If the validation study fails to meet desired performance characteristics, a computable phenotype could be developed, which would require synthesizing EHR data from at least five different laboratory results.

**Limitations:** This fitness-for-purpose recommendation is based on literature reviews as of January 2025. We assume an ideal performance equates to a PPV of at least 80%.

#### Detailed Discussion

**Reason for ARIA insufficiency concern(s) related to outcome, according to FDA memo(s):** The ARIA insufficiency concerns are related to the broader outcome of hematologic adverse events. The memo notes that "[w]hile Sentinel Common Data Model captures the laboratory results on hemoglobin level, it does not capture other tests relevant to tafenoquine's hematologic AEs" (pg. 6). Specifically related to hemolytic anemia, the memo observes that there are diagnostic codes but that "they are not well-validated in administrative claims databases" (pg. 6), citing only the Danish study discussed below (Hansen 2016).

**Drug from Drug-Outcome Pair:**

1) Arakoda (tafenoquine), Memo #2018-1054
2) Krintafel (tafenoquine), Memo #2018-1054

**Orientation to HOI**

Hemolytic anemia (HA) is an anemia (i.e., reduced number of circulating red blood cells [RBCs]) due specifically to the shortened lifespan of RBCs. The typical RBC circulates around four months (110-120 days) before being destroyed. The destruction (i.e., hemolysis) of RBCs stimulates the bone marrow to produce reticulocytes which in turn mature to RBCs. Hemolytic anemia occurs when the production of new RBCs is unable to keep up with the destruction of RBCs.

Anemia is diagnosed by measuring the concentration of RBCs in the blood measured as either the concentration of hemoglobin or hematocrit (i.e., the percentage of blood that consists of RBCs). The cutoffs vary by sex and among various diagnostic criteria from <90g/L to <130g/L or a decrease of ≥ 20-30g/L (Hill 2019).

***Table 1. Proposed hemoglobin and hematocrit cutoffs from UpToDate and WHO (World Health Organization) recommendations.[1]***

| | Means & Brodsky (2024) | WHO (2024) |
|---|---|---|
| Female (hemoglobin) | <119g/L | <120g/L |
| Male (hemoglobin) | <136g/L | <130g/L |
| Female (hematocrit) | <35% | |
| Male (hematocrit) | <40% | |

[1] WHO recommendations from their published "Guideline on haemoglobin cutoffs to define anaemia in individuals and populations". The WHO recommendations are more complex than presented here, taking into account altitude, age, and other factors.

Sentinel

There is no individual test that reliably identifies hemolytic anemia. A diagnosis is determined from laboratory tests showing 1) an increased reticulocyte count (not accounted for by recent bleeding, repletion of nutrient deficiency [e.g., iron], or administration of the hormone erythropoietin); 2) signs of destruction of RBCs (e.g., increased lactate dehydrogenase [LDH], low haptoglobin, increased unconjugated bilirubin); and 3) no other obvious cause for anemia (e.g., pregnancy, altitude acclimatization) (Barcellini 2024). A peripheral blood smear can also help by identifying cells (or fragments of cells) with irregular shapes which provide clues to the underlying cause of hemolytic anemia.

Determining the etiology for hemolytic anemia is guided by exploring likely causes in the immediate medical context (e.g., transfusion reactions or drug-induced) and/or the administration of a peripheral blood smear and direct antiglobulin (i.e., Coombs) test (DAT). The findings on the blood smear may indicate:

- thrombotic microangiopathy (with schistocytes);
- congenital RBC membrane or cytoskeletal defect (with spheroctyes, elliptocytes, or stomatocytes); or
- hemoglobinopathy (with findings characteristic of sickle cell disease or thalassemia).

If the etiology remains unknown, the Coombs test can be used to determine whether the hemolytic anemia is due to an immune mechanism.

**Literature Review of Previous Validation Studies / Algorithms:**

- A Danish study (Hansen 2016) evaluated the PPV of ICD-10 diagnosis codes for hemolytic anemia from a population of 412 drawn from the Danish National Patient Register from 1994-2011. Of the 412 individuals, 249 had a diagnosis code for congenital HA and the remaining 163 had diagnosis codes for acquired HA (see Appendix 2). Validation was performed using “all the medical files and all available laboratory and pathology results” and uncertain diagnoses were reviewed by a second reviewer. For the “gold standard”, reviewers were required to confirm anemia and to identify hemolysis from laboratory results documenting an increase in reticulocytes with biochemical evidence of erythrocyte destruction (increased LDH, decreased haptoglobin, increased free hemoglobin, or hyperbilirubinemia), and no “better explanation” for the hemolysis (e.g., liver disease). The reviewers confirmed 359/412 (PPV: 87.1%) overall, and a PPV of 83.4% (136/163) for acquired HA.
- Maquet (2019) considered ICD-10 discharge diagnoses (see Appendix 2) from a single hospital in France in 2017 and validated HA codes by manual review of charts and laboratory results. For the “gold standard”, the reviewers were required to confirm anemia, a high reticulocyte count, and at least two of a) low serum haptoglobin levels, b) hyperbilirubinemia, and c) increased serum

LDH levels. The authors considered 453 patients, excluded 13 due to missing data, and found an overall PPV of 95% (418/440). For acquired HA, the authors report autoimmune HA (AIHA) as being confirmed in 49/53 individuals (PPV 92.5%). Sensitivity was not evaluated.

**Data Elements:**

| Necessary EHR Data | Useful EHR Data |
|---|---|
| Laboratory results (complete blood count [CBC], reticulocyte count, LDH, haptoglobin, unconjugated bilirubin) | Diagnosis codes |

**Clinical Complexity:** MEDIUM – Hemolytic anemia requires diagnosis of anemia and evidence of hemolysis in the absence of other explanatory factors.

Diagnosis of anemia requires the interpretation of lab results, particularly the CBC which provides hemoglobin and hematocrit measurements. These must be evaluated against one of several diagnostic criteria, and the importance of clinical interpretation is emphasized (Means & Brodsky 2024).

Once an anemia diagnosis has been reached, the clinician will need to consider the context to eliminate other obvious causes and will require additional measurements in order to determine the etiology:

- Unexplained high reticulocyte count,
- Evidence of RBC destruction such as:
  - An increase in unconjugated bilirubin,
  - An increase in LDH, or
  - Low haptoglobin.

Integrating these laboratory results can be tricky as they can show conflicting results.

The FFP clinical complexity worksheet has been included in Appendix 3.

**Data Complexity:** MEDIUM – A computable phenotype for HA requires results from at least five separate laboratory tests.

Validation of HA diagnosis codes had reasonable performance in Danish (Hansen 2016) and French (Maquet 2019) contexts, though no such study has yet been completed in the US. If a US validation study of diagnosis codes were to show insufficient performance, and a computable phenotype using EHR data were to be pursued, then the data complexity would be MEDIUM since a computable phenotype would require laboratory results from multiple tests. A CBC provides results that can indicate anemia, and at least four additional laboratory results will be required to confirm or rule out hemolysis including reticulocyte count, LDH, haptoglobin, and unconjugated bilirubin.

The FFP data complexity worksheet has been included in Appendix 4.

**Conclusion / Fitness-for-Purpose (FFP) Recommendation:** *MODERATE*

Based on promising results from European validation studies, a first step should be to conduct an ICD validation study using recent US data to determine if the hemolytic anemia diagnosis codes would be sufficient and a claims-based algorithm can be used. This validation work could be performed using data from an environment where both claims and EHR data are available, such as an integrated healthcare system or a data source that integrates insurance claims data with laboratory results.

If the US ICD validation study fails to meet expected performance, then the difficulty rating for creating a new computable phenotype that draws on EHR data would be MODERATE. An EHR-based computable phenotype for identification of HA requires extracting and integrating a number of laboratory results and eliminating other causes of anemia.

In developing a cohort for a study, other etiologies of hemolytic anemia must also be considered and excluded (e.g., metallic heart valves) as well hemolytic anemias not related to drug-induced hemolytic anemia (e.g., hereditary conditions like sickle cell anemia). We recommend that these efforts be integrated into study design and cohort selection rather than achieved as part of the computable phenotype.

#### References

Barcellini, W. (2024) Diagnosis of hemolytic anemia in adults, UpToDate. Available at: https://www.uptodate.com/contents/diagnosis-of-hemolytic-anemia-in-adults (Accessed: 5 December 2024).

Guideline on haemoglobin cutoffs to define anaemia in individuals and populations (2024). World Health Organization. Available at: https://www.who.int/publications/i/item/9789240088542 (Accessed: 16 January 2025).

Hansen, D. L. et al. (2016) 'Positive predictive value of diagnosis coding for hemolytic anemias in the Danish National Patient Register', Clinical epidemiology. Dove Medical Press Ltd., 8, pp. 241–252.

Hill, A. and Hill, Q. A. (2018) 'Autoimmune hemolytic anemia', Hematology. American Society of Hematology, 2018(1), pp. 382–389.

Hill, Q. A. et al. (2017) 'Guidelines on the management of drug-induced immune and secondary autoimmune, haemolytic anaemia', British journal of haematology. Br J Haematol, 177(2), pp. 208–220.

Hill, Q. A., Hill, A. and Berentsen, S. (2019) 'Defining autoimmune hemolytic anemia: a systematic review of the terminology used for diagnosis and treatment', Blood advances. American Society of Hematology, 3(12), pp. 1897–1906.

Jäger, U. et al. (2020) 'Diagnosis and treatment of autoimmune hemolytic anemia in adults: Recommendations from the First International Consensus Meeting', Blood reviews. Elsevier BV, 41(100648), p. 100648.

Maquet, J. et al. (2019) 'Validation of hemolytic anemia codes in the French hospital database', Blood. American Society of Hematology, 134(Supplement_1), pp. 3460–3460.

Means, R. T., Jr and Brodsky, R. A. (2003) Diagnosis of anemia in adults, UpToDate. Medical Library Association. Available at: https://www.uptodate.com/contents/diagnostic-approach-to-anemia-in-adults (Accessed: 5 December 2024).

Phillips, J. and Henderson, A. C. (2018) 'Hemolytic Anemia: Evaluation and Differential Diagnosis', American Family Physician, 98(6), pp. 354–361.

### Appendix 1: Society Guidelines for Hemolytic Anemia

There are guidelines for autoimmune hemolytic anemia (AIHA) issued by the American Society of Hematology (Hill and Hill 2018), the British Society of Hematology (Hill et al 2017), and International Consensus Recommendations (Jäger et al. 2020).

The American Society of Hematology recommends a stepwise approach in diagnosing AIHA when a patient presents with anemia.

- Identify laboratory and clinical evidence of hemolysis (the guidelines do not include specific cutoffs):
  - CBC
  - Reticulocyte count
  - Unconjugated bilirubin
  - Haptoglobin
  - Blood smear
  - Lactate dehydrogenase
- Determine immune nature of hemolysis using DAT
  - Positive DAT shows that hemolysis is autoimmune (not inherited)
- Exclude alternative causes (e.g., a positive DAT can occur from liver disease, chronic infection, and other conditions)

If no alternative explanations are found, a diagnosis of AIHA is reached. At this point, the guidelines point to identifying secondary causes and the type of AIHA.

### Appendix 2: ICD Diagnostic Codes Relevant to Hemolytic Anemia

This list of ICD-10 codes was compiled from those used by Hansen (2016) and Maquet (2019). It is not intended to be exhaustive but provided for reference purposes.

ICD-10

Code

| Code | Description | Hansen (2016) | Maquet (2019) |
|---|---|---|---|
| D55 | Anemia due to enzyme disorders | | x |
| D56 | Thalassemia | x (congenital) | x |
| D57 | Sickle cell anemia | x (congenital) | x |
| D588 | Hereditary spherocytosis | x (congenital) | x |
| D581 | Hereditary elliptocytosis | x (congenital) | x |
| D582 | Other hemoglobinopathies | x (congenital) | x |
| D589 | Hereditary hemolytic anemia, unspecified | x (congenital) | x |
| D590+592 | Drug-induced autoimmune and nonautoimmune hemolytic anemia | x (acquired) | x |
| D591 | Autoimmune hemolytic anemia (AIHA) | x (acquired) | x (PPV: 92.5) |
| D594 | Other nonautoimmune hemolytic anemias | x (acquired) | x |
| D595 | Paroxysmal nocturnal hemoglobinuria | x (acquired) | x |
| D599 | Acquired hemolytic anemia, unspecified | x (acquired) | x |

### Appendix 3: Clinical Complexity Worksheet

| Question or Characteristic | Easier | More Difficult |
|---|---|---|
| Medical specialty society or other clinical society guidelines | | |
| One or a few guidelines, clear, generally in agreement | ☒ Yes | |
| No guideline available[1] | | ☐ Yes |
| Multiple guidelines, conflicting or inconsistent | | ☐ Yes |
| Diagnosis is based on: | | |
| Objective measures or tests (e.g., lab results or imaging) | ☒ Yes | |
| Subjective findings (symptoms, physical exam findings) | | ☐ Yes |
| Diagnosis is based on 1 or 2 “gold standard” test results that are straightforward to interpret | ☐ Yes | |
| “Gold standard” is problematic or difficult to interpret; for instance, results may be ambiguous or the sensitivity of the test may be very low, leading to uncertainty | | ☐ Yes |
| Diagnosis requires considerable clinical judgement and/or interpretation of multiple pieces of data | | ☒ Yes |
| If different clinicians were given identical information, the agreement between them is likely to be: | | |
| High | ☒ Yes | |
| Low | | ☐ Yes |
| Does making the diagnosis require data from multiple points in time, such as multiple encounters with the patient or longitudinal lab results? | | ☐ Yes |
| Does making the diagnosis require integrating data from many different tests or sources? E.g., results from multiple lab tests, or different types of data (lab results, imaging, signs, symptoms) | | ☒ Yes |
| Can the diagnosis be made using 1-2 relatively common lab results that are available in structured or numeric form with clear, consistent cut off values? (e.g., neutropenia) | ☐ Yes | |
| Is it necessary to exclude competing diagnoses? (e.g., DILI) | | ☒ Yes |
| Does making the diagnosis require identifying and defining a baseline state and then a subsequent state? (e.g., Hepatitis B reactivation) | | ☐ Yes |

[1]The lack of a society guideline typically means greater clinical complexity because it indicates less certainty about how the HOI is diagnosed; however, in rare cases, the lack of a society guideline is due to a diagnosis being extremely straightforward.

Sentinel

#### Appendix 4: Data Complexity Worksheet

| Question or Characteristic | Easier | More Difficult |
|---|---|---|
| What type(s) of data elements are needed? | | |
| Laboratory test: | | |
| routine, results in structured form (e.g., creatinine, for acute kidney injury) | ☒ Yes | |
| less common (may be “send out” or stored as PDF) | | ☐ Yes |
| results may be free text or unstructured | | ☐ Yes |
| Imaging/procedure result: | | |
| more common finding, standardized language (e.g., pericardial effusion) | ☐ Yes | |
| less common finding, more variable language | | ☐ Yes |
| Clinical free text: | | |
| unstructured, signs or symptoms found in free text, (e.g., self-report of symptoms in a clinical note) | | ☐ Yes |
| How many different data elements are required to determine the diagnosis? | | |
| 1-2 | ☐ Yes | |
| 3+ | | ☒ Yes |
| Is natural language processing required? | ☒ No | ☐ Yes |
| Timing and number of laboratory or imaging results: | | |
| Does making the diagnosis require values from repeated laboratory or imaging results over time? | | ☐ Yes |
| Does the diagnosis rely on integrating results from several laboratory or imaging tests? (e.g., Hepatitis B reactivation) | | ☒ Yes |
| Does the diagnosis require comparing current lab or imaging results to a baseline? | | ☐ Yes |
| *If “Yes”*: is it likely the baseline lab or imaging results may be found in a different healthcare system? | | ☐ Yes |
| Are the desired data elements likely to be found in an image (e.g., a scanned PDF) rather than in structured data elements? (e.g., send-out labs, pulmonary function tests, EKGs) | | ☐ Yes |
| Are multiple algorithms required to identify a composite outcome? (e.g., serious infection) | | ☐ Yes |
| Is the interpretation of laboratory test results complex (e.g., hepatitis B reactivation)? | | ☒ Yes |
| For laboratory results: is this a commonly ordered lab that is expected to be available for most exposed patients? | ☒ Yes | |
| Are statistical models that integrate information from numerous features (e.g., sign, symptom, lab result) required to identify the outcome with adequate accuracy (e.g., anaphylaxis)? | | ☐ Yes |

### Fitness-for-Purpose (FFP) Assessment: Encapsulated Bacterial Infections

| | |
|---|---|
| **Health Outcome of Interest:** | Encapsulated Bacterial Infections |
| **Date:** | August 20, 2024 |
| **Subject:** | Computable Phenotype Feasibility Assessment for Encapsulated Bacterial Infections |
| **Drug(s)/ ARIA Insufficiency Memo(s):** | 1. Zinbryta (Daclizumab – High Yield Process), Memo #2016-588<br>2. Pegcetacoplan, Memo #2021-159<br>3. Sutimlimab, Memo #2022-67 |

#### Executive Summary

**Overall Assessment:** *HARD* – Computable algorithm would require results from only a few laboratory tests (bacterial culture or molecular diagnostic tests), which have a clear clinical interpretation. However, these tests have low diagnostic yield (very low sensitivity) and are often not ordered in the setting of an infection. Extracting information on pathogenic organisms from culture results may be difficult because these often are not reported in a standardized format. While technical aspects of creating a computable phenotype could be overcome, it is very unlikely that such a phenotype could successfully identify the HOI to support safety monitoring.

**HOI Description:** Encapsulated bacteria can cause serious infections. These bacteria are characteristically encased in a polysaccharide capsule which allows them to evade the host immune system and phagocytosis, enhancing their ability to cause disease. This memo focuses on three species of encapsulated bacteria that are most commonly implicated in serious infections: *Streptococcus pneumoniae, Neisseria meningitidis and Hemophilus influenzae.*

**ARIA Insufficiency Issue:** There is concern that drugs that inhibit the complement system may increase risk of serious infections caused by encapsulated bacteria. ARIA does not have algorithms to identify infections due to encapsulated bacteria.

**Existing Computable Algorithms:** ICD-9 and 10 codes exist that specify both the type of infection and the causative organism. Therefore, some infections caused by encapsulated bacteria may be identified from diagnosis codes; however, these codes' sensitivity is not established, and it is likely that many infections with these organisms will be missed.

**Clinical Complexity:** HIGH: A diagnosis is based on a few laboratory tests (culture, antigen test, or polymerase chain reaction [PCR]) that can identify the bacteria, and the identification of a pathogenic organism is considered diagnostic. However, these tests have low diagnostic yield (i.e., the organism may not grow from the cultured specimen), and therefore any algorithm is likely to have very low sensitivity.

**Data Complexity:** MEDIUM: Validation studies of ICD-9 codes for specific organisms are limited and new validation studies would be required for ICD-10 codes. Thus, the accuracy (especially the sensitivity) of these codes is not established. Results from needed laboratory tests are often not available because the tests are often not ordered. Natural Language Processing would be needed to extract data from culture results which may have heterogeneous language and structure and may not be uniformly available in the EHR.

**Conclusion / Fitness-for-Purpose Recommendation:** The validity of ICD codes has not been established, so access to laboratory results would be necessary. This may be challenging due to low availability, sensitivity, and interpretability of lab tests.

Computable phenotyping approaches cannot overcome the low diagnostic yield and incomplete availability of laboratory tests. Because encapsulated bacteria are the primary pathogens responsible for community-acquired bacterial pneumonia and bacterial meningitis, an alternative approach that is likely to be more successful is to evaluate medical product associations with these specific infections without requiring microbiological evidence of specific pathogens.

**Limitations:** This fitness-for-purpose recommendation is based on literature reviews as of July 2024. We assume an ideal performance of an algorithm equates to a PPV of at least 80% together with acceptable sensitivity.

#### Detailed Discussion

##### Description of HOI

Encapsulated bacteria can cause serious infections. These bacteria are characteristically encased in a polysaccharide capsule which allows them to evade the host immune system and phagocytosis, enhancing their ability to cause disease. The most important pathogenic encapsulated bacteria are:

- *Streptococcus pneumoniae*, which causes pneumococcal pneumonia, pneumococcal meningitis, and otitis media;[1]
- *Neisseria meningitidis*, which causes meningococcal meningitis, septicemia, and Waterhouse-Friderichsen syndrome;[2] and
- *Hemophilus influenzae*, which causes pneumonia, septicemia, meningitis, epiglottitis, cellulitis, and infectious arthritis.

An encapsulated bacterial infection is typically diagnosed through the detection of the bacteria by culture from the blood, cerebrospinal fluid (CSF), joint / bone, pleural fluid, or skin biopsy, or from a molecular diagnostic test (e.g., urinary antigen).[3]

##### Drug from Drug-Outcome Pair:

1) Zinbryta (Daclizumab – High Yield Process), Memo #2016-588
2) Pegcetacoplan, Memo #2021-159
3) Sutimlimab, Memo #2022-67

**Reason for ARIA insufficiency concern(s) related to outcome, according to FDA memo(s):** Memo #2022-67 notes that "ARIA has algorithms to identify serious infections," but not for "infections as specifically due to encapsulated bacteria."

##### Previous Validation Studies:

ICD-9 and -10 codes are available for a number of conditions related to *S. pneumoniae, N. meningitidis, and H. influenzae* infections (see Appendix 1). A search of the literature showed that most studies are focused on higher-level validation of the presence of infection (identified by groups of ICD-9 and ICD-10 diagnostic codes) and not the specific causative organism. These diseases can be caused by multiple pathogens, of which the encapsulated bacteria are a subset. There are some exceptions which are more specific to bacteria, but the literature is limited for studies that are specific to the three individual encapsulated bacteria of interest. Therefore, additional research is needed to understand how individual ICD codes will perform, specifically, ICD-10 codes for all three organisms and ICD-9 codes for *N. meningitidis*. The sensitivity of these codes is likely to be low compared with the true cases of serious infection caused by the pathogenic organism, both because the

[1] ICD10 (J13): https://www.icd10data.com/ICD10CM/Codes/J00-J99/J09-J18/J13-/J13
[2] ICD10 (A39.*): https://www.icd10data.com/ICD10CM/Codes/A00-B99/A30-A49/A39-
[3] https://www.uptodate.com/contents/diagnosis-of-meningococcal-infection/print

diagnostic yield of testing is low and because pathogen-specific ICD-9/10 codes may be infrequently used even when a specific organism has been identified.

*S. pneumoniae/ H. influenzae*

- Higgins et al. 2020: Cross-sectional study using data from 178 US hospitals in the Premier Healthcare Database. Patients were aged 18 years or older admitted with pneumonia and discharged between 2010 and 2015. Corresponding ICD-9 organism codes were used as indicators of diagnosis. Microbiological evidence of a pathogen was the gold standard (based on test results for blood or respiratory culture, urinary antigen, or polymerase chain reaction [PCR]). Compared to gold standard: 1) codes for *Streptococcus pneumoniae* (ICD-9 481, 482.30) had sensitivity: 60.1%, PPV: 73.4%, 2) codes for *Haemophilus influenzae* (ICD-9 482.2) had sensitivity: 42.8%, PPV: 84.7%.

No existing computable algorithms were located.

**Clinical Complexity:** *HIGH: Diagnosis is based on laboratory tests with clear interpretations (pathogen present or absent), but the tests have important limitations.*

The gold standard for the diagnosis of an encapsulated bacterial infection is the isolation of the bacterial species from a body fluid such as sputum, blood, cerebral spinal fluid, or urine. Isolation of the organism by culture confirms the etiology of infection. Other tests that can identify a specific organism include PCR and antigen testing, though the latter is reported to have low sensitivity and specificity for *N. meningitidis* and *H. influenzae*. Assuming that test results are available and provide meaningful information, it is straightforward to make the diagnosis. However, the diagnostic yield of these tests is low, and therefore the sensitivity of any computable phenotyping algorithm will also be low. It is likely that patients with certain characteristics or in certain clinical environments are more likely to undergo lab testing. An issue that raises clinical complexity specifically for cases of pneumonia (relevant to *S. pneumoniae* and *H. influenzae*) is the low likelihood of being able to identify an etiologic pathogen from the specimens typically taken.

**Data Complexity:** *MEDIUM: Natural Language Processing (NLP) needed to read free-text lab culture results which are likely to have heterogeneous language, spelling and formatting and which may often be missing.*

Data needed for identification of an encapsulated bacterial infection come from a few laboratory tests (culture, antigen test, or PCR). However, results from these tests are often not available in the chart because tests are inconsistently ordered. In addition, NLP would be needed to read lab culture results with heterogeneous language and structure. We expect these results to include different notations, spellings, misspellings and abbreviations of organisms' names, as well as variable numeric units, display formats and anatomical source information. There is evidence that identifying a specific organism from culture results using NLP with high validity is feasible, based on a study of another bacterium (methicillin-resistant Staphylococcus aureus; Frielin et al., 2008).

Although the NLP tasks are anticipated to be simpler than reading free text from clinical notes, key lab results may not be available in the EHR for a variety of reasons. It may take 3-5 days for an organism to grow in culture, by which time a patient may have been discharged home. If a patient was transferred from one hospital to another, culture results may be available only from the first hospital. Other challenges include inconsistent ordering of tests and low diagnostic yield, as discussed above. Thus, even an algorithm that accurately and completely captures laboratory evidence of encapsulated bacteria is likely to have very low sensitivity.

**Data Sources:**

| Necessary EHR Data | Useful EHR Data |
|---|---|
| Lab results (bacterial culture; antigen and PCR tests) | |

**Conclusion / Fitness-for-Purpose (FFP) Recommendation:**

While the technical aspects of creating a computable phenotype could be overcome, it is very unlikely that such a phenotype could successfully identify the outcome of interest to support safety monitoring. Phenotyping approaches are likely to have very low sensitivity for the identification of encapsulated bacteria, primarily because of the low diagnostic yield of testing. The low sensitivity is likely to introduce substantial bias. Because encapsulated bacteria are the primary pathogens responsible for community-acquired bacteria pneumonia and bacterial meningitis, an alternative approach to post-market surveillance that is likely to be more successful is to evaluate medical product associations with these specific infections without limiting outcome ascertainment to cases with microbiological evidence of specific pathogens.

### References

https://www.cdc.gov/hi-disease/about/index.html

Friedlin J, Grannis S, Overhage M. Using Natural Language Processing to Improve Accuracy of Automated Notifiable Disease Reporting. AMIA Annu Symp Proc. 2008; 2008: 207–211. PMID: 18999177. PMCID: PMC2656046

Higgins TL, Deshpande A, Zilberberg MD, Lindenauer PK, Imrey PB, Yu PC, Haessler SD, Richter SS, Rothberg MB. Assessment of the Accuracy of Using ICD-9 Diagnosis Codes to Identify Pneumonia Etiology in Patients Hospitalized With Pneumonia. JAMA Netw Open. 2020 Jul 1;3(7):e207750. doi: 10.1001/jamanetworkopen.2020.7750. PMID: 32697323; PMCID: PMC7376393.

Lo Re V, Carbonari DM, Jacob J, Short WR, Leonard CE, Lyons JG, Kennedy A, Damon J, Haug N, Zhou EH, Graham DJ, McMahill-Walraven C, Parlett LE, Nair V, Selvan M, Zhou Y, Pocobelli G, Maro JC, Nguyen MD. Validity of ICD-10-CM diagnoses to identify hospitalizations for serious infections among patients treated with biologic therapies. Pharmacoepi Drug Saf 2021; 30:899-909. Doi: 10.1002/pds.5253.Epub 2021 May 3.

### Appendix: ICD-9 and ICD-10 Diagnostic Codes Relevant to Encapsulated Bacterial Infections

This list of ICD-9 and -10 codes was compiled from Lo Re et al. 2021 and Higgins et al. 2020 with additional codes from http://www.icd9data.com. It is not intended to be exhaustive but instead is provided for reference purposes.

#### ICD-9

##### *Streptococcus pneumoniae*

| Code | Description | Source |
|---|---|---|
| 481 | Pneumococcal pneumonia [Streptococcus pneumoniae pneumonia] | http://www.icd9data.com |
| 482.30 | Pneumonia due to Streptococcus, unspecified | Higgins et al. 2020 |

##### *Neisseria meningitidis*

| Code | Description | Source |
|---|---|---|
| 036.0 | Meningococcal meningitis | http://www.icd9data.com |
| 036.3 | Waterhouse-Friderichsen syndrome, meningococcal | |
| 036.2 | Meningococcemia | |
| 036.4 | Meningococcal carditis | |
| 036.40 | Meningococcal carditis, unspecified | |
| 036.42 | Meningococcal endocarditis | |
| 036.43 | Meningococcal myocarditis | |
| 036.8 | Other specified meningococcal infections | |
| 036.1 | meningococcal encephalitis | |
| 036.81 | meningococcal optic neuritis | |
| 036.82 | meningococcal arthropathy | |
| 036.89 | other specified meningococcal infection | |
| 036.9 | meningococcal infection, unspecified | |

##### *Haemophilus influenzae*

| Code | Description | Source |
|---|---|---|
| 482.2 | Pneumonia due to Hemophilus influenzae | Higgins et al. 2020 |

**ICD-10**

***Streptococcus pneumonia***

| Code | Description | Source |
|---|---|---|
| A40.3 | Sepsis due to Streptococcus pneumoniae | Lo Re et al. 2021 |
| B95.3 | Streptococcus pneumoniae causing diseases classd elswhr | Lo Re et al. 2021 |
| J13 | Pneumonia due to streptococcus pneumoniae | Lo Re et al. 2021 |
| G00.1 | Pneumococcal meningitis | Lo Re et al. 2021 |

***Neisseria meningitidis***

| Code | Description | Source |
|---|---|---|
| A39.0 | Meningococcal meningitis | Lo Re et al. 2021 |
| A39.1 | Waterhouse-Friderichsen syndrome | Lo Re et al. 2021 |
| A39.2 | Acute meningococcemia | Lo Re et al. 2021 |
| A39.3 | Chronic meningococcemia | Lo Re et al. 2021 |
| A39.4 | Meningococcemia, unspecified | Lo Re et al. 2021 |
| A39.50 | Meningococcal carditis, unspecified | Lo Re et al. 2021 |
| A39.51 | Meningococcal endocarditis | Lo Re et al. 2021 |
| A39.52 | Meningococcal myocarditis | Lo Re et al. 2021 |
| A39.53 | Meningococcal pericarditis | Lo Re et al. 2021 |
| A39.81 | Meningococcal encephalitis | Lo Re et al. 2021 |
| A39.82 | Meningococcal retrobulbar neuritis | Lo Re et al. 2021 |
| A39.83 | Meningococcal arthritis | Lo Re et al. 2021 |
| A39.84 | Postmeningococcal arthritis | Lo Re et al. 2021 |
| A39.89 | Other meningococcal infections | Lo Re et al. 2021 |
| A39.9 | Meningococcal infection, unspecified | Lo Re et al. 2021 |

***Haemophilus influenzae***

| Code | Description | Source |
|---|---|---|
| A41.3 | Sepsis due to Hemophilus influenzae | Lo Re *et al.* 2021 |
| A49.2 | Hemophilus influenzae infection, unspecified site | Lo Re *et al.* 2021 |
| B96.3 | Hemophilus influenzae as the cause of diseases classd elswhr | Lo Re *et al.* 2021 |
| J14 | Pneumonia due to Hemophilus influenzae | Lo Re *et al.* 2021 |
| J20.1 | Acute bronchitis due to Hemophilus influenzae | Lo Re *et al.* 2021 |
| G00.0 | Hemophilus meningitis | Lo Re *et al.* 2021 |

### Fitness-for-Purpose (FFP) Assessment: Pulmonary Arterial Hypertension

| | |
|---|---|
| **Health Outcome of Interest:** | Pulmonary Arterial Hypertension |
| **Date:** | March 26, 2025 |
| **Subject:** | Health Outcome of Interest Feasibility Assessment for Pulmonary Arterial Hypertension |
| **Drug(s)/ ARIA Insufficiency Memo(s):** | FINTEPLA (Fenfluramine hydrochloride, ZX008), Memo #2020-953 |

#### Executive Summary

**Overall Assessment:** *HARD DIFFICULTY* – Developing computable algorithms for pulmonary arterial hypertension (PAH) would be challenging, because they would require substantial data from multiple sources including application of Natural Language Processing (NLP) to semi-structured electronic health records (EHR) and efforts to rule out multiple other causes of pulmonary hypertension for a definite diagnosis.

**ARIA Insufficiency Issue(s):** There's no validated claims-based algorithm in the published literature that can reliably identify patients with PAH, which is the type of PH likely to result from the medication of interest.

**Orientation to HOI:** PAH is one form of a broader condition known as pulmonary hypertension (PH), which refers to high blood pressure in the lungs due to various etiologies. In PAH, this increased pressure in the vessels is caused by obstruction in the small arteries in the lung for a variety of reasons.

**Existing Computable Algorithms:** No claims-based algorithms have been validated for identification of PAH in the general population. A machine learning algorithm using EHR data performed well in a PH-enriched population, but the transparency and transportability of the algorithm to other data sources and populations is questionable.

**Clinical Complexity:** *HIGH* - Despite objective measurements and clear guidelines for the diagnosis of PAH, the requirement of excluding multiple other causes of PH (e.g., chronic lung or heart disease) makes the diagnosis difficult.

**Data Complexity:** *MEDIUM* - The diagnosis of PAH requires measurements from right heart catheterization (RHC), which would require NLP algorithms to extract data from free text in EHR. Furthermore, the amount and complexity of data required for ruling out other causes of PH would be substantial, and such data may be stored in the EHR of a different healthcare system.

**Conclusion / Fitness-for-Purpose Recommendation:** There's no existing algorithm that can reliably capture PAH using claims data, i.e., a code-based algorithm, and it will be difficult to develop a computable phenotype algorithm for this outcome. Challenges include the need to exclude other groups of PH, which results in HIGH clinical complexity and HIGH data complexity.

**Limitations:** This fitness-for-purpose recommendation is based on literature reviews as of February 2025. We assume an ideal performance equates to a PPV of at least 80%.

#### Detailed Discussion

**Reason for ARIA insufficiency concern(s) related to outcome, according to FDA memo(s):** The claims-based algorithms used for identifying patients with PAH in the published literature varied considerably across studies and few are validated (Gillmeyer *et al*. 2019, Mathai *et al*. 2019). The algorithms requiring use of only PAH-related diagnosis codes or PAH-specific medications perform poorly. The most restrictive algorithms requiring use of PAH-related diagnosis codes together with PAH-specific medications and procedure codes (RHC or echocardiography) maximized PPV, but would miss patients who are either untreated or do not undergo RHC or echocardiography[1] during the study period (Mathai *et al*. 2019).

**Drug from Drug-Outcome Pair:**

FINTEPLA[2] (Fenfluramine hydrochloride, ZX008), Memo #2020-953

*Note: There is a Boxed warning and Warnings and Precautions on the association between serotonergic drugs with 5-HT2B receptor[3] agonist activity, including fenfluramine (the active ingredient in FINTEPLA), and PAH. Cardiac monitoring is required prior to starting treatment, during treatment, and after treatment with FINTEPLA.*

**Orientation to HOI**

Pulmonary arterial hypertension (PAH) is one form of a broader condition known as pulmonary hypertension (PH), which is high blood pressure in the lungs. PAH is referred to as Group 1: PAH among the five types of pulmonary hypertension. In PAH, the increased pressure in the blood vessels in the lungs is caused by obstruction in the small arteries in the lung for a variety of reasons.[4] The increased blood pressure in the lungs forces the heart to work harder to pump blood, which can eventually damage the heart's ability to effectively pump blood throughout the body.

**Literature Review of Previous Validation Studies / Algorithms:**

As concluded in the FDA memo assessing ARIA sufficiency, algorithms to identify PAH in administrative databases vary widely and few are validated (Gillmeyer *et al*. 2019, Mathai *et al*. 2019). Algorithms using only ICD codes perform poorly (PPV 3.3%-33.3%), and although algorithms requiring three components -- diagnosis codes,

---

[1] Because of the FINTEPLA REMS program, all patients treated with FINTEPLA will undergo regular echocardiograms, so this might not be an issue for FINTEPLA.

[2] Indicated for the treatment of seizures associated with Dravet syndrome and Lennox-Gastaut syndrome in patients 2 years of age and older.

[3] 5-HT2B receptor is involved in pulmonary vasoconstriction.

[4] American Lung Association: https://www.lung.org/lung-health-diseases/lung-disease-lookup/pulmonary-arterial-hypertension

PAH-specific medications, and RHC or echocardiogram-- maximize the PPV, they may miss patients who are either untreated or do not undergo RHC or echocardiography. Furthermore, all PAH algorithms using multiple components were designed to identify PAH patients from among patients with PH, which does not address the validity of the algorithm for capturing PAH patients among the general population. When restricted to the PH population, the PPV estimates are expected to be much higher than the PPV of the same algorithm when applied to the general population, which has much lower prevalence of PAH. This limitation also applies to studies published after the FDA memo (Gillmeyer *et al*. 2021, Jambon-Barbara *et al*. 2024).[5]

More recently, machine learning algorithms were applied to identify PH and/or PAH using EHR data (Thompson et al. 2023, Ong et al. 2020, Schuler et al. 2022). The Schuler 2022 study demonstrated good performance of a random forest algorithm for identifying PAH in Vanderbilt's Synthetic Derivative database. The model considered strength, persistence, and durability of ICD-9/10 codes for primary pulmonary hypertension, CPT codes for right heart catheterization, or PAH-specific medications.[6] ***However***, this algorithm was developed and tested in a population that is highly enriched for cases of PH, which may falsely inflate the PPV and sensitivity. In addition, the portability of this algorithm to claims-only databases is questionable due to varying coding practices. Although the study limited the features of the algorithms to variables that also exist in claims, the information in claims may still differ from that in EHR, which would impact the algorithm's performance. Furthermore, the paper does not include adequate information about which data were extracted to construct the features. The lack of operational details prevents other groups from replicating the approach. Furthermore, this algorithm draws on data recorded during follow-up of patients after PAH diagnosis, potentially for many months or even years. An important feature in the model is the number of mentions of diagnoses and medications throughout the medical record over time. This is a limitation in the surveillance setting when we want to accurately detect a disease soon after it develops.

---

[5] The best performing algorithm developed by Gillmeyer et al. reached sensitivity of 77.6%, specificity of 97.1%, PPV of 70.0% in data from the Veterans Health Administration and sensitivity of 78.9%, specificity of 95.0%, PPV of 86.0% in data from Boston Medical Center, specifically, a PAH referral center. However, this study was conducted among PAH-enriched populations.

[6] Strength was defined as the number of mentions throughout the medical record. Persistence was defined as the number of days that code or term stayed on the record. Durability was defined as the persistence divided by the length of the record after the first appearance on the record.

Data Sources:

| Necessary EHR Data | Useful EHR Data |
|---|---|
| RHC test report | Echocardiography |
| | Imaging reports |
| | Pulmonary function tests |
| | Arterial blood gases |
| | Laboratory test results |
| | Clinical notes (medical history) |
| | Diagnosis codes and medications |

**Clinical Complexity:** *HIGH*

PAH can be hard to diagnose because its symptoms could be caused by many other conditions.[7]

The main diagnostic algorithm for PH follows a three-step approach consisting of suspicion by first-line physicians, detection by echocardiography, and confirmation with right heart catheterization in a PH center. Patients typically present with exertional dyspnea and fatigue that progresses over time until severe PH with overt right ventricular (RV) failure develops. Therefore, the diagnosis is often delayed. It has been estimated that more than 20 percent of patients have symptoms of PH for longer than two years before it is recognized (Brown 2011). This is particularly common for patients younger than 36 years and those with coexisting medical conditions. These challenges make the diagnosis of PH difficult, and in addition, the diagnosis of PAH requires further classification of PH into one of 5 groups.

The final diagnosis of PAH, classified as Group 1 PH, requires right heart catheterization (RHC) to demonstrate a mean pulmonary arterial pressure (mPAP) ≥20 mmHg at rest and a pulmonary vascular resistance (PVR) ≥2 Wood Units.[8] Several additional criteria to exclude the remaining groups of PH (representing alternative mechanisms or underlying causes) must also be met:

- Mean pulmonary capillary wedge pressure (PCWP) ≤15 mmHg (to exclude PH due to left heart disease, Group 2 PH).
- Chronic lung diseases and other causes of hypoxemia are mild or absent (to exclude PH owing to chronic lung diseases or hypoxemia, Group 3 PH).

---

[7] Cleveland Clinic: https://my.clevelandclinic.org/health/diseases/23913-pulmonary-arterial-hypertension

[8] UpToDate. https://www.uptodate.com/contents/clinical-features-and-diagnosis-of-pulmonary-hypertension-of-unclear-etiology-in-adults

- Venous thromboembolic disease and pulmonary artery obstructions are absent (to exclude Group 4 PH).
- Certain other medical disorders are absent, including systemic disorders (e.g., sarcoidosis, chronic kidney disease ), hematologic disorders (e.g., myeloproliferative diseases and chronic hemolytic anemias), and metabolic disorders (e.g., glycogen storage disease). The purpose is to exclude PH with unclear multifactorial mechanisms, that is, Group 5 PH.

In addition, clinicians should ensure that all etiologies associated with PAH have been investigated. In addition to drug-induced PAH, other conditions associated with group 1 PAH include:

- Systemic disorders, such as connective tissue diseases, HIV, portal hypertension (most often due to chronic liver disease), congenital heart disease, schistosomiasis
- Heritable - PAH due to heritable genetic defects
- PAH with features of venous/capillary involvement

Therefore, despite the clear guidelines for the diagnosis of PAH, the requirements of excluding other conditions make the diagnosis difficult. It requires considerable clinical judgment and interpretation of multiple pieces of data to rule out other diseases.

The FFP clinical complexity worksheet has been included in Appendix 3.

**Data Complexity:** *HIGH*

The final diagnosis of PAH requires measurements from RHC which are typically documented as semi-structured EHR data. NLP algorithms will be required to extract this information from free text. In addition, data on medical history, diagnoses, and results from other procedures, laboratory tests and/or imaging results are needed to rule out other conditions, including PH caused by chronic lung diseases, venous thromboembolic disease, pulmonary artery obstructions, systemic disorders (e.g., sarcoidosis, chronic kidney disease), hematologic disorders (e.g., myeloproliferative diseases and chronic hemolytic anemias), and metabolic disorders (e.g., glycogen storage disease). The amount and complexity of data required for ruling out these conditions would be substantial, and data on historical medical conditions may be stored in the EHR of a different healthcare system. It is possible there are validated algorithms for some of these conditions. An important step would be to survey the literature to determine in which cases a diagnosis-code based algorithm would be sufficient; however, this is out of the scope for this report.

The FFP data complexity worksheet has been included in Appendix 4.

**Conclusion / Fitness-for-Purpose (FFP) Recommendation:** *HIGH DIFFICULTY*

There is no existing algorithm that can reliably capture PAH using claims data, and it will be difficult to develop a computable algorithm for this outcome. Challenges include the need to exclude other groups of PH, which results in HIGH clinical complexity and HIGH data complexity. NLP algorithms will be required to extract RHC results from semi-structured EHR procedure notes. A large number of data elements would be needed to rule out other groups of PH (caused by different underling conditions), which might be located in the EHRs of different healthcare systems. Therefore, it would be challenging to develop a computable algorithm for PAH with high PPV and sensitivity. There is potential for computable phenotyping to be useful for enhanced case identification, with subsequent medical record review to validate cases.

#### References

Gillmeyer KR, Lee MM, Link AP, Klings ES, Rinne ST, Wiener RS. Accuracy of Algorithms to Identify Pulmonary Arterial Hypertension in Administrative Data: A Systematic Review. Chest. 2019 Apr;155(4):680-688. doi: 10.1016/j.chest.2018.11.004. Epub 2018 Nov 22.

Mathai, S. C., A. R. Hemnes, S. Manaker, R. H. Anguiano, B. B. Dean, V. Saundankar, P. Classi, A. C. Nelsen, K. Gordon and C. E. Ventetuolo (2019). "Identifying Patients with Pulmonary Arterial Hypertension Using Administrative Claims Algorithms." Ann Am Thorac Soc 16(7): 797-806.

Gillmeyer KR, Nunez ER, Rinne ST, Qian SX, Klings ES, Wiener RS. Development and Validation of Algorithms to Identify Pulmonary Arterial Hypertension in Administrative Data. Chest. 2021 May;159(5):1986-1994. doi: 10.1016/j.chest.2020.12.010. Epub 2020 Dec 17.

Jambon-Barbara C, Hlavaty A, Bernardeau C, Bouvaist H, Chaumais MC, Humbert M, Montani D, Cracowski JL, Khouri C. Development and validation of a code-based algorithm using in-hospital medical records to identify patients with pulmonary arterial hypertension in a French healthcare database. ERJ Open Res. 2024 Aug 12;10(4):00109-2024. doi: 10.1183/23120541.00109-2024. eCollection 2024 Jul.

Thompson WE, Vidmar DM, Jessica K. Freitas K, Pfeifer JM, Fornwalt BK, Chen R, Altay G, Manghnani K, Nelsen AC, Morland K, Stumpe MC, Miotto R. Large Language Models with Retrieval-Augmented Generation for Zero-Shot Disease Phenotyping. Deep Generative Models for Health Workshop NeurIPS 2023. https://arxiv.org/abs/2312.06457

Ong MS, Klann JG, Lin KJ, Maron BA, Murphy SN, Natter MD, Mandl KD. Share Claims-Based Algorithms for Identifying Patients With Pulmonary Hypertension: A Comparison of Decision Rules and Machine-Learning Approaches. J Am Heart Assoc. 2020 Oct 20;9(19):e016648. doi: 10.1161/JAHA.120.016648. Epub 2020 Sep 29.

Schuler KP, Hemnes AR, Annis J, Farber-Eger E, Lowery BD, Halliday SJ, Brittain EL. An algorithm to identify cases of pulmonary arterial hypertension from the electronic medical record. Respir Res. 2022 May 28;23(1):138. doi: 10.1186/s12931-022-02055-0.

Brown LM, Chen H, Halpern S, et al. Delay in recognition of pulmonary arterial hypertension: factors identified from the REVEAL Registry. Chest 2011; 140:19.

### Appendix 1: Society Guidelines for Pulmonary Arterial Hypertension in the United States

2022 ESC/ERS Guidelines for the Diagnosis and Treatment of Pulmonary Hypertension: Developed by the Task Force for the Diagnosis and Treatment of Pulmonary Hypertension of the European Society of Cardiology (ESC) and the European Respiratory Society (ERS). Eur Heart J 2022; 43, 3618-3731

https://doi.org/10.1093/eurheartj/ehac237

A multistep, pragmatic approach to diagnosis should be considered in patients with unexplained dyspnoea or symptoms/signs raising suspicion of PH. This strategy is depicted in detail in the Figure below. Characteristic diagnostic features of patients with PAH are shown in the table.

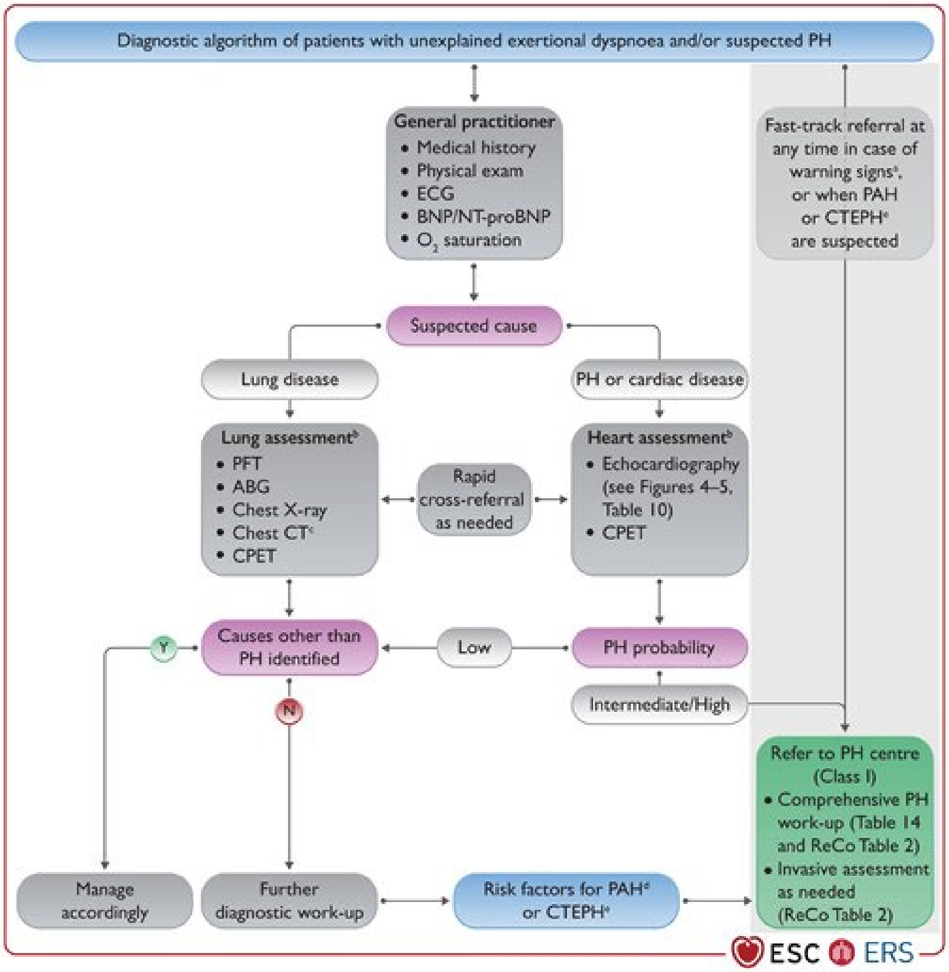

*Figure 1. Diagnostic algorithm of patients with unexplained dyspnoea and/or suspected pulmonary hypertension.*

*Table 1. Characteristic diagnostic features of patients with different forms of pulmonary hypertension.*

| Diagnostic tool | Characteristic findings/features | Group 1 (PAH) |
|---|---|---|
| 5.1.1 Clinical presentation | Clinical features | Variable age, but young, female patients may be predominantly affected. Clinical presentation depends on associated conditions and phenotype See Section 5.1.1 |
| | Oxygen requirement for hypoxaemia | Uncommon, except for conditions with low DLCO or right-to-left shunting |
| 5.1.3 Chest radiography | | RA/RV/PA size ↑<br>Pruning of peripheral vessels |
| 5.1.4 Pulmonary function tests and ABG | Spirometry/PFT impairment | Normal or mildly impaired |
| | DLCO | Normal or mild-to-moderately reduced (low DLCO in SSc-PAH, PVOD, and some IPAH phenotypes) |
| | Arterial blood gas<br>$PaO_2$<br>$PaCO_2$ | Normal or reduced<br>Reduced |
| 5.1.5 Echocardiography | | Signs of PH (increased sPAP, enlarged RA/RV)<br>Congenital heart defects may be present |
| 5.1.6 Lung scintigraphy | Planar – SPECT V/Q | Normal or matched |
| 5.1.7 Chest CT | | Signs of PH or PVOD |
| 5.1.11 Cardiopulmonary exercise testing | | High $VE/VCO_2$ slope<br>Low $P_{ET}CO_2$, decreasing during exercise<br>No EOV |
| 5.1.12 Right heart catheterization | | Pre-capillary PH |

### Appendix 2: ICD Diagnostic Codes Relevant to Pulmonary Arterial Hypertension

This list of ICD-9 and -10 codes was compiled from various relevant literature, with additional codes from https://icdlist.com/icd-10/codes/other-pulmonary-heart-diseases-i27. It is not intended to be exhaustive, but provided for reference purposes.

**ICD-9**

| Code | Description |
|---|---|
| 416.0 | Primary pulmonary hypertension |
| 416.8 | Other chronic pulmonary heart diseases |
| 416.9 | Chronic pulmonary heart disease, unspecified |

**ICD-10**

| Code | Description |
|---|---|
| 127.0 | Primary pulmonary hypertension (idiopathic PAH) |
| 127.20 | Pulmonary hypertension, unspecified |
| 127.21 | Secondary pulmonary arterial hypertension |
| 127.22 | Pulmonary hypertension due to left heart disease |
| 127.23 | Pulmonary hypertension due to lung diseases and hypoxia |
| 127.24 | Chronic thromboembolic pulmonary hypertension |
| 127.29 | Other secondary pulmonary hypertension |
| 127.83 | Eisenmenger syndrome |
| 127.89 | Other specified pulmonary heart diseases |
| 127.9 | Pulmonary heart disease, unspecified |

#### Appendix 3: Clinical Complexity Worksheet

| Question or Characteristic | Easier | More Difficult |
|---|---|---|
| Medical specialty society or other clinical society guidelines | | |
| One or a few guidelines, clear, generally in agreement | ☒ Yes | |
| No guideline available[1] | | ☐ Yes |
| Multiple guidelines, conflicting or inconsistent | | ☐ Yes |
| Diagnosis is based on: | | |
| Objective measures or tests (e.g., lab results or imaging) | ☒ Yes | |
| Subjective findings (symptoms, physical exam findings) | | ☐ Yes |
| Diagnosis is based on 1 or 2 "gold standard" test results that are straightforward to interpret | ☐ Yes | |
| "Gold standard" is problematic or difficult to interpret; for instance, results may be ambiguous or the sensitivity of the test may be very low, leading to uncertainty | | ☐ Yes |
| Diagnosis requires considerable clinical judgement and/or interpretation of multiple pieces of data | | ☒ Yes |
| If different clinicians were given identical information, the agreement between them is likely to be: | | |
| High | ☒ Yes | |
| Low | | ☐ Yes |
| Does making the diagnosis require data from multiple points in time, such as multiple encounters with the patient or longitudinal lab results? | | ☐ Yes |
| Does making the diagnosis require integrating data from many different tests or sources? E.g., results from multiple lab tests, or different types of data (lab results, imaging, signs, symptoms) | | ☒ Yes |
| Can the diagnosis be made using 1-2 relatively common lab results that are available in structured or numeric form with clear, consistent cut off values? (e.g., neutropenia) | ☐ Yes | |
| Is it necessary to exclude competing diagnoses? (e.g., DILI) | | ☒ Yes |
| Does making the diagnosis require identifying and defining a baseline state and then a subsequent state? (e.g., Hepatitis B reactivation) | | ☐ Yes |

[1]The lack of a society guideline typically means greater clinical complexity because it indicates less certainty about how the HOI is diagnosed; however, in rare cases, the lack of a society guideline is due to a diagnosis being extremely straightforward.

#### Appendix 4: Data Complexity Worksheet

| Question or Characteristic | Easier | More Difficult |
|---|---|---|
| What type(s) of data elements are needed? | | |
| Laboratory test: | | |
| routine, results in structured form (e.g., creatinine, for acute kidney injury) | ☒ Yes | |
| less common (may be "send out" or stored as PDF) | | ☒ Yes |
| results may be free text or unstructured | | ☒ Yes |
| Imaging/procedure result: | | |
| more common finding, standardized language (e.g., pericardial effusion) | ☒ Yes | |
| less common finding, more variable language | | ☐ Yes |
| Clinical free text: | | |
| unstructured, signs or symptoms found in free text, (e.g., self-report of symptoms in a clinical note) | | ☐ Yes |
| How many different data elements are required to determine the diagnosis? | | |
| 1-2 | ☐ Yes | |
| 3+ | | ☒ Yes |
| Is natural language processing required? | ☐ No | ☒ Yes |
| Timing and number of laboratory or imaging results: | | |
| Does making the diagnosis require values from repeated laboratory or imaging results over time? | | ☐ Yes |
| Does the diagnosis rely on integrating results from several laboratory or imaging tests? (e.g., Hepatitis B reactivation) | | ☒ Yes |
| Does the diagnosis require comparing current lab or imaging results to a baseline? | | ☐ Yes |
| *If* "Yes": is it likely the baseline lab or imaging results may be found in a different healthcare system? | | ☐ Yes |
| Are the desired data elements likely to be found in an image (e.g., a scanned PDF) rather than in structured data elements? (e.g., send-out labs, pulmonary function tests, EKGs) | | ☐ Yes |
| Are multiple algorithms required to identify a composite outcome? (e.g., serious infection) | | ☐ Yes |
| Is the interpretation of laboratory test results complex (e.g., hepatitis B reactivation)? | | ☐ Yes |

| | | |
|---|---|---|
| For laboratory results: is this a commonly ordered lab that is expected to be available for most exposed patients? | ☐ Yes | |
| Are statistical models that integrate information from numerous features (e.g., sign, symptom, lab result) required to identify the outcome with adequate accuracy (e.g., anaphylaxis)? | | ☐ Yes |
